# Assessing the Burden of COVID-19 in Developing Countries: Systematic Review, Meta-Analysis, and Public Policy Implications

**DOI:** 10.1101/2021.09.29.21264325

**Authors:** Andrew Levin, Nana Owusu-Boaitey, Sierra Pugh, Bailey K. Fosdick, Anthony B. Zwi, Anup Malani, Satej Soman, Lonni Besançon, Ilya Kashnitsky, Sachin Ganesh, Aloysius McLaughlin, Gayeong Song, Rine Uhm, Daniel Herrera-Esposito, Gustavo de los Campos, Ana Carolina Pecanha Antiono, Enyew Birru Tadese, Gideon Meyerowitz-Katz

## Abstract

**Introduction:** The infection-fatality rate (IFR) of COVID-19 has been carefully measured and analyzed in high-income countries, whereas there has been no systematic analysis of age-specific seroprevalence or IFR for developing countries.

**Methods:** We systematically reviewed the literature to identify all COVID-19 serology studies in developing countries that were conducted using population representative samples collected by early 2021. For each of the antibody assays used in these serology studies, we identified data on assay characteristics, including the extent of seroreversion over time. We analyzed the serology data using a Bayesian model that incorporates conventional sampling uncertainty as well as uncertainties about assay sensitivity and specificity. We then calculated IFRs using individual case reports or aggregated public health updates, including age-specific estimates whenever feasible.

**Results:** Seroprevalence in many developing country locations was markedly higher than in high-income countries. In most locations, seroprevalence among older adults was similar to that of younger age cohorts, underscoring the limited capacity that these nations have to protect older age groups. Age-specific IFRs were roughly 2x higher than in high-income countries. The median value of the population IFR was about 0.5%, similar to that of high-income countries, because disparities in healthcare access were roughly offset by differences in population age structure.

**Conclusion:** The burden of COVID-19 is far higher in developing countries than in high-income countries, reflecting a combination of elevated transmission to middle-aged and older adults as well as limited access to adequate healthcare. These results underscore the critical need to accelerate the provision of vaccine doses to populations in developing countries.

**Key Points:**

- Age-stratified infection fatality rates (IFRs) of COVID-19 in developing countries are about twice those of high-income countries.
- Seroprevalence (as measured by antibodies against SARS-CoV-2) is broadly similar across age cohorts, underscoring the challenges of protecting older age groups in developing countries.
- Population IFR in developing countries is similar to that of high-income countries, because differences in population age structure are roughly offset by disparities in healthcare access as well as elevated infection rates among older age cohorts.
- These results underscore the urgency of disseminating vaccines throughout the developing world.

## Introduction

An important unknown during the COVID-19 pandemic has been the relative severity of the disease in developing countries compared to higher-income nations. The incidence of fatalities in many developing countries appeared to be low in the early stages of the pandemic, suggesting that the relatively younger age structure of these countries might have protected them against the harms of the disease. More recently, however, it has become clear that the perceived differences in mortality may have been illusory, reflecting poor vital statistics systems leading to underreporting of COVID-19 deaths (4, 5). Moreover, relatively low mortality outcomes in developing countries would be starkly different from the typical pattern observed for many other communicable diseases, reflecting the generally lower access to good-quality healthcare in these locations (6, 7).

As shown in Table 1, mortality attributable to COVID-19 in many developing locations exceeds 2,000 deaths per million. Of the ten nations with the highest number of deaths attributed to COVID-19, seven are developing countries. Furthermore, these statistics may understate the true death toll in a number of lower- and middle income countries. Numerous studies of excess mortality have underscored the limitations of vital registration and death reporting, particularly in developing countries (4, 5, 8-12). For example, recent studies of India have found that actual deaths from COVID-19 were about ten times higher than those in official reports (5, 8). Similarly, a study in Zambia found that only 1 in 10 of those who died with COVID-19 symptoms and whose post-mortem COVID-19 test was positive were recorded as COVID-19 deaths in the national registry (13). Strikingly, the continuation of that study has demonstrated the catastrophic impact of COVID-19 in Zambia, raising the overall mortality by as much as five to ten times relative to a normal year (14).

**Table 1:**
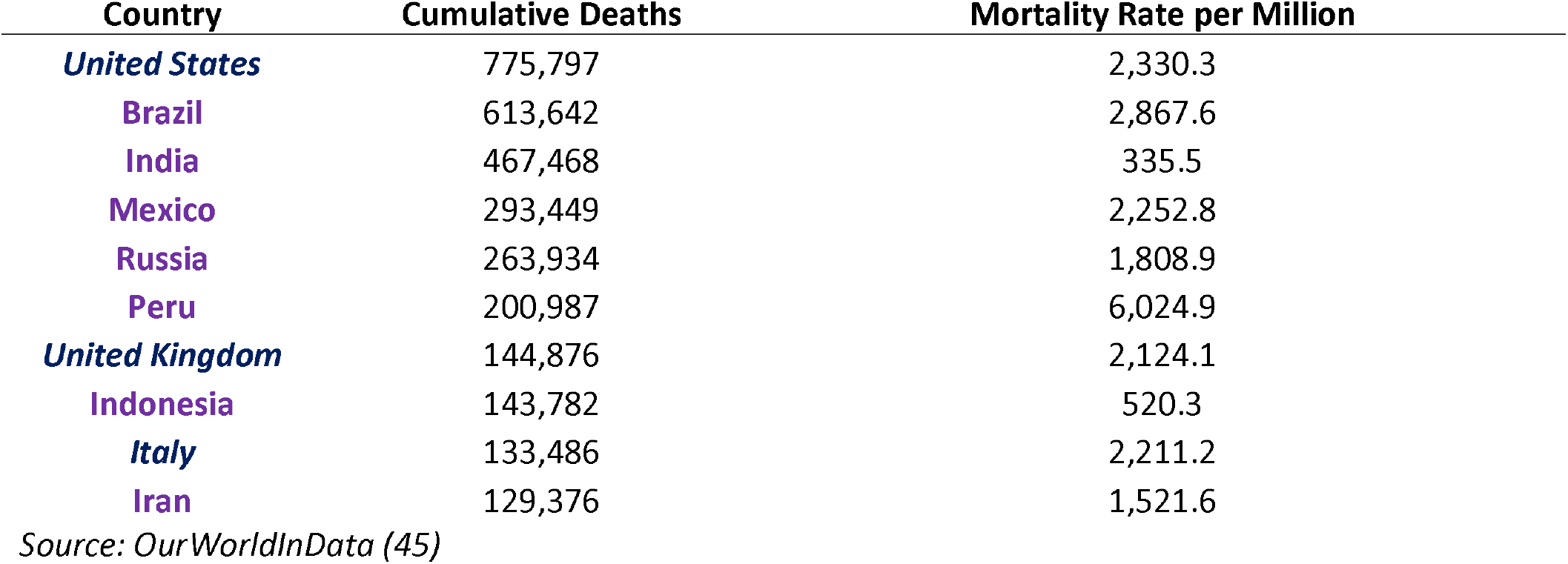
Confirmed COVID-19 Deaths as of 25 Nov 2021.

| Country | Cumulative Deaths | Mortality Rate per Million |
| --- | --- | --- |
| *United States* | 775,797 | 2,330.3 |
| *Brazil* | 613,642 | 2,867.6 |
| *India* | 467,468 | 335.5 |
| *Mexico* | 293,449 | 2,252.8 |
| *Russia* | 263,934 | 1,808.9 |
| *Peru* | 200,987 | 6,024.9 |
| *United Kingdom* | 144,876 | 2,124.1 |
| *Indonesia* | 143,782 | 520.3 |
| *Italy* | 133,486 | 2,211.2 |
| *Iran* | 129,376 | 1,521.6 |

Source: OurWorldInData (45)

There has, however, been a relative dearth of systematic research concerning the early experience of COVID-19 and the associated infection fatality rate (IFR) in developing countries. Previous evaluations have largely focused on assessing these patterns in high-income countries, where high quality data on seroprevalence and fatalities has been readily available throughout the pandemic (15, 16). In particular, seroprevalence studies conducted in high-income countries in 2020 found low overall prevalence of antibodies to COVID-19 (generally less than 10%) (17), with much lower prevalence among older adults compared to younger cohorts. Analysis of these data has clearly underscored the extent to which the IFR of COVID-19 increases exponentially with age, that is, the disease is far more dangerous for middle-aged and older adults compared to children and young people (3, 15, 16). Two prior meta-analytic studies have considered variations in IFR by age but did not consider the possibility that IFR in developing locations might differ systematically from high-income countries due to healthcare quality, access, and other socioeconomic factors (15, 18).

## Objectives

1. Determine overall prevalence of COVID-19 infection in locations in developing countries
2. Assess age-specific patterns of seroprevalence in these locations
3. Estimate age-specific IFRs and compare to benchmark values for high-income countries
4. Investigate possible reasons for differences in population IFR between locations

## Methods

To perform this meta-analysis, we collected published papers, preprints, and government reports of COVID-19 serology studies for which all specimens were collected before March 1^st^ 2021 and that were publicly disseminated by July 14, 2021. The full search methodology is given in the supplementary appendices. The study was registered on the Open Science Foundation: https://osf.io/edpwv/

We restricted the scope of our analysis to locations in developing countries using the classification system of the International Monetary Fund (IMF); that is, we excluded locations that the IMF classifies as “high-income countries.” (19). In some contexts developing countries are also described as low-to middle-income countries or as emerging and developing economies.

### Inclusion/Exclusion Criteria

Our analysis only included studies that had a random selection of participants from a sample frame representative of the general population (20, 21). Consequently, studies of convenience samples – such as blood donors or residual sera from commercial laboratories – were excluded. Such samples are subject to intrinsic selection biases that may vary across different settings and hence would detract from systematic analysis of the data Indeed, there is abundant evidence from the pandemic that convenience samples provide inaccurate estimates of seroprevalence, with assessments indicating that they are likely to overestimate the true proportion infected (22, 23).

A crucial part of our analysis entailed adjusting raw seroprevalence to reflect the sensitivity and specificity of the particular assay used in each serology study, and to construct credible intervals that reflect uncertainty about assay characteristics as well as conventional sampling uncertainty. Where a reported study did not include that information, we requested it from study authors. Other data needed and extracted for the analysis included start and end dates of specimen collection, the specific assay used, and age-specific serology data.

See the supplementary appendices for further details on inclusion and exclusion criteria.

### Deaths

For locations with publicly-available databases of all individual cases, we tabulated the fatality data to match the age brackets of that serology study, using cumulative fatalities as of 14 days after the midpoint date of specimen collection to reflect the time lags between infection, seropositivity, and fatal outcomes. In the absence of individual case data, we searched for contemporaneous public health reports and tabulated cumulative deaths as of 28 days after the midpoint date of specimen collection to incorporate the additional time lags associated with real-time reporting of COVID-19 fatalities (see supplementary appendices for detailed modelling concerning death lags).

Matching prevalence estimates with subsequent fatalities is not feasible if a serology study was conducted in the midst of an accelerating outbreak. Therefore, as in previous work (3), we estimated seroprevalence but did not analyze IFRs for locations where the cumulative death toll increased by 3x or more over the four-week period following the midpoint date of specimen collection. For details, see the supplementary appendices. In instances where we were not able to match deaths to serology data, or there were accelerating outbreaks, we used this information to look at serology only.

Additionally, we extracted data on excess deaths for all countries that were included in our analysis. We used two primary sources of estimates on excess mortality: the Institute for Health Metrics and Evaluation (IHME) (2) and the World Mortality Dataset (WMD) (4). The IHME produces national or regional estimates of excess mortality for every location included in this review, while the WMD has estimates for a subset of those locations. We then computed the ratio of excess mortality to reported fatalities for each location in order to assess the impact of potential death underreporting, and calculated adjusted IFRs using excess mortality as the numerator, as well as the ratio between IFRs calculated using reported and excess deaths. We used excess mortality as it is likely to represent the true burden of COVID-19 accurately in developing nations (4).

### Statistical Analysis

We use a Bayesian modelling framework to simultaneously estimate age-specific prevalence and infection fatality rates (IFRs) for each location in our study. First 2e model age-specific prevalence for each location at the resolution of the serology data reported. Then, we model the number of people that test positive in a given study location and age group as coming from a binomial distribution with a test positivity probability that is a function of the true prevalence, sensitivity and specificity, accounting for seroconversion and seroreversion (see the supplementary appendices).

As in Carpenter and Gelman (2020) (24), we consider sensitivity and specificity to be unknown and directly model the lab validation data (e.g., true positives, true negatives, false positives, false negatives) for each test. Independent weakly informative priors are placed on the seroprevalence parameters, and independent, informative priors akin to those in Carpenter and Gelman (2020) (24) are placed on the sensitivity and specificity parameters. To avoid assumptions about the variability of prevalence across age within a serology age bin, we aggregate deaths for each location to match their respective serology age bins. Independent mildly informative priors are assumed on the age group specific IFR parameters.

Prevalence for a given age group and location is estimated by the posterior mean and equal-tailed 95% credible interval. Uniform prevalence across age is deemed plausible for locations where the 95% credible intervals for the ratio of seroprevalence for age 60 and older over the seroprevalence estimate for ages 20 to 60 contains 1.

### IFR Calculation and Comparison

We model the number of individuals at a given location and age group that are reported dying of COVID-19 as Poisson distributed with rate equal to the product of the age group IFR, age group population, and age group prevalence. For locations where deaths were reported separately for different age bins this model provides IFR estimates for specific age groups and for broader population cohorts, including adults aged 18-65 years. For locations where death data was not disaggregated by age the model provides a population IFR. The model was implemented in the programming language R, with posterior sampling computation implemented with the Stan software package (25).

To perform a meta-analysis of age-specific IFRs across locations, we conduct a meta-regression with random effects. In the metaregression, the dependent variable is the estimated IFR for a specific age group in a specific geographical location, the explanatory variable is the median age of that particular age group, and the standard deviation of each idiosyncratic error is taken from the Bayesian analysis described above. We used a random-effects procedures to allow for residual heterogeneity between studies and across age groups by assuming that these divergences are drawn from a Gaussian distribution. We also allowed for fixed effects by location, to account for locations that deviate from the norm. Since the metaregression used IFR estimates based on reported deaths, we compared the location-specific fixed effects to two estimates of the ratio of excess mortality to COVID-19 deaths in each location. We also compared these metaregression results to a prior metaregression of age-specific IFR for high-income countries (3). This was performed using the meta regress procedure in Stata v17.

### Covariates

We selected covariates that were judged likely to have an impact either on the IFR of COVID-19 itself or on the accuracy of official data on COVID-related mortality based on prior research and expertise. Where possible, we extracted these covariates at a state or regional level within a country, otherwise they were identified at national level. A full list of covariates and the method of extraction can be found in the supplementary appendices. In instances where a covariate was only available at the national level, we aggregated location-specific seroprevalence and IFRs by weighting each location using the square root of the number of serology specimens collected in that location.

## Results

We identified a total of 2,384 study records, with 2,281 records identified from online databases and a further 103 from Twitter and Google Scholar. After excluding 2,062 records we assessed 322 records for inclusion in the final analyses. There were a total of 89 studies that could be used to describe either seroprevalence or IFR. The final sample for IFR estimates included 62 estimates from 25 developing countries. The search and exclusion process can be seen in the supplementary appendices. The distribution of included seroprevalence estimates can be seen in Figure 1. A full list of studies included in the IFR calculations can be found in Table 2, and the full list of studies and links to each study can be seen on our Github repository https://covid-ifr.github.io/.

**Figure 1.**
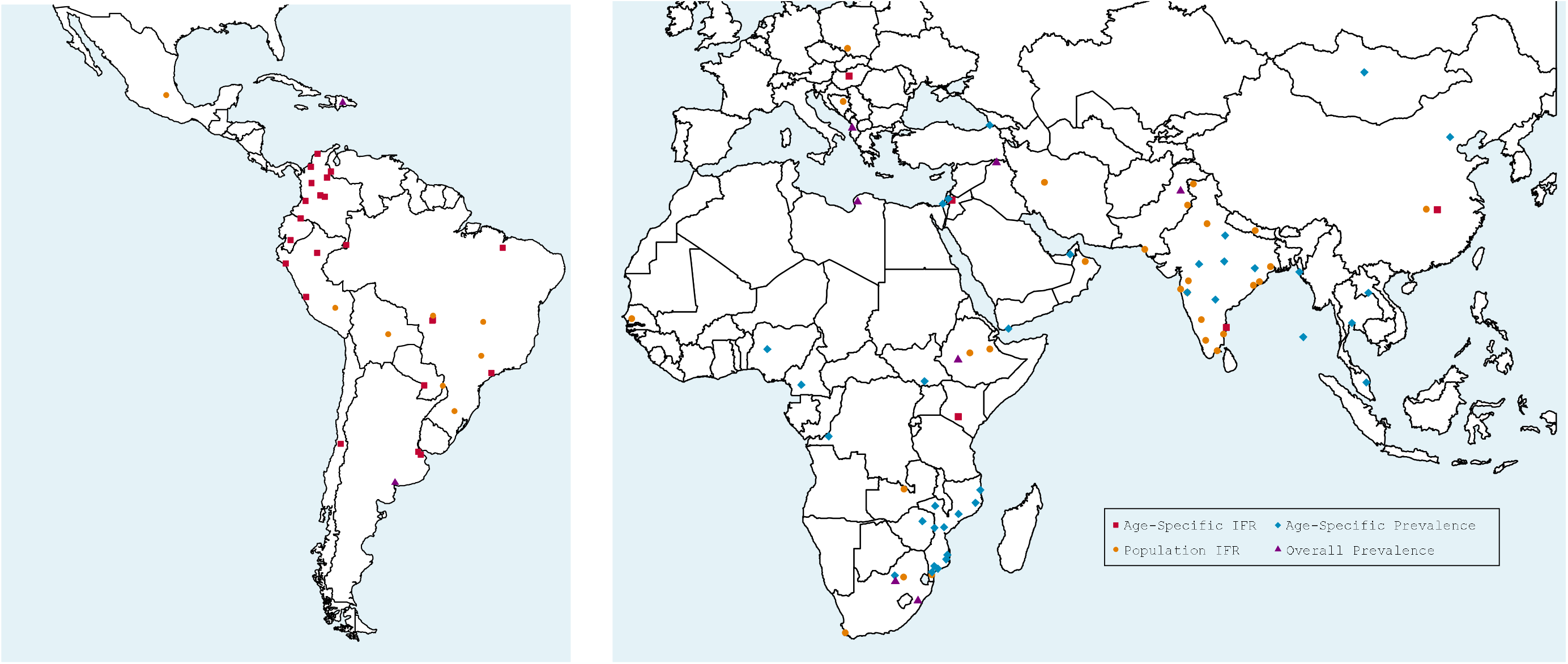
Map of Study Locations. Map of study locations with specifics of how these locations were used in the study. St. Petersburg, Russia (not shown on the map) has total IFR data.

**Table 2.**
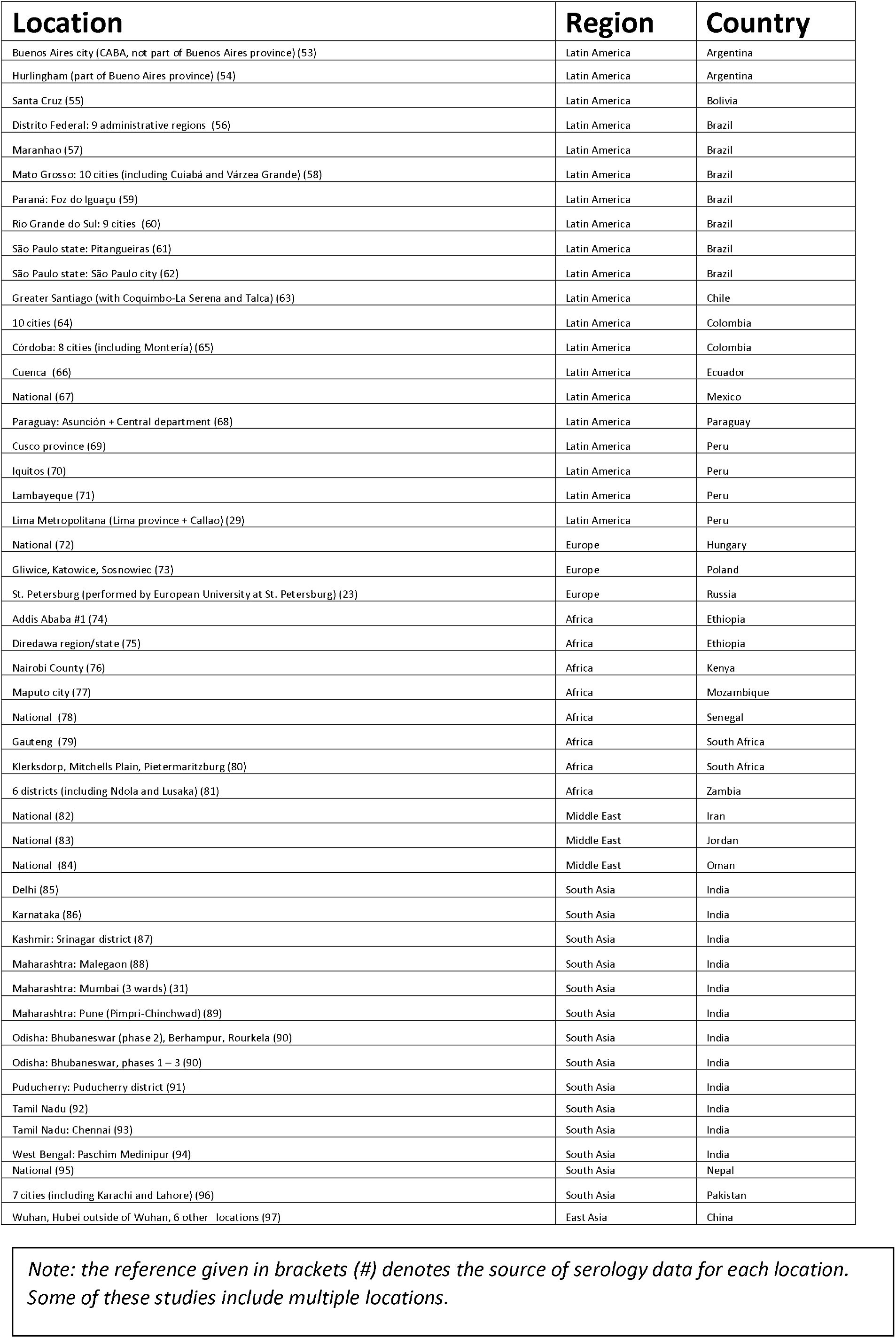
Included Studies for IFR.

| Location | Region | Country |
| --- | --- | --- |
| Buenos Aires city (CABA, not part of Buenos Aires province) (53) | Latin America | Argentina |
| Hurlingham (part of Buenos Aires province) (54) | Latin America | Argentina |
| Santa Cruz (55) | Latin America | Bolivia |
| Distrito Federal: 9 administrative regions (56) | Latin America | Brazil |
| Maranhao (57) | Latin America | Brazil |
| Mato Grosso: 10 cities (including Cuiabá and Várzea Grande) (58) | Latin America | Brazil |
| Paraná: Foz do Iguaçu (59) | Latin America | Brazil |
| Rio Grande do Sul: 9 cities (60) | Latin America | Brazil |
| São Paulo state: Pitangueiras (61) | Latin America | Brazil |
| São Paulo state: São Paulo city (62) | Latin America | Brazil |
| Greater Santiago (with Coquimbo-La Serena and Talca) (63) | Latin America | Chile |
| 10 cities (64) | Latin America | Colombia |
| Córdoba: 8 cities (including Montería) (65) | Latin America | Colombia |
| Cuenca (66) | Latin America | Ecuador |
| National (67) | Latin America | Mexico |
| Paraguay: Asunción + Central department (68) | Latin America | Paraguay |
| Cusco province (69) | Latin America | Peru |
| Iquitos (70) | Latin America | Peru |
| Lambayeque (71) | Latin America | Peru |
| Lima Metropolitana (Lima province + Callao) (29) | Latin America | Peru |
| National (72) | Europe | Hungary |
| Gliwice, Katowice, Sosnowiec (73) | Europe | Poland |
| St. Petersburg (performed by European University at St. Petersburg) (23) | Europe | Russia |
| Addis Ababa #1 (74) | Africa | Ethiopia |
| Diredawa region/state (75) | Africa | Ethiopia |
| Nairobi County (76) | Africa | Kenya |
| Maputo city (77) | Africa | Mozambique |
| National (78) | Africa | Senegal |
| Gauteng (79) | Africa | South Africa |
| Klerksdorp, Mitchells Plain, Pietermaritzburg (80) | Africa | South Africa |
| 6 districts (including Ndola and Lusaka) (81) | Africa | Zambia |
| National (82) | Middle East | Iran |
| National (83) | Middle East | Jordan |
| National (84) | Middle East | Oman |
| Delhi (85) | South Asia | India |
| Karnataka (86) | South Asia | India |
| Kashmir: Srinagar district (87) | South Asia | India |
| Maharashtra: Malegaon (88) | South Asia | India |
| Maharashtra: Mumbai (3 wards) (31) | South Asia | India |
| Maharashtra: Pune (Pimpri-Chinchwad) (89) | South Asia | India |
| Odisha: Bhubaneswar (phase 2), Berhampur, Rourkela (90) | South Asia | India |
| Odisha: Bhubaneswar, phases 1 – 3 (90) | South Asia | India |
| Puducherry: Puducherry district (91) | South Asia | India |
| Tamil Nadu (92) | South Asia | India |
| Tamil Nadu: Chennai (93) | South Asia | India |
| West Bengal: Paschim Medinipur (94) | South Asia | India |
| National (95) | South Asia | Nepal |
| 7 cities (including Karachi and Lahore) (96) | South Asia | Pakistan |
| Wuhan, Hubei outside of Wuhan, 6 other locations (97) | East Asia | China |

*Note: the reference given in brackets (#) denotes the source of serology data for each location. Some of these studies include multiple locations.*

### Seroprevalence

As shown in Figure 2, numerous locations in developing countries had relatively high levels of seroprevalence during the study period (March 2020 thru February 2021). That pattern is strikingly different from the outcomes in high-income countries, where seroprevalence generally remained below 20%.

**Figure 2.**
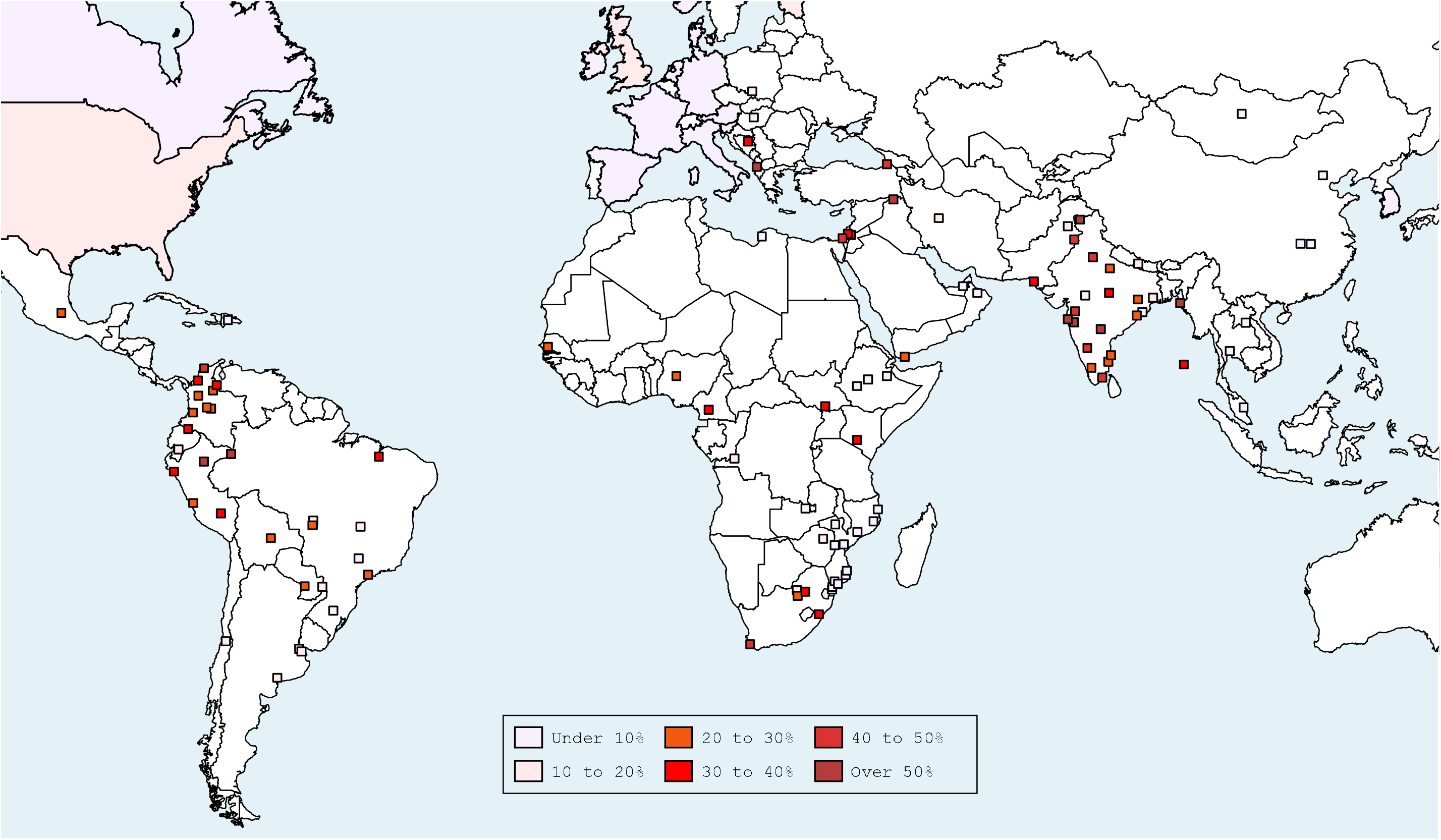
Seroprevalence during the studied period of the COVID-19 Pandemic. Map of areas with seroprevalence during the studied period. St. Petersburg, Russia (not shown on the map) had measured seroprevalence of 11% as of June 2020. This represents the same seroprevalence as used in IFR calculations.

In most developing country locations, seroprevalence was roughly uniform across age strata. Figure 3 shows the heatmap of age-specific seroprevalence across all age cohorts. As shown in Figure 4, the ratio of seroprevalence for older adults (ages 60+ years) compared to middle-aged adults (ages 40 to 59 years) is indistinguishable from unity in most of these locations. While many locations had a ratio below 1, the majority of areas were very substantially above the ratio for higher-income areas (green shaded region), and the point-estimates were not markedly below 1, indicating minimal difference in infection rates between older and younger adults in developing nations, and a markedly higher rate than that observed in higher-income countries.

**Figure 3.**
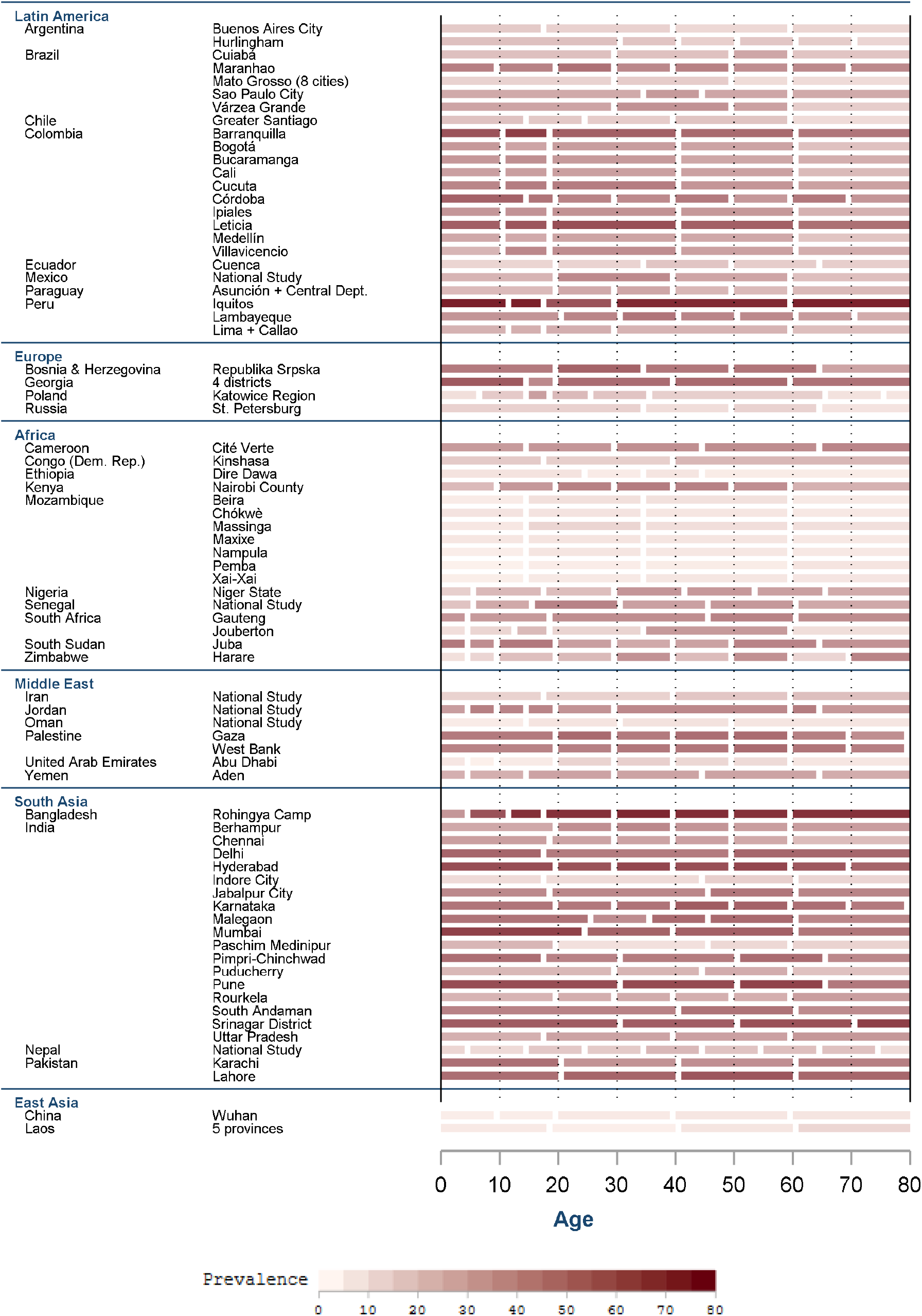
Age-Specific Seroprevalence by Location. Map of areas with seroprevalence in the studied period

**Figure 4.**
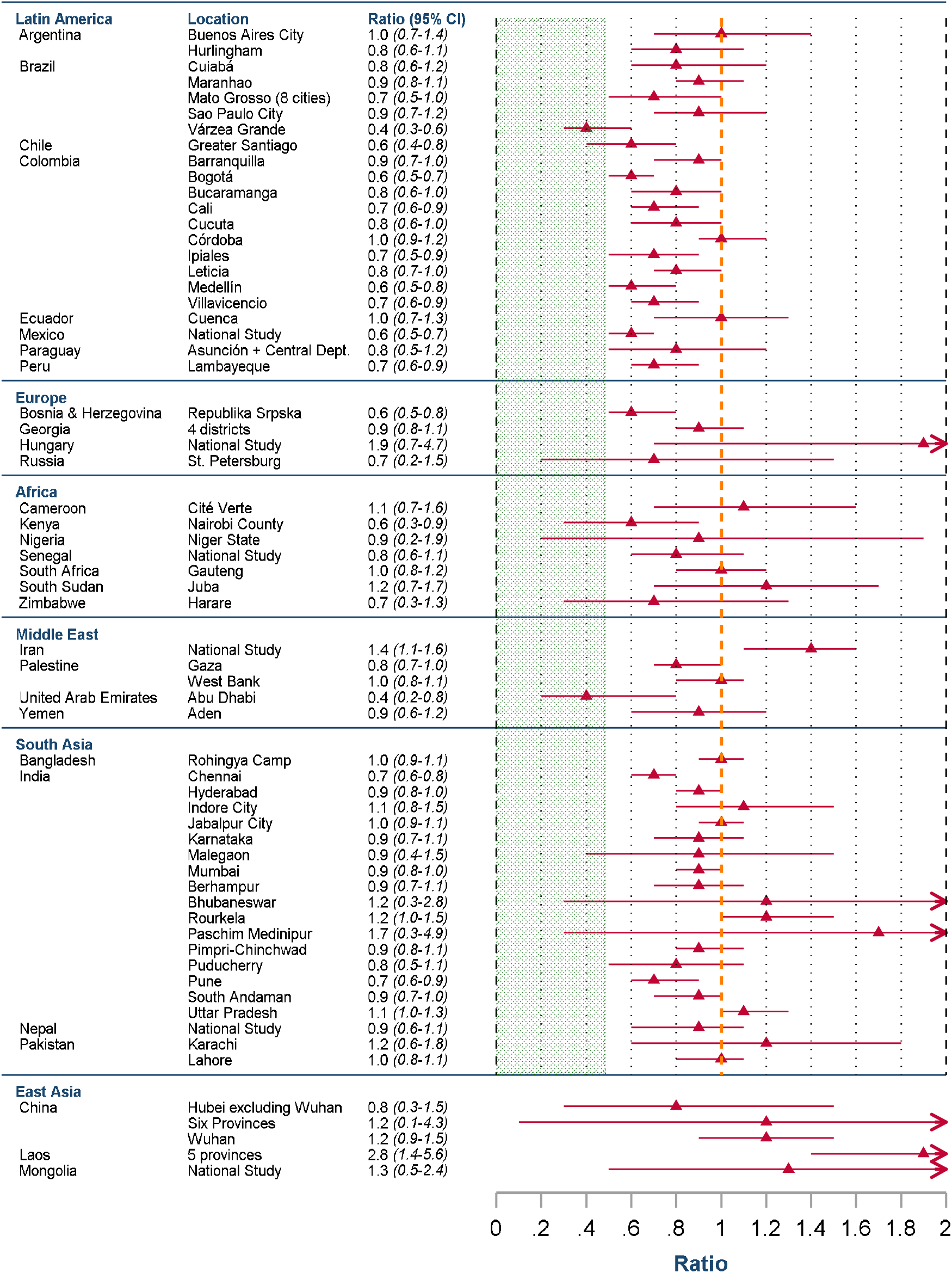
Ratio of Seroprevalence for Older Adults (60+ years) Compared to Adults (40-59 years) Green shaded area – range of high-income nations for ratio during the studied period (3), orange line – ratio of 1.

**Figure 5.**
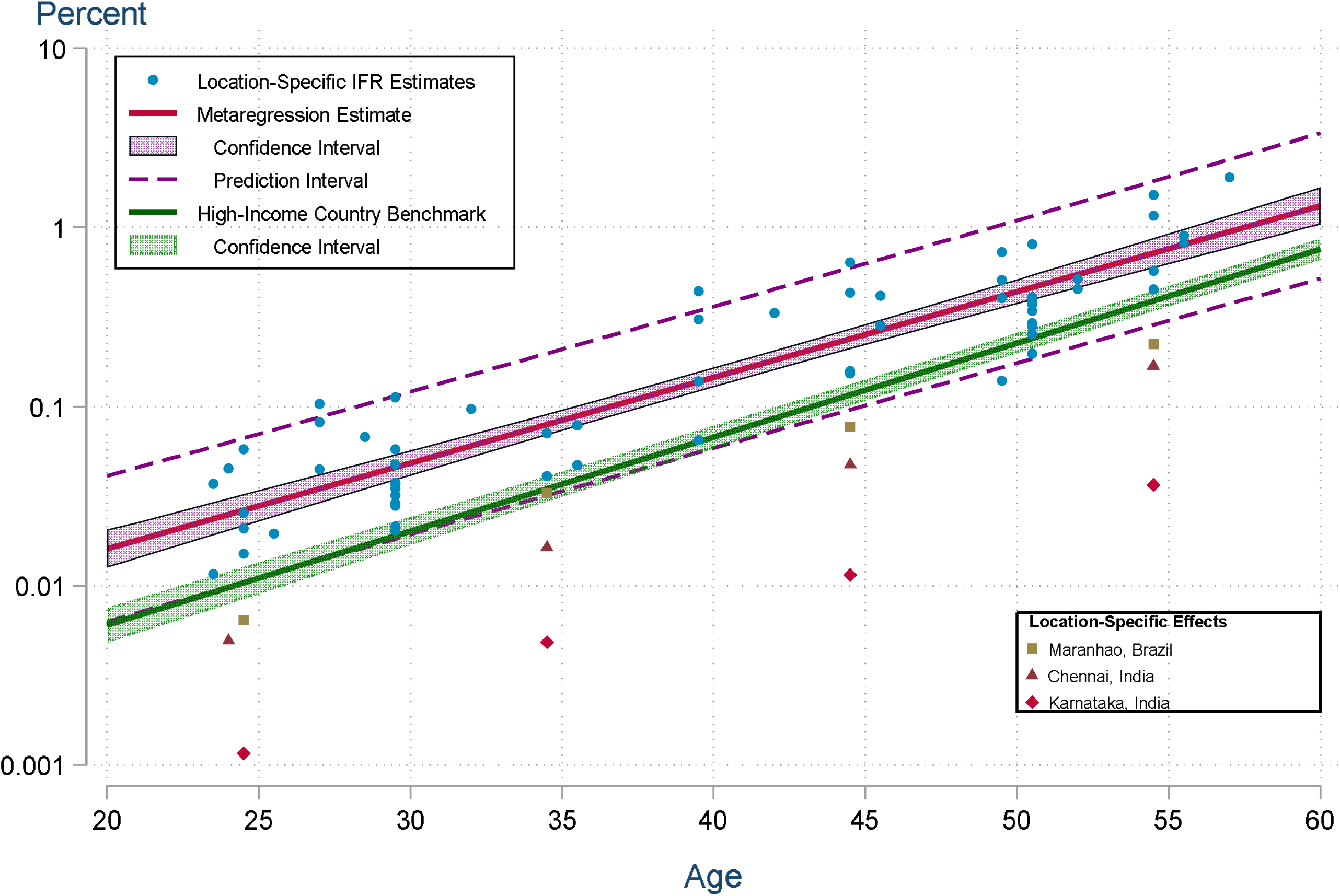
Metaregression Results. IFR estimates for the population aged 18-65 across locations.

### Infection Fatality Ratios

Our statistical analysis produced age-specific IFRs and confidence intervals for 28 locations, and population IFRs for those locations as well as an additional 27 places. The full results of this analysis are shown in the supplementary appendices. We obtain the following metaregression results:

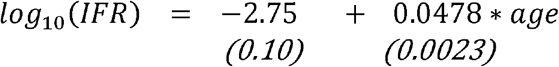

Here the standard error for each estimated coefficient is given in parentheses. These estimates are highly significant with t-statistics of -28.7 and 21.0, respectively, and p-values below 0·0001. The residual heterogeneity τ^2^ = 0.039 (p-value < 0.0001) and I^2^ = 92.5, confirming that the random effects are essential for capturing unexplained variations across studies and age groups. The adjusted R^2^ is 91.1%. Location-specific fixed effects are only distinguishable from zero for three locations: Maranhão, Brazil (−0.50); Chennai, India (−0.68); and Karnataka, India (−1.29).

The metaregression results can be seen in Figure 6. Nearly all of the observations fall within the 95% prediction interval. The importance of the location-specific effects is readily apparent. Indeed, these effects imply that the age-specific IFRs for Maranhão are about 1/3 of the metaregression prediction, while those for Chennai and Karnataka are 1/5 and 1/20, respectively.

**Figure 6.**
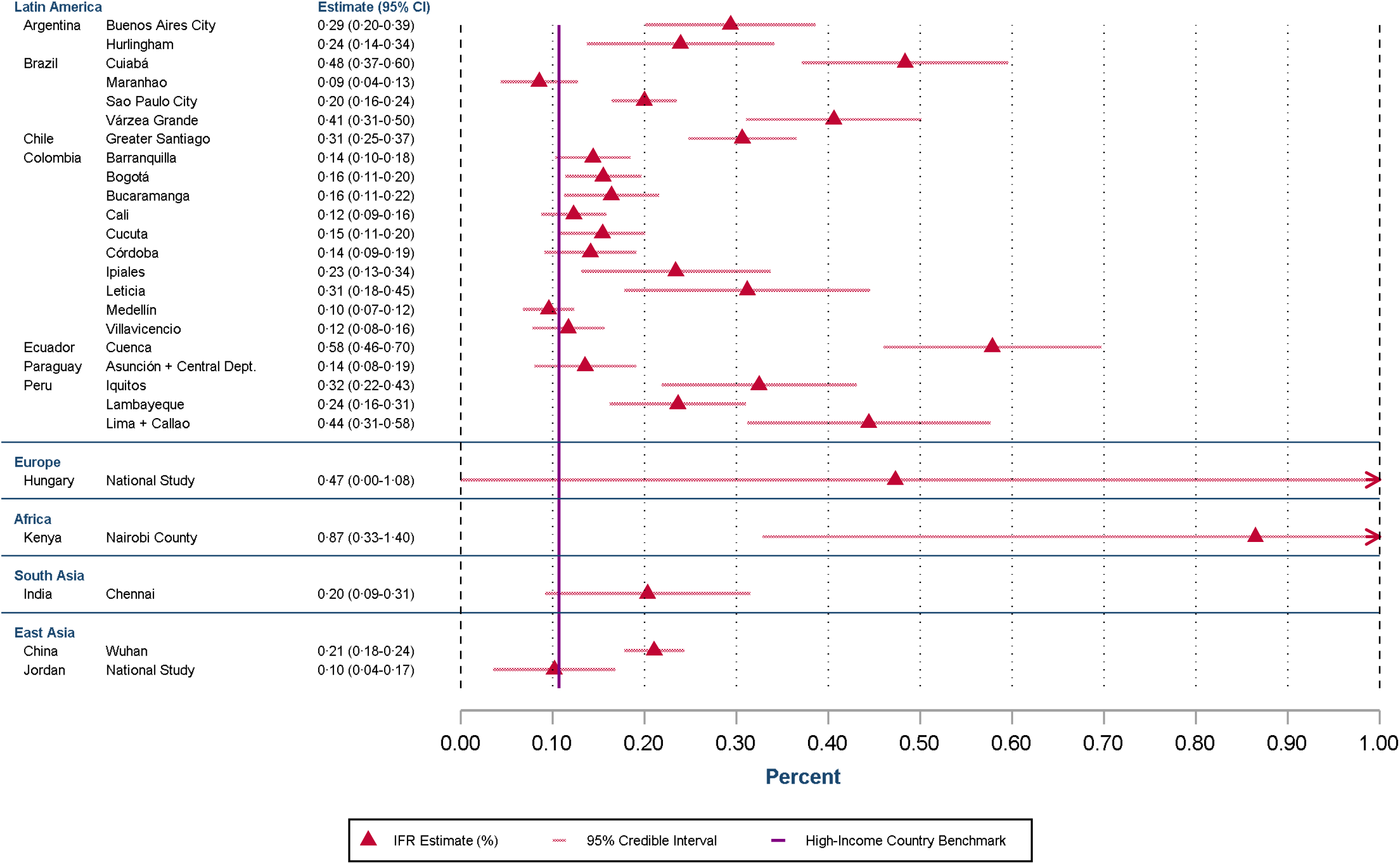
Standardized Population IFR for Ages 18-65. IFR estimates for the population aged 18-65 across locations. These population IFRs are standardized to match the age structure of a typical developing country for ages 18-65

The benchmark metaregression of high-income countries has a slope of 0.0524 (CI: 0.0499-0.0549) (3). A Welch test strongly rejects the hypothesis of equality in the slope parameters for developing countries vs. high-income countries with a p-value<0.0001.

This metaregression analysis uses age-specific IFRs based on reported COVID-19 deaths in each location. As a cross-check, table 3 reports the ratio of excess mortality to reported deaths for each of these locations.

**Table 3.**
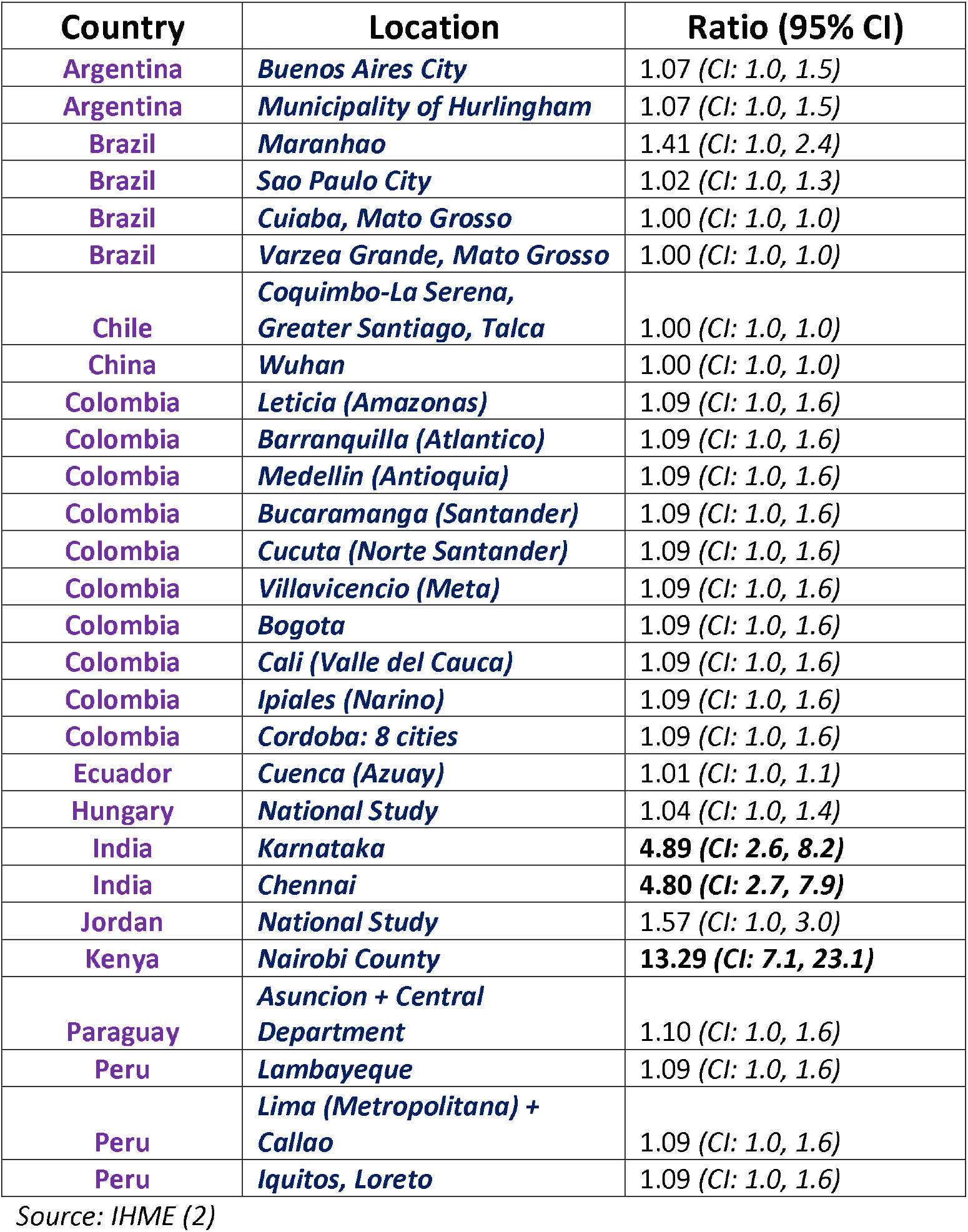
Ratio of Excess Mortality to Reported COVID-19 Deaths.

For nearly all of these locations, the ratio is indistinguishable from unity; that is, reported COVID-19 deaths are broadly consistent with the evidence from excess mortality assessments. There were three exceptions (Chennai, Karnataka, and Nairobi, Kenya), two of which had significant location-specific effects in the metaregression.

The precision of IFR estimates varied by age. At lower age groups, the number of deaths becomes very small, and thus the uncertainty is large regarding the IFR. Conversely, at older ages the number of infections and deaths can be very small in countries with extremely small populations of those aged over 65, and thus these estimates are also uncertain. The full figures across all ages can be found in the supplementary appendices.

IFR estimates are presented in figure 6 for each location. Here the age-adjusted IFR estimates for each location were weighted based on the location specific prevalence of each age group and a common baseline population structure so that these population IFR estimates are comparable across locations with differing population structure. We also adjust for excess mortality using the ratios shown in Table 3.

### Assessment of Death Reporting

For the full set of locations for which population IFR can be assessed, we found that the adequacy of death certification was highly significant in explaining cross-country variations. As shown in Figure 7, the median value of population IFR was about 0.5% in countries where a majority of deaths were well-certified (using SDG assessments (1) conducted prior to the pandemic) compared to only 0.05% in countries with lower proportions of well-certified deaths. In the latter set of countries, adjustments for excess mortality shift the population IFR upwards by an order of magnitude, to a median of 0.6%. Indeed, the population IFR for Zambia increases from 0.23% to 1.96% – the highest value of any country in our sample. In contrast, the excess mortality adjustments make relatively little difference for countries with a majority of well-certified deaths.

**Figure 7.**
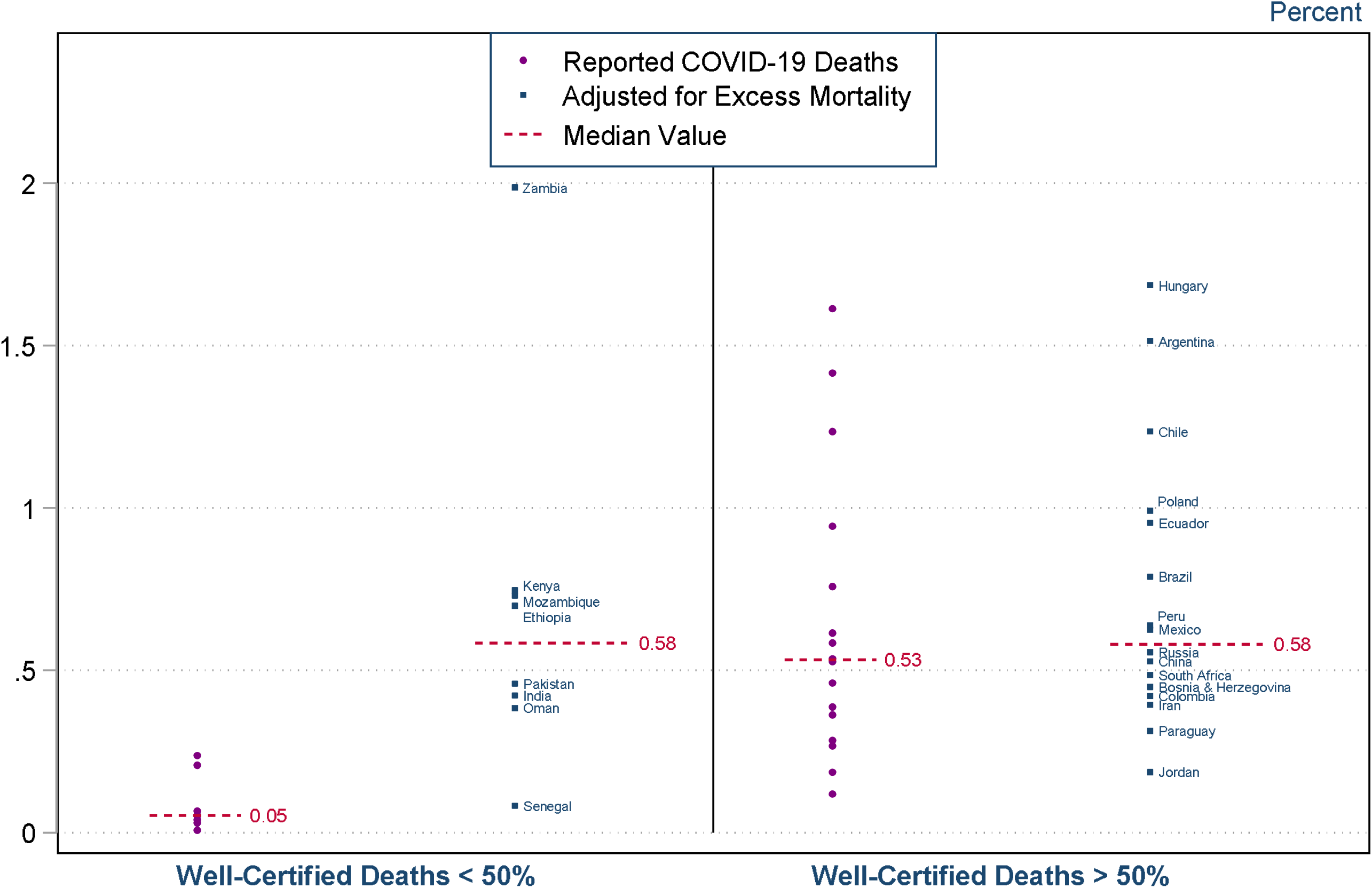
Population IFR and Well-Certified Death Registrations. Population IFR for regions divided into areas with <50% well-certified deaths and areas with >50% well-certified deaths as per SDGs (1) (purple) and these IFRs adjusted for estimates of excess mortality (blue). For two locations (Bolivia and Nepal) information on well-certified deaths was over ten years old and so these countries were excluded.

Finally, we considered the extent to which the adjusted measures of population IFR were robust to alternative estimates of the ratio of excess mortality to reported deaths. As shown in Figure 8 the estimates from IHME and WMD were generally well aligned, with just a small number of exceptions. The adjusted population IFRs had a median value of 0.49% using the IHME estimates and 0.58% using the WMD estimates.

**Figure 8.**
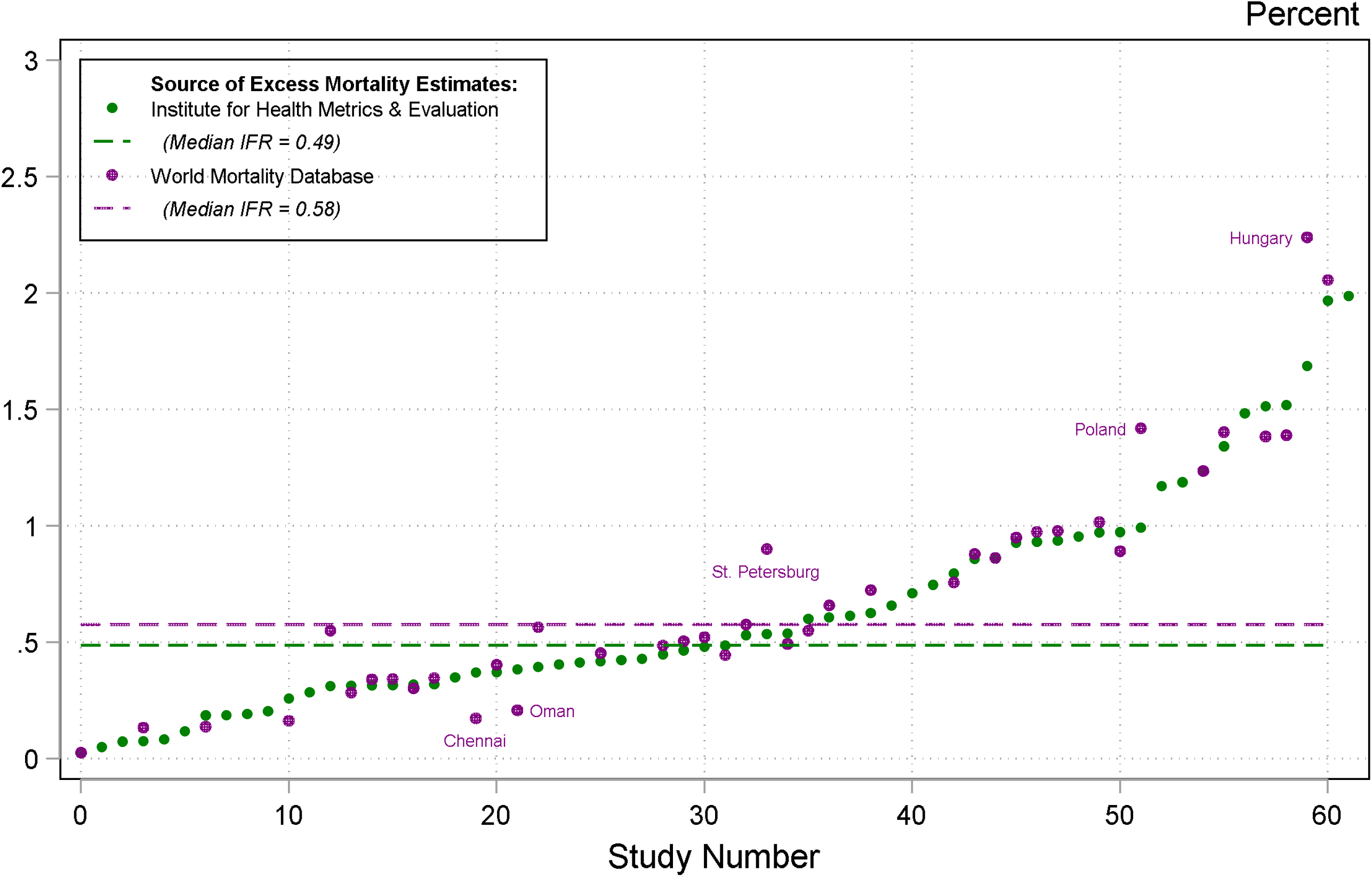
Excess Mortality Adjusted Population IFRs. Population IFR for regions adjusted with either IHME (2) or WMD (4) estimates for mortality. Population IFRs adjusted for excess mortality are shown for all locations except Santa Cruz (Bolivia).

## Discussion

This analysis shows the enormous impact that COVID-19 has had on developing countries. The risk of infection observed across developing countries is higher than in high-income nations. Prevalence in developing countries is roughly uniform across age groups, in contrast to the typical pattern in high-income countries where seroprevalence is markedly lower among middle-aged and older adults. The IFR is substantially higher in developing countries than higher-income locations.

We showed that at 20 years of age, the mean IFR in developing countries is 2.7 times higher than that in high-income countries and at age 60 the risk is doubled (Figure 9). At the oldest ages, this discrepancy is reduced, with only a modestly increased risk at age 80. These comparisons are shown in Figure 9 below. This finding does not rely on any specific modelling assumptions such as log-linearity, which is shown by the readily-apparent disparity in figure 7, showing the age-standardized IFR for each individual location compared to the benchmark of high-income countries.

**Figure 9.**
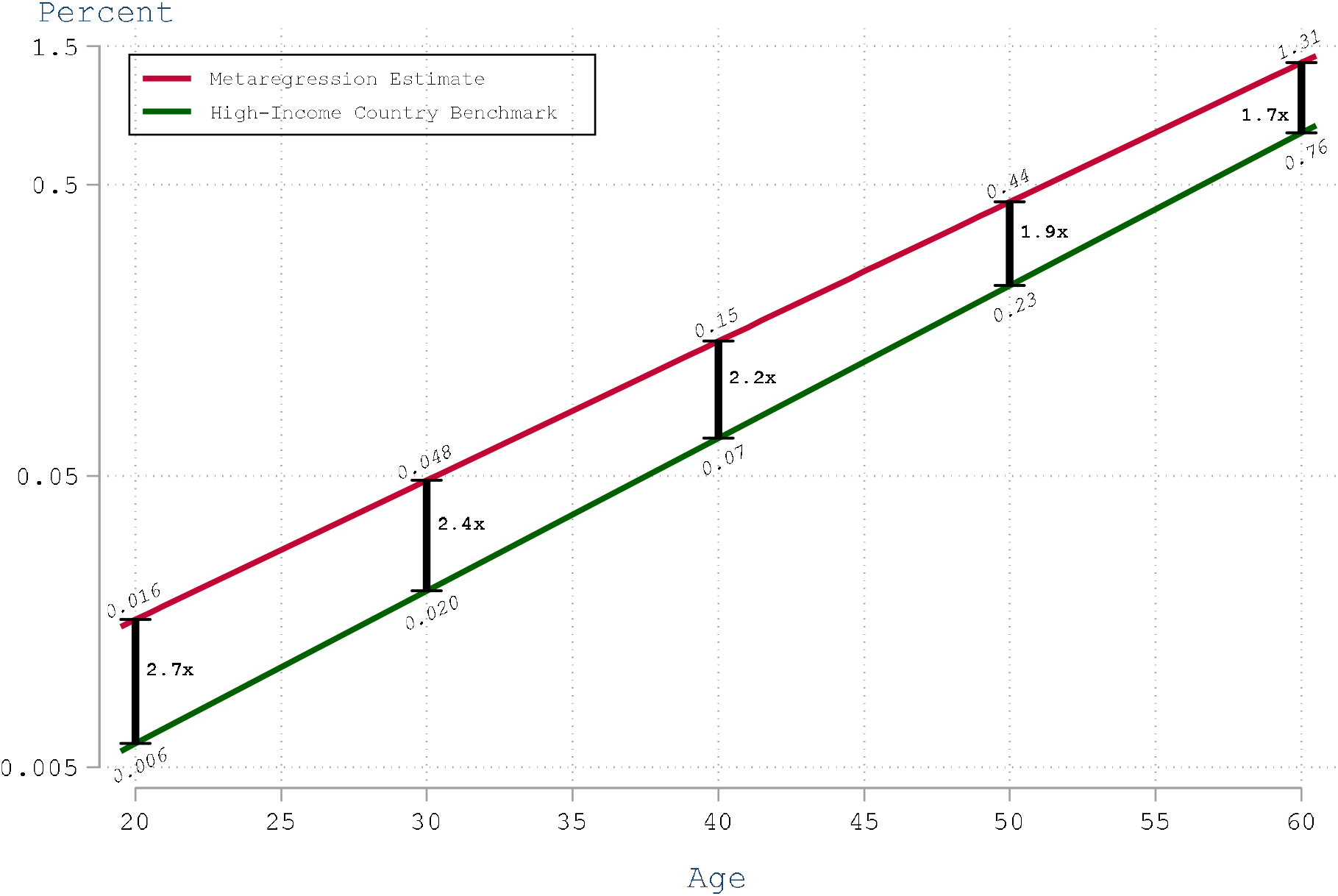
IFR in Developing Countries Compared to High-Income Countries. Comparison of IFRs at different ages for high-income vs developing countries

These results are consistent with the pattern observed for most other communicable diseases. In locations with little ability to work from home, where quarantine is difficult or impossible, where opportunities for physical distancing and access to sanitation are poor, with lower healthcare resources, and where even basic resources such as supplemental oxygen are in short supply, people have fared substantially worse during the pandemic than in high-income settings. Indeed, in low-income settings where fewer hospital beds and health care workers are available, COVID-19 has caused great devastation and an enormous death toll. With a much higher IFR, particularly in younger people, the ultimate burden for developing nations from COVID-19 is likely to be very high.

Another important facet of our results is that seroprevalence was both higher and consistent across age-groups in developing countries--a striking contrast to the typical pattern in high-income countries, where prevalence among older adults was markedly lower than among younger adults (3, 26, 27). Evidently, it is very difficult to insulate elderly people from the virus in a slum or a rural village. This is likely also impacted by the higher proportion of multi-generational families in developing countries (28), a known risk-factor for COVID-19 infection and death (29, 30) For example, seroprevalence in slum neighbourhoods of Mumbai was about four times higher than in non-slum neighbourhoods (31). Our analysis indicates that the relatively uniform prevalence of COVID-19 in developing countries has dramatically increased the number of fatalities in these locations.

Our findings reinforce the conclusions of previous studies that have assessed the IFR of COVID-19 (16, 32). In particular, COVID-19 is dangerous for middle-aged adults, not just the elderly and infirm (3). Our results are also well-aligned with IFR estimates produced for specific locations in developing countries (see supplementary appendices).

Our analysis underscores that incomplete death reporting is a crucial source of apparent differences in COVID-19 death rates. In particular, this is related to the proportion of deaths that are assigned to so-called “garbage codes” (1, 33, 34). These deaths are, by definition, not included in national tallies of the population that has died from COVID-19. In places where death reporting systems are adequate to record deaths, the IFR is on average 10x higher than in places where many deaths are left uncertified.

The divergence between population IFRs for locations is similar whether adjusted for death certification or excess mortality. Adjustment for estimates of excess mortality produced location population IFRs that were consistent with IFRs produced in the age-stratified analysis aside from a few minor outliers. The median of these population IFRs for developing nations, once adjusted for potential undercounting of COVID-19 deaths, was either 0.49% or 0.58%, which was very similar to estimates of IFR from earlier in the pandemic (16, 35).

Excess mortality is a useful metric for adjusting IFR estimates in areas where deaths are well-registered but not well-certified, that is, captured in national vital statistics but without a specific cause of death (4). Nonetheless, caution is warranted in applying national estimates of excess mortality to specific regions within a country, recognizing that death reporting systems may vary markedly with the degree of urbanization and other socioeconomic factors. In the case of Ecuador, for example, the national estimate for the ratio of excess mortality to reported COVID-19 deaths in 2020 was 2.6 (2, 4), whereas that ratio was only 1.01 in the province of Azuay (36).

Moreover, estimates of excess mortality may partly reflect indirect effects of the pandemic on other sources of mortality. On the one hand, non-pharmaceutical interventions may reduce mortality from causes such as vehicle accidents (37). Conversely, mortality may be elevated by impaired access to healthcare for non-infectious diseases such as chronic cardiovascular disease or cancer (38), or by higher burdens of non-COVID infectious diseases such as malaria, tuberculosis, or parasitic infections (39).

Finally, the true burden of COVID-19 may be practically impossible to assess in locations where many deaths are never entered into the national vital statistics system (40). For example, total mortality in Kenya was lower in 2020 than in 2019, but those statistics should certainly not be interpreted as suggesting that Kenya was unscathed by the pandemic (2). Indeed, assessments of Kenya’s vital statistics found that only two-thirds of actual deaths were recorded in the system (40). Such considerations may explain other outliers in our analysis, such as Senegal, which remains far below similar locations even when estimates are adjusted for excess mortality.

A useful example in this case is Ethiopia. Despite national statistics not showing a large increase in deaths in Ethiopia during the pandemic, an epidemiological investigation of burial sites has revealed a huge increase in mortality during this period that is not part of the official reporting of COVID-19 (41).

In the absence of better death reporting, it is challenging to assess the extent to which differences in IFR across locations reflect systematic disparities in healthcare access, socioeconomic status, and other indicators. Nonetheless, such effects have been clearly demonstrated by studies that have assessed distinct socioeconomic groups within specific regions such as Santiago, Chile (10). Moreover, these considerations are almost certainly relevant in interpreting our finding that age-stratified IFR is markedly higher in developing countries compared to high-income countries (42). Indeed, our results underscore the tragedy that a Zambian young adult with COVID-19 would be far more likely to die than a Swiss person of similar age.

Our analysis makes a novel contribution in providing a systematic and comprehensive assessment of the implications of seroreversion, that is, the proportion of people who develop antibodies but whose tests will fall below the limit of detection at a later date. Prior studies have either ignored this issue or have assumed that seroreversion occurs at a fixed geometric rate regardless of the assay used (15, 16). In contrast, we have collated detailed information about the characteristics of all assays used in the serology studies included in our analysis, including data on seroreversion as well as test specificity and sensitivity; that information is fully described in our supplementary appendices. Our analysis clearly indicates that the extent of seroreversion differs in magnitude depending on the assay used. Moreover, accounting for seroreversion and other assay characteristics is crucial for assessing seroprevalence accurately in many of the locations covered by our analysis.

Our analysis makes a very strong case for swifter action on vaccine equity. While countries have largely sought to protect their own populations, there is increasing commitment to ensuring that key populations in low and middle income countries receive vaccines, at a minimum for their front-line health and other personnel. It is widely accepted that failing to control the pandemic across the globe will contribute to the emergence of additional strains of COVID-19, potentially undermining the efficacy of available vaccines (43). Current vaccine distribution efforts are grossly inequitable (44). Current estimates suggest that fewer than 10% of people in low-income countries have received an immunization, while well above 50% of people in high-income countries have had at least one vaccination (45).

As with all research, our study is subject to a number of limitations. Firstly, while we made every effort to capture seroprevalence data, including corresponding with dozens of researchers and public health officials worldwide, it is possible that some studies have been missed. However, it is unlikely that any small number of additional studies would make a material difference to our results.

Our analysis did not incorporate time series data on the evolution of COVID-19 deaths. However, some studies of high-income countries have shown how such data can be useful in refining assessment of IFR to incorporate the stochastic timing of COVID-19 deaths (16, 46). Such analysis should be a priority for future research about IFR in developing countries.

Our work also did not consider non-mortality harms from COVID-19. Recent work has shown that even at younger ages a substantial fraction of infected individuals will have severe, long-lasting adverse effects from COVID-19 (47). Consequently, the impact on the healthcare system and society may be far greater than would be reflected in mortality rates alone. Focusing only on survival rates obscures the large number of deaths that occur from non-COVID-19 when many people are infected (48), SARS-CoV-2’s relatively high fatality rate in comparison to other pathogens and other causes of death (49), and non-mortality harms of COVID-19, such hospitalization from serious disease (47). Future work should address these non-mortality harms, including Long COVID.

In spite of our comprehensive search efforts, no search methodology can be entirely complete. Just before this paper was completed, the WHO released a systematic review of all serology studies conducted on COVID-19 during the pandemic (50). This identified a handful of additional research projects that were not included in this review, however the two systematic reviews did overlap in the vast majority of cases.

Another potentially serious limitation of our analysis is cross-reactivity in serological tests due to malaria. An investigation in Nigeria found that the commonly-used Abbott and Euroimmun serological assays had a false positive rate of 6.1% against pre-pandemic samples due to cross-reactivity with malarial antibodies (51). This would substantially lower specificity of the assay in areas with a high prevalence of past malaria infection, which would have the practical result of increasing seroprevalence estimates and reducing IFR estimates. Thus, it is plausible that in areas with a large burden of malaria, that the IFR we have calculated represents a substantial underestimate.

Finally, our analysis only includes serology studies where specimen collection was completed by the end of February 2021. Consequently, our results do not reflect any potential changes in IFR that may have resulted from more recent advances in COVID-19 care, most notably, the development of novel antiviral medications and dissemination of vaccines. Of course, the IFR could also shift with the spread of new variants of SARS-CoV-2 (52).

## Conclusion

The prevalence and IFR by age of COVID-19 is far higher in developing countries than in high-income countries, reflecting a combination of elevated transmission to middle-aged and older adults as well as limited access to adequate healthcare. These results underscore the critical need to accelerate the provision of vaccine doses to vulnerable populations in developing countries. Moreover, many developing countries require ongoing support to upgrade the quality of their vital statistics systems to facilitate public health decisions and actions, not only for the COVID-19 pandemic but for future global health concerns.

## Supporting information

PRISMA checklist

IFR supplement

## Data Availability

Data and code are available online at https://covid-ifr.github.io/

## Statements of Competing Interests

This work was not externally funded and the authors report no financial or other conflicts of interest.

## Code and Data

All data and code are available publicly online: https://covid-ifr.github.io/

## Acknowledgements

Thanks to Ariel Karlinsky for assistance with death registration and mortality data. Patients and the public were not involved in this research.

## Notes

### Competing Interest Statement

The authors have declared no competing interest.

### Funding Statement

This study was not funded

### Author Declarations

This research required no ethical approval

### Summary of Updates

Updated graphs, some minor referencing issues fixed, and updated supplementary appendices.

## References

1. Vos T, Lim SS, Abbafati C, Abbas KM, Abbasi M, Abbasifard M, et al. Global burden of 369 diseases and injuries in 204 countries and territories, 1990–2019: a systematic analysis for the Global Burden of Disease Study 2019. The Lancet. 2020;396(10258):1204–22.

2. COVID-19 estimate downloads. Seattle: Institute for Health Metrics and Evaluation; 2021. 3.

3. Levin AT, Hanage WP, Owusu-Boaitey N, Cochran KB, Walsh SP, Meyerowitz-Katz G. Assessing the age specificity of infection fatality rates for COVID-19: systematic review, meta-analysis, and public policy implications. European Journal of Epidemiology. 2020;35(12):1123–38.

4. Karlinsky A, Kobak D. Tracking excess mortality across countries during the COVID-19 pandemic with the World Mortality Dataset. eLife. 2021;10:e69336.

5. Ramachandran S, Malani A. All-cause mortality during SARS-CoV-2 Pandemic in India: Nationally-representative estimates independent of official death registry. medRxiv. 2021:2021.07.20.21260577.

6. Gilks CF, Crowley S, Ekpini R, Gove S, Perriens J, Souteyrand Y, et al. The WHO public-health approach to antiretroviral treatment against HIV in resource-limited settings. The Lancet. 2006;368(9534):505–10.

7. Dye C. Global epidemiology of tuberculosis. The Lancet. 2006;367(9514):938–40.

8. Deshmukh Y, Suraweera W, Tumbe C, Bhowmick A, Sharma S, Novosad P, et al. Excess mortality in India from June 2020 to June 2021 during the COVID pandemic: death registration, health facility deaths, and survey data. medRxiv. 2021:2021.07.20.21260872.

9. The true death toll of COVID-19. World Health Organization; 2021.

10. Mena GE, Martinez PP, Mahmud AS, Marquet PA, Buckee CO, Santillana M. Socioeconomic status determines COVID-19 incidence and related mortality in Santiago, Chile. Science. 2021;372(6545):eabg5298.

11. Wetzler HP, Wetzler EA. COVID-19 Excess Deaths in the United States, New York City, and Michigan During April 2020. medRxiv. 2020:2020.04.02.20051532.

12. Modi C, Boehm V, Ferraro S, Stein G, Seljak U. How deadly is COVID-19? A rigorous analysis of excess mortality and age-dependent fatality rates in Italy. medRxiv. 2020:2020.04.15.20067074. 13.

13. Mwananyanda L, Gill CJ, MacLeod W, Kwenda G, Pieciak R, Mupila Z, et al. Covid-19 deaths in Africa: prospective systematic postmortem surveillance study. BMJ. 2021;372:n334.

14. Gill CJ. Latest data from Lusaka morgue analysis shows spike in COVID-19 deaths. The Conversation. 2021.

15. O’Driscoll M, Ribeiro Dos Santos G, Wang L, Cummings DAT, Azman AS, Paireau J, et al. Age-specific mortality and immunity patterns of SARS-CoV-2. Nature. 2021;590(7844):140–5.

16. Brazeau N, Verity R, Jenks S, Fu H, Whittaker C, Winskill P, et al. Report 34: COVID-19 infection fatality ratio: estimates from seroprevalence. Imperial College London; 2020.

17. Chen X, Chen Z, Azman AS, Deng X, Sun R, Zhao Z, et al. Serological evidence of human infection with SARS-CoV-2: a systematic review and meta-analysis. The Lancet Global Health.

18. Meyerowitz-Katz G, Merone L. A systematic review and meta-analysis of published research data on COVID-19 infection-fatality rates. medRxiv. 2020:2020.05.03.20089854.

19. World Economic and Financial Surveys World Economic Outlook Database—WEO Groups and Aggregates Information. 2021.

20. Community Assessment for Public Health Emergency Response Toolkit. CDC; 2019.

21. Population-based age-stratified seroepidemiological investigation protocol for COVID-19 virus infection. World Health Organization; 2020.

22. Gajda M, Kowalska M, Zejda JE. Impact of Two Different Recruitment Procedures (Random vs. Volunteer Selection) on the Results of Seroepidemiological Study (SARS-CoV-2). International Journal of Environmental Research and Public Health. 2021;18(18):9928.

23. Barchuk A, Shirokov D, Sergeeva M, Tursun-zade R, Dudkina O, Tychkova V, et al. Evaluation of the performance of SARS--CoV--2 antibody assays for a longitudinal population-based study of COVID--19 spread in St. Petersburg, Russia. Journal of Medical Virology. 2021;93(10):5846–52.

24. Gelman A, Carpenter B. Bayesian analysis of tests with unknown specificity and sensitivity. medRxiv. 2020:2020.05.22.20108944.

25. Team SD. RStan: the R interface to Stan. R package version 2.19.3. 2020.

26. Stringhini S, Wisniak A, Piumatti G, Azman AS, Lauer SA, Baysson H, et al. Repeated seroprevalence of anti-SARS-CoV-2 IgG antibodies in a population-based sample from Geneva, Switzerland. medRxiv. 2020:2020.05.02.20088898.

27. Pollán M, Pérez-Gómez B, Pastor-Barriuso R, Oteo J, Hernán MA, Pérez-Olmeda M, et al. Prevalence of SARS-CoV-2 in Spain (ENE-COVID): a nationwide, population-based seroepidemiological study. The Lancet. 2020;396(10250):535–44.

28. Living arrangements of older persons around the world. Department of Economic and Social Affairs; 2019.

29. Reyes-Vega MF, Soto-Cabezas MG, Cárdenas F, Martel KS, Valle A, Valverde J, et al. SARS-CoV-2 prevalence associated to low socioeconomic status and overcrowding in an LMIC megacity: A population-based seroepidemiological survey in Lima, Peru. EClinicalMedicine. 2021;34.

30. Ghosh AK, Venkatraman S, Soroka O, Reshetnyak E, Rajan M, An A, et al. Association between overcrowded households, multigenerational households, and COVID-19: a cohort study. Public Health. 2021;198:273–9.

31. Malani A, Shah D, Kang G, Lobo GN, Shastri J, Mohanan M, et al. Seroprevalence of SARS-CoV-2 in slums versus non-slums in Mumbai, India. The Lancet Global Health. 2021;9(2):e110–e1.

32. Verity R, Okell LC, Dorigatti I, Winskill P, Whittaker C, Imai N, et al. Estimates of the severity of coronavirus disease 2019: a model-based analysis. The Lancet Infectious diseases. 2020.

33. Fullman N, Barber RM, Abajobir AA, Abate KH, Abbafati C, Abbas KM, et al. Measuring progress and projecting attainment on the basis of past trends of the health-related Sustainable Development Goals in 188 countries: an analysis from the Global Burden of Disease Study 2016. The Lancet. 2017;390(10100):1423–59.

34. Collaborators GS. Measuring progress from 1990 to 2017 and projecting attainment to 2030 of the health-related Sustainable Development Goals for 195 countries and territories: a systematic analysis for the Global Burden of Disease Study 2017. Lancet (London, England). 2018;392(10159):2091–138.

35. Meyerowitz-Katz G, Merone L. A systematic review and meta-analysis of published research data on COVID-19 infection fatality rates. International Journal of Infectious Diseases. 2020;101:138–48.

36. Cuéllar L, Torres I, Romero-Severson E, Mahesh R, Ortega N, Pungitore S, et al. Excess deaths reveal the true spatial, temporal and demographic impact of COVID-19 on mortality in Ecuador. International Journal of Epidemiology. 2021.

37. Calderon-Anyosa RJC, Kaufman JS. Impact of COVID-19 lockdown policy on homicide, suicide, and motor vehicle deaths in Peru. Preventive Medicine. 2021;143:106331.

38. Meyerowitz-Katz G, Bhatt S, Ratmann O, Brauner JM, Flaxman S, Mishra S, et al. Is the cure really worse than the disease? The health impacts of lockdowns during COVID-19. BMJ Global Health. 2021;6(8):e006653.

39. Buonfrate D, Bisanzio D, Giorli G, Odermatt P, Fürst T, Greenaway C, et al. The Global Prevalence of Strongyloides stercoralis Infection. Pathogens. 2020;9(6):468.

40. Karlinsky A. International Completeness of Death Registration 2015-2019. medRxiv. 2021:2021.08.12.21261978.

41. Ahmed Y. COVID-19 mortality: Twitter; 2021 [Available from: https://twitter.com/yakob_son/status/1450448132047790082?s=20.

42. Demombynes G. COVID-19 Age-Mortality Curves Are Flatter in Developing Countries. World Bank Group. 2021;Public Research Working Paper(9313).

43. COVAX: With a fast-moving pandemic, no one is safe, unless everyone is safe: World Health Organization; 2020 [Available from: https://www.who.int/initiatives/act-accelerator/covax.

44. Keith Collins JH. See How Rich Countries Got to the Front of the Vaccine Line. New York Times. 2021.

45. COVID-19 Data Explorer: Our World In Data; 2021 [Available from: https://ourworldindata.org/explorers/coronavirus-data-explorer.

46. Folkhälsomyndigheten. The infection fatality rate of COVID-19 in Stockholm – Technical report. Sweden: Public Health Agency of Sweden; 2020.

47. Herrera-Esposito D, de los Campos G. Age-specific rate of severe and critical SARS-CoV-2 infections estimated with multi-country seroprevalence studies. medRxiv. 2021:2021.07.29.21261282.

48. Moser W, Fahal MAH, Abualas E, Bedri S, Elsir MT, Mohamed Mfero, et al. Retrospective mortality and prevalence of SARS-CoV-2 antibodies in greater Omdurman, Sudan: a population–based cross–sectional survey. medRxiv. 2021:2021.08.22.21262294.

49. Lapidus N, Paireau J, Levy-Bruhl D, de Lamballerie X, Severi G, Touvier M, et al. Do not neglect SARS-CoV-2 hospitalization and fatality risks in the middle-aged adult population. Infectious Diseases Now. 2021;51(4):380–2.

50. Bergeri I, Whelan M, Ware H, Subissi L, Nardone A, Lewis HC, et al. Global epidemiology of SARS-CoV-2 infection: a systematic review and meta-analysis of standardized population-based seroprevalence studies, Jan 2020-Oct 2021. medRxiv. 2021:2021.12.14.21267791.

51. Steinhardt LC, Ige F, Iriemenam NC, Greby SM, Hamada Y, Uwandu M, et al. Cross-Reactivity of Two SARS-CoV-2 Serological Assays in a Setting Where Malaria Is Endemic. Journal of Clinical Microbiology. 2021;59(7):e00514–21.

52. Classification of Omicron (B.1.1.529): SARS-CoV-2 Variant of Concern. Geneva: World Health Organization; 2021.

53. Buenos Aires city seroprevalence. City of Buenos Aires; 2020.

54. COVID-19 seroprevalence study carried out by the University of the Municipality of Hurlingham.

55. The first phase of the Serovigilance Epidemiological Study concludes. Gobierno Municipal de Santa Cruz; 2020.

56. Saúde presents partial data from the Covid-19 seroepidemiological survey in DF. FEDERAL DISTRICT HEALTH DEPARTMENT; 2020.

57. Silva AAMd, Lima-Neto LG, Azevedo CdMPeSd, Costa LMMd, Bragança MLBM, Filho Akdb, et al. Population-based seroprevalence of SARS-CoV-2 is more than halfway through the herd immunity threshold in the State of Maranhão, Brazil. medRxiv. 2020:2020.08.28.20180463.

58. Research indicates that 12.5% of the Mato Grosso population has already been infected by the coronavirus: Governo de Mato Grosso; 2021 [Available from: http://www.mt.gov.br/-/15990626-pesquisa-aponta-que-12-5-da-populacao-mato-grossense-ja-foi-infectada-pelo-coronavirus.

59. Serological surveys show a drop in antibody levels against Covid-19. Federal University of Latin American Integration: Federal University of Latin American Integration; 2020.

60. Hartwig FP, Vidaletti LP, Barros AJD, Victora GD, Menezes AMB, Mesenburg MA, et al. Combining serological assays and official statistics to describe the trajectory of the COVID-19 pandemic: results from the EPICOVID19-RS study in Rio Grande do Sul (Southern Brazil). medRxiv. 2021:2021.05.21.21257634.

61. . !!! INVALID CITATION !!! {}.

62. Tess BH, Granato CFH, Porto Alves Mcg, Pintao MC, Rizzatti E, Nunes MC, et al. SARS-CoV-2 seroprevalence in the municipality of São Paulo, Brazil, ten weeks after the first reported case. medRxiv. 2020:2020.06.29.20142331.

63. Vial PAaG, Claudia and Icaza, Gloria and Ramirez-Santana, Muriel and Quezada-Gaete, Ruben and Nuñez-Franz, Loreto and Apablaza, Mauricio and Vial, M. Cecilia and Rubilar, Paola and Correa, Juan and Pérez, Claudia and Florea, Andrei and Guzman, Eugenio and Lavin, Maria-Estela and Concha, Paula and Najera-de Ferrari, Manuel and Najera-de Ferrari, Manuel and Aguilera, Ximena,. Seroprevalence, Spatial Distribution, and Social Determinants of SARS-CoV-2 in Three Urban Centers of Chile.. SSRN. 2021.

64. Mercado-Reyes MaM-R, Jeadran N. and Zapata, Silvana and Rodriguez-Barraquer, Isabel and Wiesner, Magdalena and Toloza-Pérez, Yesith Guillermo and Cucunubá, Zulma M. and Hernández-Ortíz, Juan P. and Acosta-Reyes, Jorge and Estupiñan, Maria I. and Galindo, Marisol and Rubio, Vivian V. and Muñoz-Galindo, Lyda and Osorio-Velázquez, Ericson Gabriel and Ibáñez-Pinilla, Edgar A. and Parra Barrera, Eliana L and Bermúdez, Andrea del Pilar and Quinche, Gianni G. and Puerto-Castro, Gloria M. and Villar, Luis A. and Franco-Muñoz, Carlos and Castellanos, Jaime and Navarro-Lechuga, Edgar and Valle, Edna Margarita and Pinto, Nelson and Gore-Saravia, Nancy and OVIEDO Arango, Juan Daniel and Ospina-Martínez, Martha Lucía and Miranda, Maria C. Seroprevalence of Anti-Sars-Cov-2 Antibodies in Colombia, 2020: A Population-Based Study. SSRN. 2021.

65. Alvis Guzman N, De la Hoz Restrepo F, Serrano-Coll H, Gastelbondo B, Mattar S. Using serological studies to assess COVID-19 infection fatality rate in developing countries: A case study from one Colombian department. International Journal of Infectious Diseases. 2021;110:4–5.

66. Acurio-Páez D, Vega B, Orellana D, Charry R, Gómez A, Obimpeh M, et al. Seroprevalence of SARS-CoV-2 Infection and Adherence to Preventive Measures in Cuenca, Ecuador, October 2020, a Cross-Sectional Study. International Journal of Environmental Research and Public Health. 2021;18(9):4657.

67. Informe de Resultados de la Encuesta Nacional de Salud y Nutrición - Continua COVID-19. Encuesta Nacional de Salud y Nutrición; 2020.

68. Sequera Guillermo CA, Samudio Margarita, Vázquez Cynthia, Ocampos Sandra, Galeano Rosa, Von Horoch Marta, Ortega Maria José. Infección por COVID 19: estudio seroepidemiológico de cohorte de base poblacional estratificado por edad en Asunción y Central. Consejo Nacional De Ciencia Y Tecnologia; 2020.

69. Huamaní C, Velásquez L, Montes S, Mayanga-Herrera A, Bernabé-Ortiz A. Population-based seroprevalence of SARS-CoV-2 antibodies in a high-altitude setting in Peru. medRxiv. 2021:2021.01.17.21249990.

70. Álvarez-Antonio C, Meza-Sánchez G, Calampa C, Casanova W, Carey C, Alava F, et al. Seroprevalence of anti-SARS-CoV-2 antibodies in Iquitos, Peru in July and August, 2020: a population-based study. The Lancet Global Health. 2021;9(7):e925–e31.

71. Díaz-Vélez C, Failoc-Rojas VE, Valladares-Garrido MJ, Colchado J, Carrera-Acosta L, Becerra M, et al. SARS-CoV-2 seroprevalence study in Lambayeque, Peru. June–July 2020. PeerJ. 2021;9:e11210.

72. Merkely B, Szabó AJ, Kosztin A, Berényi E, Sebestyén A, Lengyel C, et al. Novel coronavirus epidemic in the Hungarian population, a cross-sectional nationwide survey to support the exit policy in Hungary. Geroscience. 2020;42(4):1063–74.

73. Zejda JE, Brożek GM, Kowalska M, Barański K, Kaleta-Pilarska A, Nowakowski A, et al. Seroprevalence of Anti-SARS-CoV-2 Antibodies in a Random Sample of Inhabitants of the Katowice Region, Poland. International Journal of Environmental Research and Public Health. 2021;18(6):3188.

74. Alemu BN, Addissie A, Mamo G, Deyessa N, Abebe T, Abagero A, et al. Sero-prevalence of anti-SARS-CoV-2 Antibodies in Addis Ababa, Ethiopia. bioRxiv. 2020:2020.10.13.337287.

75. Shaweno T, Abdulhamid I, Bezabih L, Teshome D, Derese B, Tafesse H, et al. Seroprevalence of SARS-CoV-2 antibody among individuals aged above 15 years and residing in congregate settings in Dire Dawa city administration, Ethiopia. Tropical Medicine and Health. 2021;49(1):55.

76. Ngere I, Dawa J, Hunsperger E, Otieno N, Masika M, Amoth P, et al. High seroprevalence of SARS-CoV-2 but low infection fatality ratio eight months after introduction in Nairobi, Kenya. Int J Infect Dis. 2021;112:25–34.

77. Inquérito Sero-epidemiológico de SARS-CoV-2 na Cidade de Maxixe e Vila de Massinga (InCOVID 2020) - Resultados Preliminares -. República de Moçambique: Ministério da Saúde; 2020.

78. Talla CaL, Cheikh and Roka, Jerlie Loko and Barry, Aliou and Ndiaye, Seynabou and Diarra, Maryam and Faye, Oumar and Dia, Moussa and Tall, Adama and Ndiaye, Oumar and Faye, Rokhaya and Mbow, Adji Astou and Diouf, Babacar and Diallo, Jean Pierre and Ndiaye, Mamadou and Woudenberg, Tom and White, Michael and Ting, Jim Y. and Diagne, Cheikh Tidiane and Pasi, Omer and Diop, Boly and Sall, Amadou and Vigan-Womas, Inès and Faye, Ousmane. Seroprevalence of Anti-SARS-CoV-2 Antibodies in Senegal: A National Population-Based Cross-Sectional Survey, between October and November 2020. SSRN. 2020.

79. Mutevedzi PC, Kawonga M, Kwatra G, Moultrie A, Baillie V, Mabena N, et al. Estimated SARS-CoV-2 infection rate and fatality risk in Gauteng Province, South Africa: a population-based seroepidemiological survey. International Journal of Epidemiology. 2021.

80. Sue Aitken JY, Tamika Fellows, Tetelo Makamadi, Sarah Magni, Renay Weiner, Cherie Cawood, Anne von Gottberg, Jinal N. Bhiman, Sibongile Walaza, Jocelyn Moyes, Meredith McMorrow, Stefano Tempia, Neil Martinson, Limakatso Lebina, Cheryl Cohen, Nicole Wolter. COVID-19 SEROPREVALENCE DURING THE SECOND WAVE OF THE PANDEMIC IN THREE DISTRICTS OF SOUTH AFRICA - PRELIMINARY FINDINGS. NATIONAL INSTITUTE FOR COMMUNICABLE DISEASES; 2021.

81. Mulenga LB, Hines JZ, Fwoloshi S, Chirwa L, Siwingwa M, Yingst S, et al. Prevalence of SARS-CoV-2 in six districts in Zambia in July, 2020: a cross-sectional cluster sample survey. The Lancet Global Health. 2021;9(6):e773–e81.

82. Khalagi K, Gharibzadeh S, Khalili D, Mansournia MA, Samiee SM, Aghamohamadi S, et al. Prevalence of COVID-19 in Iran: Results of the first survey of the Iranian COVID-19 Serological Surveillance program. medRxiv. 2021:2021.03.12.21253442.

83. Bellizzi S, Alsawalha L, Sheikh Ali S, Sharkas G, Muthu N, Ghazo M, et al. A three-phase population based sero-epidemiological study: Assessing the trend in prevalence of SARS-CoV-2 during COVID-19 pandemic in Jordan. One Health. 2021;13:100292.

84. Al-Abri SS, Al-Wahaibi A, Al-Kindi H, Kurup PJ, Al-Maqbali A, Al-Mayahi Z, et al. Seroprevalence of SARS-CoV-2 antibodies in the general population of Oman: results from four successive nationwide sero-epidemiological surveys. International Journal of Infectious Diseases. 2021;112:269–77.

85. Sharma N, Sharma P, Basu S, Saxena S, Chawla R, Dushyant K, et al. The seroprevalence of severe acute respiratory syndrome coronavirus 2 in Delhi, India: a repeated population-based seroepidemiological study. Transactions of The Royal Society of Tropical Medicine and Hygiene. 2021.

86. Mohanan M, Malani A, Krishnan K, Acharya A. Prevalence of COVID-19 In Rural Versus Urban Areas in a Low-Income Country: Findings from a State-Wide Study in Karnataka, India. medRxiv. 2020:2020.11.02.20224782.

87. Khan SMS, Qurieshi MA, Haq I, Majid S, Ahmad J, Ayub T, et al. Seroprevalence of SARS-CoV-2-specific IgG antibodies in Kashmir, India, 7 months after the first reported local COVID-19 case: results of a population-based seroprevalence survey from October to November 2020. BMJ Open. 2021;11(9):e053791.

88. Saple P, Gosavi S, Pawar T, Chaudhari G, Mahale H, Deshmukh P, et al. Seroprevalence of anti-SARS-CoV-2 of IgG antibody by ELISA: Community-based, cross-sectional study from urban area of Malegaon, Maharashtra. J Family Med Prim Care. 2021;10(3):1453–8.

89. Banerjee A, Gaikwad B, Desale A, Jadhav SL, Rathod H, Srivastava K. Severe acute respiratory syndrome-coronavirus-2 seroprevalence study in Pimpri-Chinchwad, Maharashtra, India coinciding with falling trend - Do the results suggest imminent herd immunity? Indian journal of public health. 2021;65(3):256–60.

90. Kshatri JS, Bhattacharya D, Praharaj I, Mansingh A, Parai D, Kanungo S, et al. Seroprevalence of SARS-CoV-2 in Bhubaneswar, India: findings from three rounds of community surveys. Epidemiology and infection. 2021;149:e139.

91. Kar S SS, Murali S, Dhodapkar R, Joseph N, Aggarwal R. Prevalence and Time Trend of SARS-CoV-2 Infection in Puducherry, India. Emerg Infectious Diseases. 2021;27(2):666–9.

92. Malani A, Ramachandran S, Tandel V, Parasa R, Imad S, Sudharshini S, et al. SARS-CoV-2 Seroprevalence in Tamil Nadu in October-November 2020. medRxiv. 2021:2021.02.03.21250949.

93. Selvaraju S KM, Thangaraj J, Bhatnagar T, Saravanakumar V, Kumar C, et al. Population-Based Serosurvey for Severe Acute Respiratory Syndrome Coronavirus 2 Transmission, Chennai, India. Emerg Infectious Diseases. 2021;27(2):586–9.

94. Satpati P, Sarangi S, Gantait K, Endow S, Mandal N, Panchanan K, et al. Sero-surveillance (IgG) of SARS-CoV-2 among Asymptomatic General population of Paschim Medinipur, West Bengal, India. medRxiv. 2020:2020.09.12.20193219.

95. ENHANCED SURVEILLANCE ON SERO-PREVALENCE OF SARS-COV-2 IN GENERAL POPULATION Government of Nepal; 2020.

96. Haq M, Rehman A, Ahmad J, Zafar U, Ahmed S, Khan MA, et al. SARS-CoV-2: big seroprevalence data from Pakistan—is herd immunity at hand? Infection. 2021;49(5):983–8.

97. Li Z, Guan X, Mao N, Luo H, Qin Y, He N, et al. Antibody seroprevalence in the epicenter Wuhan, Hubei, and six selected provinces after containment of the first epidemic wave of COVID-19 in China. The Lancet Regional Health – Western Pacific. 2021;8.

