## Supplementary Appendices for "Assessing the Burden of COVID-19 in Developing Countries: Systematic Review, Meta-Analysis, and Public Policy Implications"

Contents:

1. Data collection and methodology
   a. Systematic review methodology
   b. Death data
   c. Full inclusion and exclusion criteria
   d. Covariates
2. Statistical methodology
   a. Bayesian model for estimating seroprevalence and IFR
   b. Seroconversion and seroreversion
3. Additional results and figures
   a. Age-specific IFRs
   b. Metaregression results (additional)
   c. Population IFR
   d. Risk of bias ratings
   e. Covariate examination
   f. Seroprevalence comparison to high-income countries
   g. IFRs from included studies
   h. Out-of-sample analysis
   i. Excess mortality for age-specific IFR
   j. PRISMA flow diagram
1. Data Collection and Methodology
a. **Systematic Review Methodology**

**Search Procedure**

We searched Serotracker using “[h]ousehold and community samples” and “[p]ersons living in slums*” in the “[d]emographics” field. We also searched MedRxiv, PubMed, Google Scholar, Biorxiv, SSRN, Twitter, and the Pan American Health Organization database using the pre-specified search term “COVID-19 seroprevalence”. Then we cross-checked with recently published systematic reviews of worldwide seroprevalence (1-3), while identifying further studies by searching the grey literature and government websites where appropriate. This included searching using the term “COVID-19 seroprevalence” on Google in languages on Google Translate such as Portuguese, Spanish, English, French, German, and Italian, with an additional search for "inquérito sorológico, COVID-19". Duplicates were reviewed by authors on Google sheets and resolved independently.

We completed searches on October 22^nd^, 2020, and at least monthly afterwards until July 14, 2021. We also performed searches monthly until September 22, 2021 during initial drafting of our paper. After our completing initial draft, we performed additional searches monthly up to December 17, 2021, which represented the final cut-off date for studies included in our analysis. Only studies with results from an official source were included, such as a published paper, pre-print, presentation by government officials, or the website of the institution that performed the study. If a press report or another unofficial source was found, we performed more detailed searches using information from the unofficial source in order to find a matching official source. Study authors were contacted by email or Twitter for further information, when needed. We also ran detailed searches after September 22, 2021 on older studies for which preliminary results were found by September 22, but for which updates were posted after September 22. We include links to the studies at each location in our GitHub repository under appendix material.

Searches were conducted by one member of the team and then repeated to ensure consistency by another. Data were similarly extracted by one member then cross-checked by another. No data collection was automated. This process was recorded by the team working across regions in Google sheets. Data collection is more fully described below but included extracting seroprevalence information from included studies by age where available, as well as death data specific to COVID-19 from each country/region with valid seroprevalence information. Where serology data was not evident in publicly available reports, we reached out to researchers and public health officials using both email and social media. For death data we largely relied on publicly available official reports.

Studies were reviewed by two authors and screened for inclusion. Disagreements were resolved through discussion between all authors at weekly meetings and via email. Where essential data were missing despite efforts to access them, we excluded the study from our synthesis, as noted in “*Full Inclusion and Exclusion Criteria*”. Our aim was to provide the most robust estimate of age-specific IFR in developing countries, and thus we considered it inappropriate to rely on potentially flawed assumptions regarding these studies in our analysis.

**Study Bias**

We included only those studies that exhibited sufficient quality from which to derive an estimate of IFR in our primary analysis, although we included all estimates of age-stratified serology in our secondary analysis. Given that many of these were from public rather than academic sources, the quality varied widely; however, where possible we contacted the authors to reduce the risk of including biased estimates. All included surveys had an adequate sampling frame and used probabilistic techniques to sample participants, as discussed in “*Full Inclusion and Exclusion Criteria*”.

**Bias Across Studies**

In this context, publication bias in which studies exhibiting certain findings are more likely (or not) to be published, is very unlikely to have an impact, as studies with both high and low seroprevalence estimates are of interest to the scientific literature. Consistent with this, in prior work we found no evidence of publication bias for seroprevalence studies from high-income countries (4). However, to mitigate against the possibility of publication bias influencing our results, we included lengthy searches of grey literature, following up on media reports of seroprevalence studies to ensure that every age-stratified that we were able to identify was in our metasynthesis.

This review is also, to an extent, an examination of uncertainty. Given the numerous sources of data and uncertainty therein that go into making even a single IFR estimate, quality of the literature is a relatively small part of the overall certainty. Many of the included studies were conducted in areas where death reporting was a major source of uncertainty – if only 1 in 10 COVID-19 deaths were recorded as such, the impact of poor study quality is much smaller than the inadequacies of the reporting systems, as discussed further in “*Death Data*” and “*Out-of-Sample Analysis*”. Moreover, this is not a traditional systematic appraisal with a single output concerning a clinical question.

Therefore, we did not formally assess confidence in the estimates, but did produce statistical measures examining the likely range that IFRs may hold given various assumptions about how developing countries managed the pandemic. We also informally assess studies for risk of bias, as discussed in ”*Risk of Bias Ratings*”. And in “*Out-of-Sample Analysis*” we present additional evidence that methodological differences between our included studies may not strongly bias our IFR estimates.

b. **Death Data**

Building on our prior work (4), we assess the length of the lag between the midpoint of serology sampling and the time at which COVID-19 deaths were reported. The time interval between symptom onset and death had an interquartile range (IQR) of:

- 7 to 22 days for Argentina (5)

- 9 to 24 days for Colombia (6)

- 10 to 26 days for the Brazilian states of Espírito Santo (7) and Parana (8)

These intervals largely agree with IQRs reported for the USA as:

- 9 to 24 days for ages 18-64

- 7 to 19 days for ages >64 (4)

The IQR for the interval between death and official reporting for the USA was 2 to 19 days (4). This largely matches the interval ranges for Argentina (5), Colombia (6), and Paraguay (9) before March 2021 when included seroprevalence studies stopped collecting samples (see “*Full Inclusion and Exclusion Criteria*”), as shown below:

**Figure A1 – Death Reporting Lags**

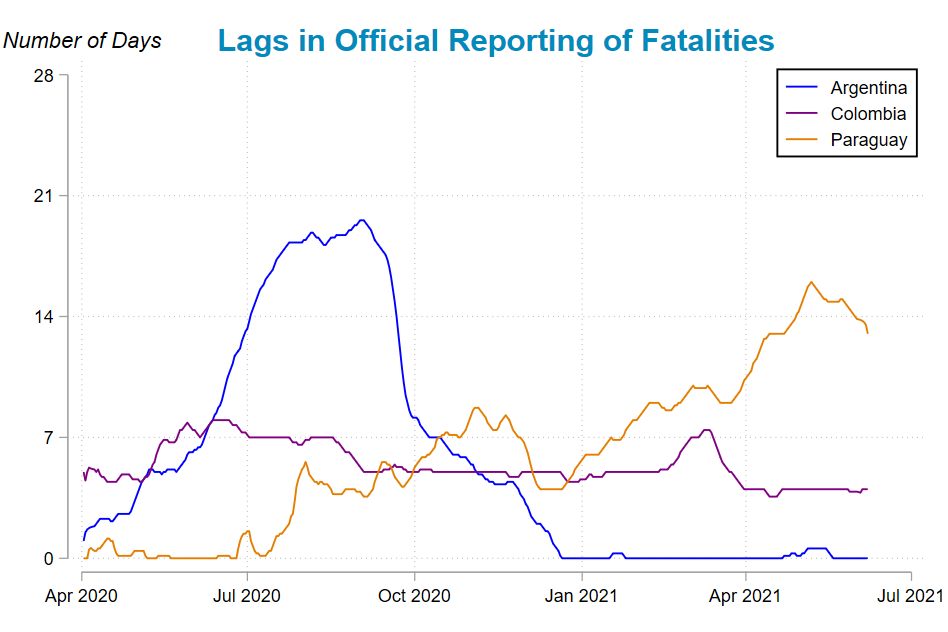

Argentina, Colombia, and Paraguay may be outliers with respect to the systematic collection and publication of vital statistics during the pandemic (10); so other developing countries may have substantially longer reporting lags that may not be documented in the absence of detailed case data. The time interval between symptom onset and official death reporting thus appears roughly similar in developing countries as in our previous analysis of high-income countries such as the USA (4).

Therefore, as with our previous work (4), we extracted death information four weeks after the midpoint of the study in question for developing countries where we rely on official death reports issued in “real time.” However, for some countries it was possible to extract real-time death data, i.e. the dates on which people passed away as well as the date those deaths were reported. For these cases we used a lag of two weeks from the midpoint instead of four weeks, as we did not need to account for reporting lag. These lags reflect the 95^th^ percentiles of the approximately:

- 2-week interval between symptom onset and seropositivity,

- 4-week interval between symptom onset and death,

- 2-week interval between death and official reporting.

There is also the question of what is the most appropriate death data to use. In most countries there are two sets of COVID-19 deaths: confirmed or suspected. In some countries the government will also present a third tally of deaths, which is modelled using excess mortality statistics or similar. Confirmed COVID-19 deaths may under-estimate the total number of COVID-19 deaths due to insufficient testing (11-14). This may be detected by comparing reported COVID-19 deaths with excess deaths, as reflected in the Peruvian government increasing their tally of reported deaths in a manner that better approximated total excess deaths (10, 15). Nepal’s government also later substantially increased their reported tally of COVID-19 deaths (16).

Problems with death reporting and how this can be included in a review of IFR is well-illustrated by Mexico. Mexico has one of the largest tolls from COVID-19 of any country in the world, however the death reporting system for the country has notable gaps. When looking at the raw data on individuals, 90% of those who died did not have a date of death entered into the publicly available data. Previous research also demonstrated that large numbers of people who died from COVID-19 in Mexico failed to access a test and thus are not included in the country’s mortality statistics (14). This means that the reported death data available for Mexico is not sufficient to derive a high-quality COVID-19-related IFR. We therefore instead used an alternative official source in Mexico that accounted for this COVID-19 death under-estimation (17).

So for the purposes of the primary analysis, we included the confirmed + suspected death figures where available instead of only confirmed deaths, as confirmed + suspected is the more robust estimate of reported COVID-19 deaths in developing countries. Death data were extracted from national datasets in each country where possible, with alternative sources noted where applicable. Where death data were not immediately available, we contacted the national or local authority through email or social media. We also attempted to confirm death data using the most robust source, and in most cases took the estimate directly from the relevant health authority rather than data aggregation websites. We include our informal assessment of risk of COVID-19 death under-estimation in our GitHub repository under appendix material (see “*Risk of Bias Ratings*”). This is assessment is based on percentage of deaths well-certified in the past decade (18), and on comparison of reported COVID-19 deaths to excess deaths.

c. **Full Inclusion and Exclusion Criteria**

We included only studies that met both of the following conditions:

1. Report seroprevalence from a representative sample in developing countries, meaning: random selection of participants from a sample frame representative of the general population, such as household sampling, or sampling >50% of the general population by census (19), conducted in countries classified by the International Monetary Fund as “Emerging and Developing Economies” (20).
2. Available online and accessible in English, or via translatable text if not in English.

For countries with no reported age-stratified seroprevalence, but sufficient information to otherwise calculate age-stratified IFRs, we calculated these IFRs assuming equal seroprevalence across age-groups instead of excluding the study. When total sample size and age-specific seroprevalence were known, but age-specific sample sizes were not precisely reported, age-specific sample size was imputed based on the age-distribution of the general population.

We excluded:

1. Convenience samples (3), including those utilizing residual sera from clinical specimens and blood donors (see the “*Blood Donors and Residual Sera*” section below), dialysis centres, healthcare workers, and actively recruited participants constituting less than 50% of the total population sampled (3, 4, 21).
2. Studies sampling a high-income country, as classified by the International Monetary Fund (20), or a wealthy micronation such as Andorra or Monaco.
3. Studies in which sampling extended after February 2021, to help mitigate the risk of seroreversion on longer timeframes (see “*Seroconversion and Seroreversion*”).
4. If gender ratios were reported, then <35% of the sample reported as male or <35% reported as female, without cited evidence that the study’s gender ratio matched the general population.
5. Studies that used the Wondfo serology test. This test is unreliable, as discussed in “*Seroconversion and Seroreversion*”, with some Wondfo batches having very low sensitivity and others more reasonable values, making it difficult to adjust for test sensitivity.
6. Studies in which the total number of individuals tested was unknown or seroprevalence was not reported.
7. Studies using serology tests with insufficient data for estimating sensitivity and specificity from a known number of tested samples.
8. IFR estimate excluded if: A) the sampling start-week or end-week was not known to allow for accurate determination of the corresponding number of COVID-19 deaths, B) test-adjusted population-wide seroprevalence overlapped with 0%, or C) samples were taken during an accelerating outbreak in which reported COVID-19 deaths increased by a factor of three or more from the midpoint date of sampling to 4 weeks later (4).
9. Seroprevalence estimate excluded if both of the following conditions were met: A) IFR estimate was excluded for other reasons listed above, and B) the study overlapped geographically with another included study. This geographical exclusion avoided oversampling the same location (4). IFR estimates that met condition B but not condition A are discussed in the out-of-sample analysis below.

We excluded from the body of the paper studies meeting at least one of the conditions below:

1. IFR estimate for a location that geographically overlapped with an included study, and thus inclusion of both estimates risked oversampling the same location. The “*Out-of-Sample Analysis*” includes IFR estimates for these additional locations.
2. Zero COVID-19 deaths reported for the sampled location, making the calculated IFR non-robust (22). So IFR estimates were not calculated for these locations. Our GitHub repository includes seroprevalence estimates for these out-of-sample locations under appendix material.
3. The total population from which the sample was drawn was less than 20,000, which may not reflect the wider population of the region. IFR estimates were thus not calculated for these locations. Seroprevalence estimates for these out-of-sample locations are included in our GitHub repository under appendix material.

**Blood Donors and Residual Sera**

Blood donor studies are widely used as blood donors are a convenient population from which to draw a population estimate – donors already have blood taken, can be tested easily, and tend to include people from a relatively wide area (23). However, as has been noted in research prior to the pandemic, donors are a highly selected sample and donor studies often have a large bias in terms of estimates of seroprevalence for other diseases (24) (25). Moreover, in many areas, particularly at initial stages of the pandemic, donating blood was one of few methods available to access a serological test. It is unclear which direction this bias generally runs, especially considering the dynamics of a novel pathogen in the community (26).

Residual sera studies examine blood samples taken initially for other reasons. These samples have an obvious bias in that they are representative of people going to have blood taken for reasons other than SARS-CoV-2 tests, a group that may not be representative of the general population (27). Though there may be methods for using residual sera samples to derive estimates of population infection rates, we excluded studies of residual sera and of blood donors from our main analysis to mitigate this additional source of bias. Convenience sampling of such populations may be sufficient for other purposes, but probabilistic selection from a representative sample frame better facilitates accurate estimation of population-wide infection rates (3, 28).

d. **Covariates**

We extracted data from the most recent year prior to the pandemic, which in most cases was 2019 or earlier. For some estimates such as workforce, we relied on the best available data, some of which was several years old for some countries.

The covariates are:

1. GDP per capita
2. Healthcare spending
3. GNI per capita
4. Hospital beds per capita
5. Life expectancy at birth
6. Healthy life expectancy at age 60
7. Global health security index
8. Skilled healthcare workers per capita
9. Universal health coverage index
10. % of deaths well-certified (18)

Briefly, these covariates were chosen because they either relate to the expected quality of the health system itself (i.e. doctors/nurses per population) or to how likely a country was to be accurately recording the burden of COVID-19 (WHO indicators, human development index). We also included the ratio of life expectancy between age 60 and 20 to account for the potential for survivorship bias – if there was a significant element of survivorship bias in the countries examined, we would expect the ratio to be higher as more elderly people survived longer periods in places with higher mortality in youth.

2. Statistical Methodology
   a. **Bayesian Model for Estimating Seroprevalence and IFR**

### **Model for COVID-19 infections**

Let $R_{l,A}^{\star}$ be the number of individuals who tested seropositive in age group *A* at location $l$, and $n_{l,A}$ give the number of individuals tested in that age group for this location. We model the number of individuals with a positive serology test in the study as

$R_{l,A}^{\star}\sim\text{Binomial}\left( n_{l,A},p_{l,A} \right), \text{ }\text{where}\text{ }$	(1)

$p_{l,A,j}=\text{sens}_{t_{l}}\pi_{l,A}+\left( 1-\text{spec}_{t_{l}} \right)\left( 1-\pi_{l,A} \right).$	(2)

To account for the error rates of the test, the test positivity probability, $p_{l,A}$, is defined as a function of test sensitivity ($\text{sens}_{t_{l}}$), test specificity ($\text{spec}_{t_{l}}$), and the true seroprevalence ($\pi_{l,A}$) for the associated location and age group at the time of the study. For many studies, we did not have seropositivity by age, in which case *A* represented all ages.

To account for uncertainty in the test characteristics, we model the lab validation data directly. Let *n*_sens_*_,t_* denote the number of positive specimens tested with test *t*, and *x*_sens_*_,t_* the number of positive specimens that correctly tested positive. Similarly, let *n*_spec_*_,t_* and *x*_spec_*_,t_* denote the number of negative specimens tested and the number of negative specimens that correctly tested negative with test *t*, respectively. We model these quantities as follows:

| *x*sens*_,t_* ∼ Binomial(*n*sens*_,t_,* sens*_t_*) | (3) |
| --- | --- |
| *x*spec*_,t_* ∼ Binomial(*n*spec*_,t_,* spec*_t_*)*.* | (4) |

### **Model for COVID-19 deaths**

Let $D_{l,A}^{\star}$ give the number of recorded COVID-19 deaths, for age group *A* at location $l$. Note that if only a single death record is available, then *A* represents the entire age range. We model the recorded COVID-19 deaths as

$D_{l,A}^{\star}\sim\text{Poisson}\left( N_{l,A}\times\pi_{l,A}\times\text{IFR}_{l,A} \right)$	(5)

where $N_{l,A}$ gives the number of individuals at location $l$ in age group *A*. Then $N_{l,A}\times\pi_{l,A}$ gives the expected number of infected individuals, and $\text{IFR}_{l,A}$ is the infection fatality rate for location $l$ and age group *A*, representing the probability an individual dies from COVID-19, given the individual had COVID-19. Note, the Poisson distribution reflects the relative rarity of a COVID-19 fatality relative to the entire population.

### **Accounting for data collected in varying age bins**

Notice that the models above for deaths and infections in (1) and (5) are functions of prevalence and IFR, respectively, defined on discrete age bins. However, the discrete age bins are not necessarily the same for the death data and the seroprevalence studies. The following adjustments were made to match serology and death age bins:

- **Death bins nested within a serology bin:** We aggregate deaths for each location to match the respective serology age bins to avoid placing assumptions about the variability of prevalence across ages within a single serology age bin.
- **Serology bins nested within a death bin:** The average seroprevalence for the death age bin is calculated as an average of the serology age bins, weighted by the percent of the population in each age bin.
- **Bin endpoints slightly off:** When age bins were within one or two years of matching, serology age bins were adjusted to match the corresponding death age bins.

All modifications to age bins are documented in a spreadsheet in the data folder.

#### **Population Age Distribution**

Let $f_{l}\left( a \right)$ denote the number of individuals of age *a* at location $l$ for *a* ∈ {0*,*1*,...,*84+}. Note, if population age structure is only available in 5 year age bins, then define

$$f_{l}\left( a \right)=\sum_{b\in\{0,5,\ldots,80\}} \frac{f_{l}\left( \left[ b,b+5 \right) \right)}{5}I_{\left[ b,b+5 \right)}\left( a \right)$$

where $f_{l}\left( \left[ b,b+5 \right) \right)$ is the proportion of the population ages $\left[ b,b+5 \right)$.

In cases where the location specific age structure is only available in large bins, but the national age structure is available in 5 year age bins, we leverage the national age structure to inform the location specific age structure as follows. Let *A* denote an interval the location specific age structure is available for (e.g., [0*,*18)). If *f*(*A*) is the proportion of the population at location $l$ with an age in *A* and *f_n_*(*a*) is the proportion of the population aged *a* at the national level, then we estimate $f_{l}\left( a \right)$, the proportion at location $l$ that is age *a*, as

$f_{l}\left( a \right)=f\left( A \right)\frac{f_{n}\left( a \right)}{\sum_{b\in A\cap N} f_{n}\left( b \right)}.$	(6)

Essentially, we rescale *f_n_*(*a*) such that the total mass in *A* matches the observed total mass in *A* at location $l$, *f*(*A*). Since we model seroprevalence as constant past age 85, we let $f_{l}\left( 85 \right)$ represent the proportion of the population aged 85 or older, rather than just the proportion aged 85.

#### **Calculating Average Seroprevalence within a Death Age Bin**

Define the population age density for age bin *A* as

$f_{l,A}\left( a \right)=\frac{f_{l}\left( a \right)}{\sum_{b\in A\cap N} f_{l}\left( b \right)}, a\in\{0,1,\ldots,84+\}$	(7)

in order to truncate $f_{l}\left( a \right)$ to age bin *A*.

The prevalence for age bin $B=\cup_{A\in\mathcal{A}}A$ is then defined

$\pi_{l,B}=\sum_{A\in\mathcal{A}} \left[ \pi_{l,A}\sum_{b\in A\cap N} f_{l,A}\left( b \right) \right].$	(8)

For the locations where we only have serology study information with no corresponding fatality data, the proportion of all study participants that were in a given age bin was assumed representative of the proportion of the population in each age bin since the studies were designed to have representative samples. For the locations with both serology and fatality data, population data were recorded in the Population Distributions tabs with citations.

### **Priors**

For the seroprevalence parameters, $\pi_{l,A}$, we used independent, weakly informative priors:

$\pi_{l,A}\sim\text{Beta}\left( 2,6 \right)\quad\text{for all }l,A.$	(9)

We also used independent priors for the test sensitivities and specificities. Because there are infinitely many combinations of prevalence, sensitivity, and specificity, that can result in the same test positivity rate, similar to Gelman and Carpenter (2020), we used informative priors to avoid a multimodal posterior. For each test assay *t*, the priors on the sensitivity and specificity were

| sens*_t_* ∼ Beta(10*,*1) | (10) |
| --- | --- |
| spec*_t_* ∼ Beta(50*,*1)*.* | (11) |

To further narrow the seroprevalence, sensitivity, and specificity combinations, we used independent, mildly informative priors for each IFR parameter based on expert knowledge. IFR for COVID-19 is known to increase with age. We also expect IFR to be more extreme (smaller than average or larger than average) when the age bin is small. For example, we would expect an age bin from 20-80 to look similar to the country average, but we would expect an age bin from 70-80 to be much higher than the country average. To formulate a prior that reflects these characteristics, we modeled

$\text{IFR}_{l,A}\sim\text{Beta}\left( 1,\text{IFR}_{l,A}^{\text{prior}} \right)$	(12)

where

$\text{IFR}_{l,A}^{\text{prior}}=30-20\left[ \frac{U_{l,A}-50}{50}\left( 1-\frac{U_{l,A}-L_{l,A}}{100} \right) \right].$	(13)

The lower and upper bounds of age bin *A* at location $l$ are given by $L_{l,A}$ and $U_{l,A}$, respectively. For open

ended upper ages, we set $U_{l,A}=100$. As an example, $\text{IFR}_{l,\left[ 0,100 \right)}^{\text{prior}}=30$, while $\text{IFR}_{l,\left[ 80,100 \right)}^{\text{prior}}=14$, allowing for larger IFR estimates when focusing on the older individuals.

### **Model Implementation**

The model was implemented in version 4.0.2 of the programming language R, and posterior samples were obtained via the software package Stan (version 2.21.1). We ran three chains for 10,000 iterations, where the first 5,000 iterations were discarded as warm-up samples. All parameters had an effective sample size greater than 1,200. Additionally, the $\hat{R}$ value was within 0.0016 of 1 for each parameter, suggesting convergence. Examination of traceplots also suggested convergence.

Out-of-sample observation were run as a separate model, so information on test sensitivity and specificity was not pooled between in-sample observations and out-of-sample observations. Traceplots, effective sample size (minimum of 2100), and $\hat{R}$ values (within 0.0029 of 1) suggested convergence of the out-of-sample model as well.

We compared plugin estimates for parameters to the posterior distribution for each parameter to check model fit. In each case there was good agreement, or the Bayesian estimate was superior. For example, in Figure 1, we compare the posterior distribution of the sensitivity and specificity estimates to the raw estimate. In most cases, the middle 50% of the posterior distribution contains the raw estimate. However, for test kit ID 11 (Qingdao Hightop Biotech IgM/IgG Duo), the raw estimate of specificity is outside the range of the posterior draws. In the case of this test, it was used in locations with extremely low prevalence, such that the Gladen-Rogan (29) adjustment results in a negative estimate of prevalence (meaning the expected number of false positives is greater than the number that tested positive.) Since this is unreasonable, the Bayesian model raises the specificity estimate, lowering the expected number of false positives.

**Figure A2 – Sensitivity and Specificity Posterior Distributions**

Boxplots showing the posterior distribution of the (a) sensitivity and (b) specificity for each test kit. Raw sensitivity/specificity based on the lab validation data is shown by a red point for each test kit.

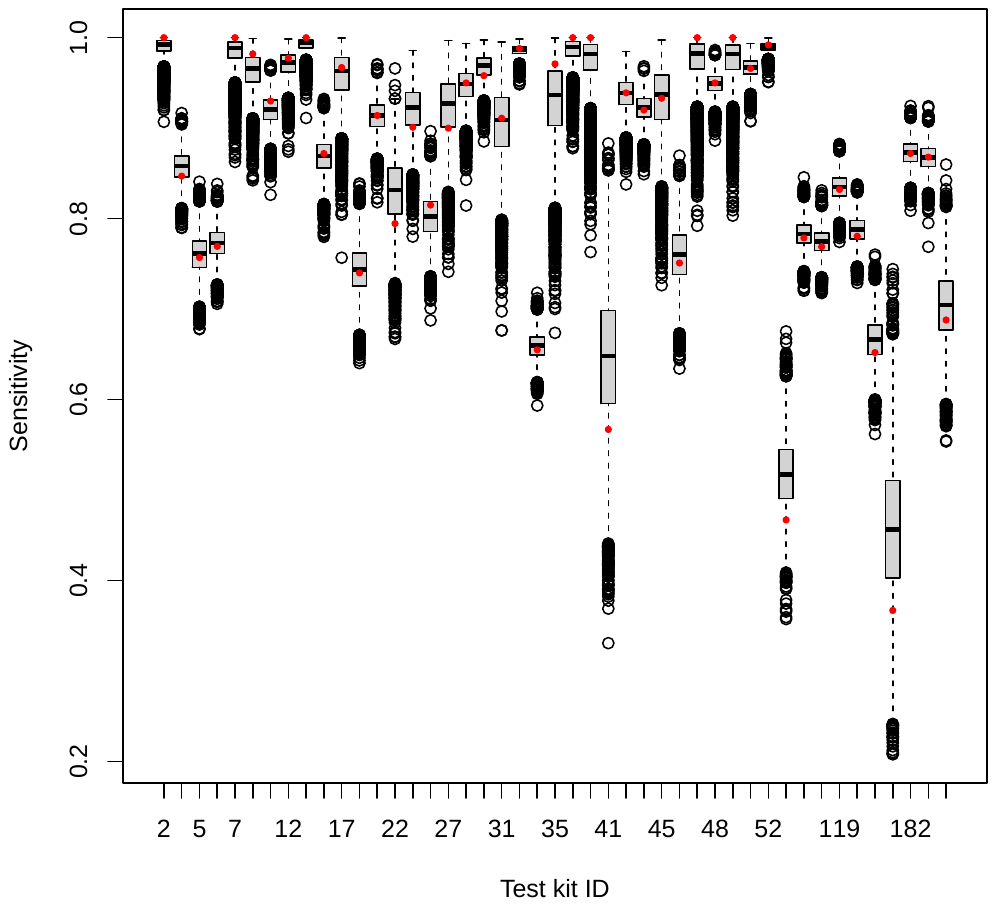

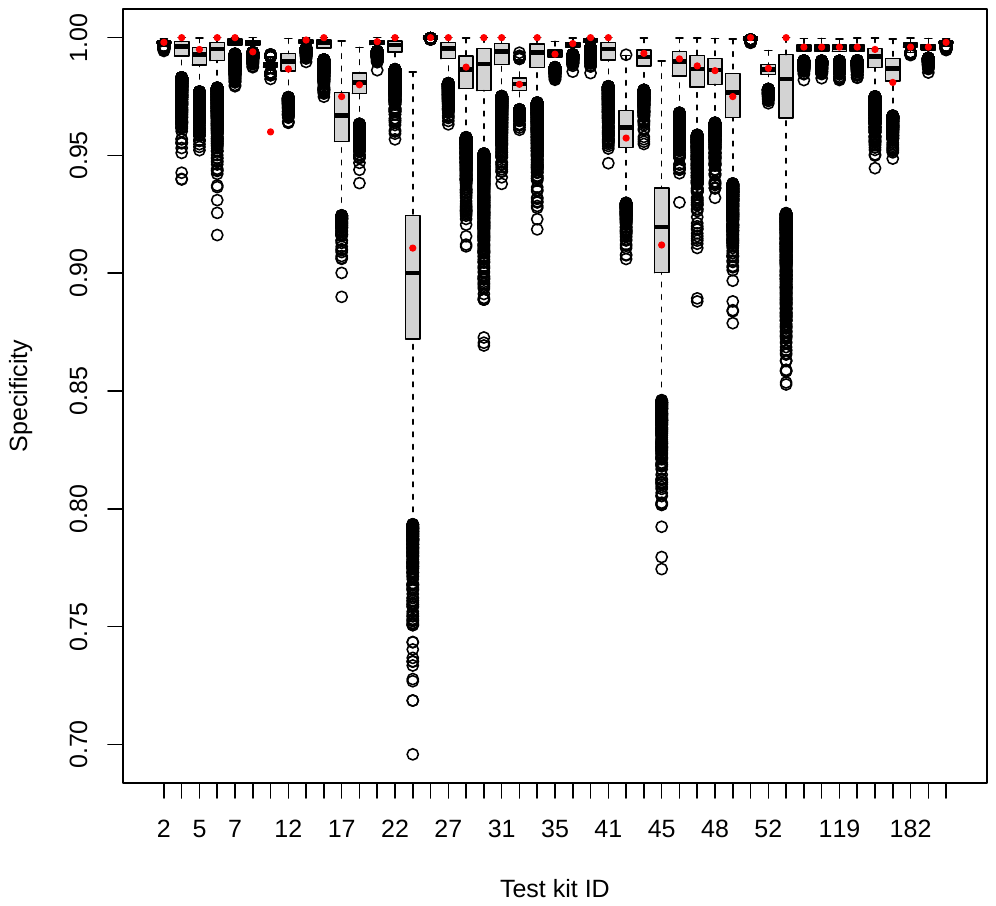

(a)	(b)

- for links to the corresponding assay names see our GitHub repository under appendix material

### **Model Outputs**

For each model parameter, we use the posterior mean as the point estimate and produce 95% equal-tail credible intervals to describe uncertainty.

#### **Total seroprevalence**

Similar to calculating the average seroprevalence for a death age bin, we estimate total seroprevalence for a location by taking an average of the age bin seroprevalences, weighting by the population distribution at that location:

$\pi_{l,\left[ 0,100+ \right)}=\sum_{A\in\mathcal{A}_{\mathcal{l}}} \left[ \pi_{l,A}\sum_{b\in A\cap N} f_{l,A}\left( b \right) \right]$	(14)

where $\mathcal{A}_{\mathcal{l}}$ are the serology age bins associated with location $l$.

#### **Assessing Uniformity of Seroprevalence Across Age**

We calculated total seroprevalence from younger adults and middle aged adults, compared to older adults. The age bins used for each location were selected as follows:

- **Younger adults (approximately 18 to 59):** Any age bins such that 15 ≤ lower age *<* 60 and 20 *<* upper age ≤ 65 were included
- **Middle aged adults (approximately 40 to 59):** Any age bins such that 40 *<* lower age *<* 60 and 40 *<* upper age ≤ 65 were included
- **Older adults (approximately 60 and older):** Any age bins such that 60 ≤ lower age were included.

This resulted in sets of bins where the 18-59 bins and the 40-59 bins did not overlap the 60+ bins.

We then calculated total seroprevalence from age *a* to *b* using appropriate age bins, $\mathcal{A}$, similar to equation (8)

$\pi_{l,a-b}=\sum_{A\in\mathcal{A}} \pi_{l,A}\frac{n_{l,A}}{\sum_{B\in\mathcal{A}} n_{l,B}}$	(15)

where $\frac{n_{l,A}}{\sum_{B\in\mathcal{A}} n_{l,B}}$ estimates the percent of the total population in that age bin, assuming representative age distributions in the serology studies.

For each draw from the posterior distribution, we calculated $\pi_{l,a-b}$ for each of our three age intervals of interest. We then calculated the ratios $\frac{\pi_{l,60+}}{\pi_{l,18-59}}$ and $\frac{\pi_{l,60+}}{\pi_{l,40-59}}$ for each draw.

#### **Total IFR and Comparison to High-income (EJE) Prediction**

##### *Total IFR*

Suppose location $l$ has death age bins $\mathcal{A}_{\mathcal{l}}$. Let $\sum_{b\in A\cap N} f_{l,A_{i}}\left( b \right)=\text{pop}_{l,A}$ for $A\in\mathcal{A}_{\mathcal{l}}$. Then

$$\text{IFR}_{\text{total}}=\frac{\text{number of deaths}}{\text{number of infections}}$$

$$=\frac{\sum_{A\in\mathcal{A}_{\mathcal{l}}} \text{IFR}_{l,A}\times\pi_{l,A}\times\text{pop}_{l,A}}{\sum_{B\in\mathcal{A}_{\mathcal{l}}} \pi_{l,B}\times\text{pop}_{l,B}}$$

$=\sum_{A\in\mathcal{A}_{\mathcal{l}}} \text{IFR}_{l,A}\times\left( \frac{\pi_{l,A}\times\text{pop}_{l,A}}{\sum_{B\in\mathcal{A}_{\mathcal{l}}} \pi_{l,B}\times\text{pop}_{l,B}} \right)$	(16)

estimates the total IFR for location $l$. By calculating IFR_total_ for each posterior sample, we can then obtain posterior mean and credible intervals for IFR_total_.

##### *High income country benchmark*

We compare the IFR estimate to a high income country benchmark based on results from Levin et. al. (4) which found a log-linear relationship between age and IFR. Define

$\text{HICB}_{a}=\left\{ \begin{aligned} \int_{a}^{a+1} \frac{1}{100}{10}^{-3.27+0.0524a}da \text{if }a<85 \\ \frac{1}{100}{10}^{-3.27+0.0524\left( 85 \right)}\text{ if }a\geq85 \end{aligned}. \right.$	(17)

Then HICB*_a_* represents the IFR predicted by the high income countries line averaged over the interval [*a,a*+ 1) for ages less than 85 and assumes the high income countries line flattens out and becomes uniform for ages 85 and older.

Then if we assume uniform prevalence for a location, the total IFR estimate over age bin *A* for high income countries is

$\text{HICB}_{A}=\frac{\sum_{a\in A} \text{EJE}_{a}\times f_{l}\left( a \right)}{\sum_{b\in A} f_{l}\left( b \right)}.$	(18)

##### *Subsetting to ages 18-65*

To estimate the IFR between ages 18 and 65, we used the same strategy in picking age bins as we did when testing uniform prevalence. That is, we selected $\mathcal{B}_{\mathcal{l}}$ to be the death age bins in $\mathcal{A}$ such that the lower age of the bin is greater than or equal to 18 and the upper age of the bin is less than 66. We then applied equation (16), replacing $\mathcal{A}_{\mathcal{l}}$ with $\mathcal{B}_{\mathcal{l}}$. We were not able to calculate the IFR between 18 and 65 for locations there were no age bins in $\mathcal{B}_{\mathcal{l}}$.

##### *Baseline population*

In order to compare the impact of the age specific IFR while controlling for population age distribution, we calculated the Total IFR substituting $f_{l}\left( a \right)$ for a baseline population age distribution, $\bar{f}\left( a \right)$ in (16). We calculated $\bar{f_{A}}\left( a \right)$ following (7). The baseline population was calculated as a median across locations for each age, then rescaled to sum to one:

$\bar{f_{\star}}\left( a \right)=\text{median}\left\{ f_{l}\left( a \right) \mid l\text{ is one of the observed locations with fatality data} \right\}$	(19)

$\bar{f}\left( a \right)=\frac{\bar{f_{\star}}\left( a \right)}{\sum_{a=0}^{85} \bar{f_{\star}}\left( a \right)}.$	(20)

We also considered taking the mean across locations for each age and taking the mean across locations for each age after removing the top five and bottom five locations for that age (a censored mean). All three approaches gave similar values as shown in Figure A3.

*Baseline age distribution*

Various estimates of baseline age distribution (mean, median, and censored mean) plotted on age distribution for observed locations in gray:

**Figure A3 – Estimates of Baseline Age Distribution**

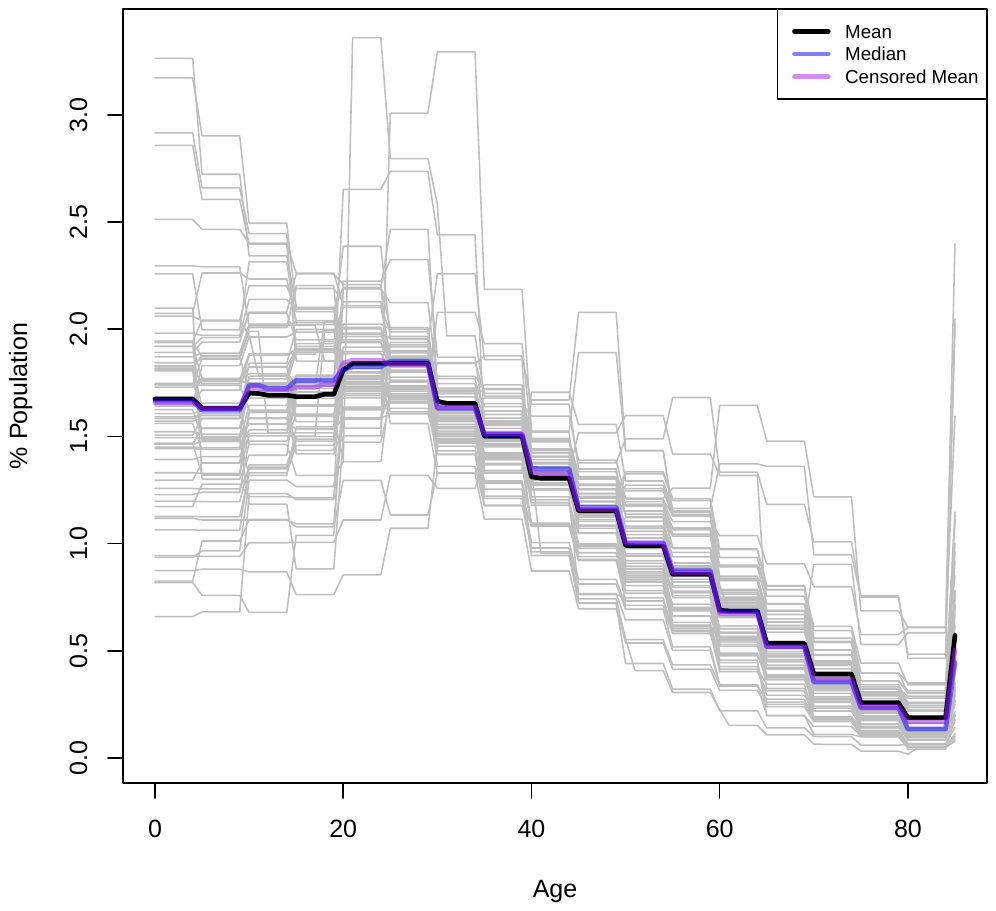

##### *Country average*

To get an average total IFR estimate for each country, we took a weighted average of the total IFR estimate of the locations within that country. We chose to weight by $1/\sqrt{n_{l}}$ in order to give more weight to locations with more certain seroprevalence estimates. In locations with multiple age bins, we took the average across $n_{l,A}$ as $n_{l}$. We weighted by certainty in the seroprevalence estimates rather than the IFR estimates because larger IFR estimates tend to be due to small seroprevalence estimates and consequently have more uncertainty (i.e., small differences in the denominator, seroprevalence, can result in large changes in the IFR estimate when seroprevalence is small). We did not want to bias the average by down weight all of the larger IFR estimates unless there was a smaller sample size in the seroprevalence.

We followed the same process to estimate the country average IFR between 18 and 65, with the added step of removing any locations in the country where $\mathcal{B}_{\mathcal{l}}$ was empty.

b. **Seroconversion and Seroreversion**

**Seroconversion**

For seroprevalence to approximate the number of people infected, almost all infected people need to seroconvert by increasing antibody levels after infection. Studies of large populations suggest that >85% or >90% of SARS-CoV-2-infected individuals seroconvert by approximately 2 weeks after infection (30-33). This increases confidence in the accuracy of seroprevalence-based infection estimates that use tests with sufficiently high sensitivity (30, 34). Moreover, >50% of the total population seroconverted in several locations, which would not occur if a substantial proportion of infected individuals failed to seroconvert. The table below illustrates this with studies reporting >50% seroprevalence before the onset of widespread SARS-CoV-2 vaccination:

**Table A2 - Locations with >50% Reported Seroprevalence**

| **Region** | **Location** | **Reported seroprevalence** | **Number tested for seroprevalence estimate** |
| --- | --- | --- | --- |
| Latin America | Argentina: Buenos Aires (Barrio Padre Mugica) | 53.4%  (CI: 52.8 - 54.1%) | 873 |
|  | Argentina: Metropolitan Area of Buenos Aires  (17 de Noviembre) | 56.7%  (CI: 55.8 - 57.6%) | 300 |
|  | Colombia, 10 cities: Barranquilla | 53%  (CI: 41 - 65%) | 1426 |
|  | Colombia, 10 cities: Guapi | 78%  (CI: 65 - 91%) | 721 |
|  | Colombia, 10 cities: Leticia | 62%  (CI: 51 - 73%) | 1417 |
|  | Colombia, Córdoba: Montería | 55.3%  (CI: 52.5 - 57.8%) | 1368 |
|  | Peru: Iquitos; July,  August | 70% (CI: 67 - 73%)  66% (CI: 62 - 70%) | 716  621 |
| Africa | Ethiopia:  Addis Ketema | 54.2%  (CI: 47.5 - 60.7%) | 218 |
| Middle East | Afghanistan: Kabul | 53% (CI: < +/-6%) | - |
|  | Iran, 18 cities: Qom,  Rasht | 58.5% (CI: 37.2 - 83.9%)  72.6% (CI: 53.9 - 92.8%) | 108  99 |
|  | Iraq: Duhok city | 62.6% | 743 |
| South Asia | Bangladesh: Dhaka (“slums”) | 74% | - |
|  | India: Delhi | 56.1%  (CI: 55.5 - 56.8%) | 28,169 |
|  | India: Hyderabad | 54.2%  (CI: 53.2 - 55.2%) | 9363 |
|  | India: Karnataka (urban areas) | 53.8%  (CI: 48.4 - 59.2%) | 453 |
|  | India: Mumbai, 3 “slums” (in: Chembur West,  Dahisar,  Matunga) | 56.4%  55.1% (CI: 52.4 - 57.8%)  51.1% (CI: 46.4 - 55.8%)  57.0% (CI: 54.7 - 59.2%) | 4202  1511  570  2121 |
|  | India: Pune, 5 subwards  (Lohiyanagar-Kasewadi,  Navi Peth-Parvati,  Yerwada) | 51.3% (CI: 39.9% - 62.4%)  66.4% (CI: 57.8% - 74.1%)  54.1% (CI: 48.3% - 61.7%)  55.5% (CI: 46.6% - 64.1%) | 1659  307  331  367 |

- CI: confidence interval

- this analysis is restricted to studies with at least 75 people tested

- “*Seroprevalence Comparison to High-income Countries*” shows our calculated seroprevalence estimates for some of these locations

- for links to the studies at each location see our GitHub repository under appendix material

These high seroprevalence estimates may shed light on high vs. low herd immunity thresholds (35-37). For example, Leticia suffered another wave of SARS-CoV-2 infections after reported seroprevalence of 62% (6), as did Delhi after reported seroprevalence of 56% (38), the state of Maranhão after reported seroprevalence of 40%, and Jordan after reported seroprevalence of 34% (39). Cross-reactivity also likely does not account for elevated seroprevalence in many of the regions listed in table A2, since cross-reactivity did not significantly reduce test specificity in locations such as Colombia, Ethiopia, and Iran (40-42). These high seroprevalence estimates instead imply that the vast majority of infected individuals seroconverted, increasing the reliability of seroprevalence-based infection estimates.

**Seroreversion Estimates and Correcting for Test Sensitivity + Specificity**

Another area of concern is seroreversion. Briefly, seroreversion occurs when the specific antibodies a serological assay tests decline to below the assay’s level of detection, preventing the assay from identifying infected individuals.

As many studies conducted in developing countries were performed long after initial COVID-19 waves passed, the risk of seroreversion could be high. This could lead to underestimation of the proportion of infected people and thus unreliable estimates in our computed IFRs. Moreover, any modelling using assumptions about seroreversion for one serological test would almost certainly lead to errors in other places as different tests can substantially differ in characteristics (43).

It is not only important to account for decreased test sensitivity that results from seroreversion, but also to correct for test sensitivity and specificity even in the absence of seroreversion. For example, some serological assays display lower specificity in African populations, possibly due to cross-reactivity with other pathogens (41, 44). This may contribute to divergent seroprevalence estimates between two studies performed in Addis Ababa, Ethiopia (42, 45) (see “*Out-of-Sample Analysis*” for further discussion). However, specificity likely remains high in African populations for many of the assays used in our included studies (42, 46, 47).

The traditional method (Gladen-Rogan correction (29)) of adjusting for specificity and sensitivity assumes test characteristics are precisely known, without accounting for the uncertainty that comes with inferring characteristics from a limited number of tested samples (48). To account for this uncertainty, we incorporated a Bayesian test correction.

We also conducted a review to assess serological tests for risk of seroreversion and catalogued both the test manufacturer’s estimates of sensitivity as well as third-party assessments of the test. The review was restricted to studies that tested the same individuals at two different time-points separated by at least two months, or tested individuals at least two months after their first known positive test for SARS-CoV-2. We placed emphasis on commercial assays or assays used in seroprevalence studies. The search used the terms “COVID-19 seroreversion”, “COVID-19 longitudinal, antibody waning” in Medrxiv, Biorxiv, Google Scholar, and SSRN. Searches were completed at least monthly from March 2021 to June 2021, with the final search performed on June 30, 2021. We supplemented this with seroreversion studies found during searches up to July 14 for seroprevalence studies with representative sampling, and for which further information was released after July 14 (see “*Systematic Review Methodology*”).

We then classified serological assays as having low, medium, or high risk of seroreversion, based on the extent to which assay sensitivity decreased overtime. In classifying assays, priority was given to testing of those from a representative sample of the general population. Our review revealed that the Wondfo assay suffered from high risk of seroreversion and variation in test sensitivity between assay batches (49-51). So seroprevalence studies using this assay were excluded from our IFR and seroprevalence analyses.

**Table A3 – Assay Seroreversion Risk Based on Serial Testing**

| **Risk of seroreversion** | **Assay** | **Studies** |
| --- | --- | --- |
| high | Abbott Architect SARS-CoV-2 IgG  (anti-nucleocapsid) | representative sample (31, 52-56)  non-representative sample (57-61) |
|  | Beckman ACCESS SARS-CoV-2 IgG  (anti-RBD) | representative sample (53) |
|  | EDI Novel coronavirus COVID19 IgG  (anti-nucleocapsid) | non-representative sample (62) |
|  | Wondfo SARS-CoV-2 IgG IgM  (anti-RBD) | representative sample  non-representative sample (49) |
| moderate | Biosynex COVID-19 BSS IgG/IgM  (anti-RBD) | non-representative sample (62) |
|  | Euroimmun anti-SARS-CoV-2 IgG  (anti-spike) | representative sample (63, 64)  non-representative sample (58, 63, 65) |
|  | Ortho VITROS IgG  (anti-spike) | non-representative sample (43) |
| low | Abbott SARS-CoV-2 Quant II IgG  (anti-RBD) | non-representative sample (62) |
|  | COVIDAR IgG  (anti-spike) | representative sample (66)  non-representative sample (67) |
|  | DiaSorin Liaison SARS-CoV-2 IgG  (anti-spike) | representative sample (31, 52)  non-representative sample (43, 60, 61) |
|  | Genetico CoronaPass total antibody  (anti-RBD) | representative sample (55) |
|  | InBios SCoV-2 Detect IgG  (anti-spike) | non-representative sample (65) |
|  | Netherlands in-house assay IgG  (anti-spike) | representative sample (68) |
|  | Ortho VITROS total Ig  (anti-spike) | non-representative sample (43) |
|  | Roche Elecsys anti-SARS-CoV-2 IgG IgM IgA  (anti-spike) | representative sample (56, 63) |
|  | Roche Elecsys anti-SARS-CoV-2 IgG IgM IgA  (anti-nucleocapsid) | representative sample (31, 56, 63, 69, 70)  non-representative sample (60, 61, 63, 71, 72) |
|  | Siemens SARS-CoV-2 Total, IgG IM  (anti-RBD) | non-representative sample (60, 61) |
|  | Sinai Health, Toronto; ELISA  (anti-nucleocapsid, anti-spike) | non-representative sample (73) |
|  | University of Rio de Janeiro (UFRJ); ELISA  (anti-spike) | non-representative sample (49, 74) |
|  | University of Oxford  (anti-RBD) (75, 76) | representative sample (77) |
|  | VectorBest ELISA IgG  (anti-spike) | representative sample (55, 78) |
|  | Wantai SARS-CoV-2 Total, pan-Ig  (anti-RBD) | representative sample (56, 79)  non-representative sample (80) |
|  | Combination: Abbott SARS-CoV-2 IgG + Euroimmun anti-SARS-CoV-2 IgG  (anti-nucleocapsid +  anti-spike, respectively) | representative sample (22) |
|  | Combination: Roche Elecsys anti-SARS-CoV-2 IgG IgM IgA + Forsa neutralization assay  (anti-nucleocapsid + neutralization assay) | representative sample (81) |

- RBD: receptor-binding domain, a component of the spike protein

Some seroprevalence studies tested a representative sample of the general population, including those with a prior positive SARS-CoV-2 PCR test weeks or months before serological testing, a previous COVID-19 diagnosis weeks or months before serology, etc. If many of these prior-positive individuals later tested seronegative, then that is unlikely to represent failed seroconversion, as previously discussed. It instead likely indicates a high risk of seroreversion during the time following their initial positive test (82). A threshold of <75% sensitivity was selected for this risk of seroreversion because at least 75% of prior-positives tested seropositive using the Roche assay that is at low risk of seroreversion (see table A3), and the vast majority of sources reported sensitivity of at least 75% before seroreversion, as shown in the input data of our GitHub repository.

The following tests are at medium or high risk of seroreversion based on less than 75% of prior-positives testing seropositive at the locations indicated:

**Table A4 – Medium or High Seroreversion Risk Based on Delayed Testing of SARS-CoV-2-infected Individuals**

| **Location** | **Assay** |
| --- | --- |
| Spain | Abbott Architect SARS-CoV-2 IgG (anti-nucleocapsid) |
| India: Delhi | ErbaLisa COVID-19 IgG  (anti-spike) |
| India: Paschim Medinipur |  |
| Germany: Mitte | Euroimmun anti-SARS-CoV-2 IgG  (anti-spike) |
| Germany: Straubing |  |
| England (REACT-2) | Fortress Diagnostics COVID-19 Total, IgG IgM (anti-spike) |
| Denmark (We Test) | Livzon COVID-19 IgG IgM (anti-nucleocapsid, anti-spike) |
| South Africa: Gauteng | Luminex in-house assay IgG (anti-RBD) |
| India: Delhi | Zydus COVID-19 Kavach IgG (whole virus antigen) |

- for links to the studies at each location see our GitHub repository under appendix material

Based on this analysis, we accounted for seroreversion in four ways detailed below. We could not, however, apply these correction methods to all the seroprevalence studies included in our analysis due to insufficient information for these assays. So in our GitHub repository under appendix material, we categorized a study as low risk of bias for seroreversion if we could account for seroreversion using one of these methods, and high risk of bias if we could not.

We accounted for seroreversion as follows:

1. *For assays at low risk of seroreversion based on table A3*: we applied no adjustment to reported test sensitivity and assumed a low risk of seroreversion.
2. *For studies in which less than 75% of prior-positives tested seropositive*: we treated sensitivity as equivalent to the proportion of prior-positives that tested seropositive. This approach is consistent with prior work on how waning of antibodies reduces the proportion of prior-positives who test seropositive (83{Mutevedzi, 2021 #848)}.
3. *For studies that used the Abbott Architect assay and did not report that less than 75% of prior-positives tested seropositive*: we used prior publications to detail how sensitivity decreased overtime for the Abbott assay (see table A5). We then compared this sensitivity decrease to the accumulation of reported SARS-CoV-2 cases for study locations that used this assay. From this we derived a weighted sensitivity specific to each location and which accounted for seroreversion decreasing sensitivity with time. This approach was not extended to other assays at high risk of seroreversion in table A3 because our included seroprevalence studies did not use these other assays and we lacked sufficient data to precisely detail how sensitivity decreased with time for these tests.
4. *For studies that did not use the Abbott Architect assay and in which at least 75% of prior-positives tested seropositive*: we applied no adjustment to reported test sensitivity and assumed a low risk of seroreversion.

**Table A5 – Studies Used for Abbott Architect Sensitivity Overtime**

| **Type of Sampling Drawn from** | **Location of Information in Source** |
| --- | --- |
| representative(52) | main text |
| representative (53) | table S3 |
| representative (31) | figure 3 |
| non-representative(60) | figure 1 |
| non-representative(57) | figure 2, supplementary figure 2 |
| non-representative(58) | tables 1 and 2 |
| non-representative(59) | table 2 |
| non-representative(84) | table 1 |

3. **Additional Results and Figures**

**a. Age-specific IFRs**

**Figure A4 – Children IFR**

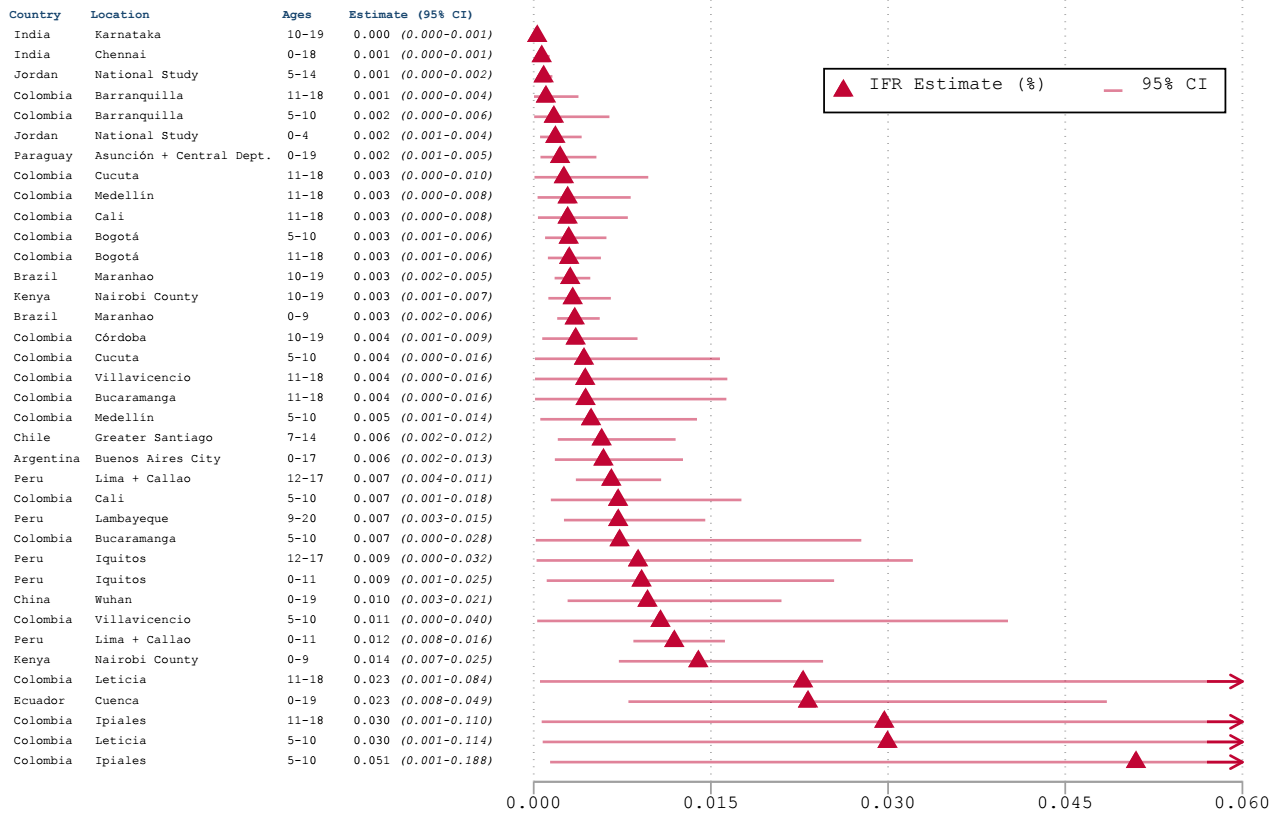

- for links to the studies at each location see our GitHub repository under appendix material

**Figure A5 – Young Adults IFR**

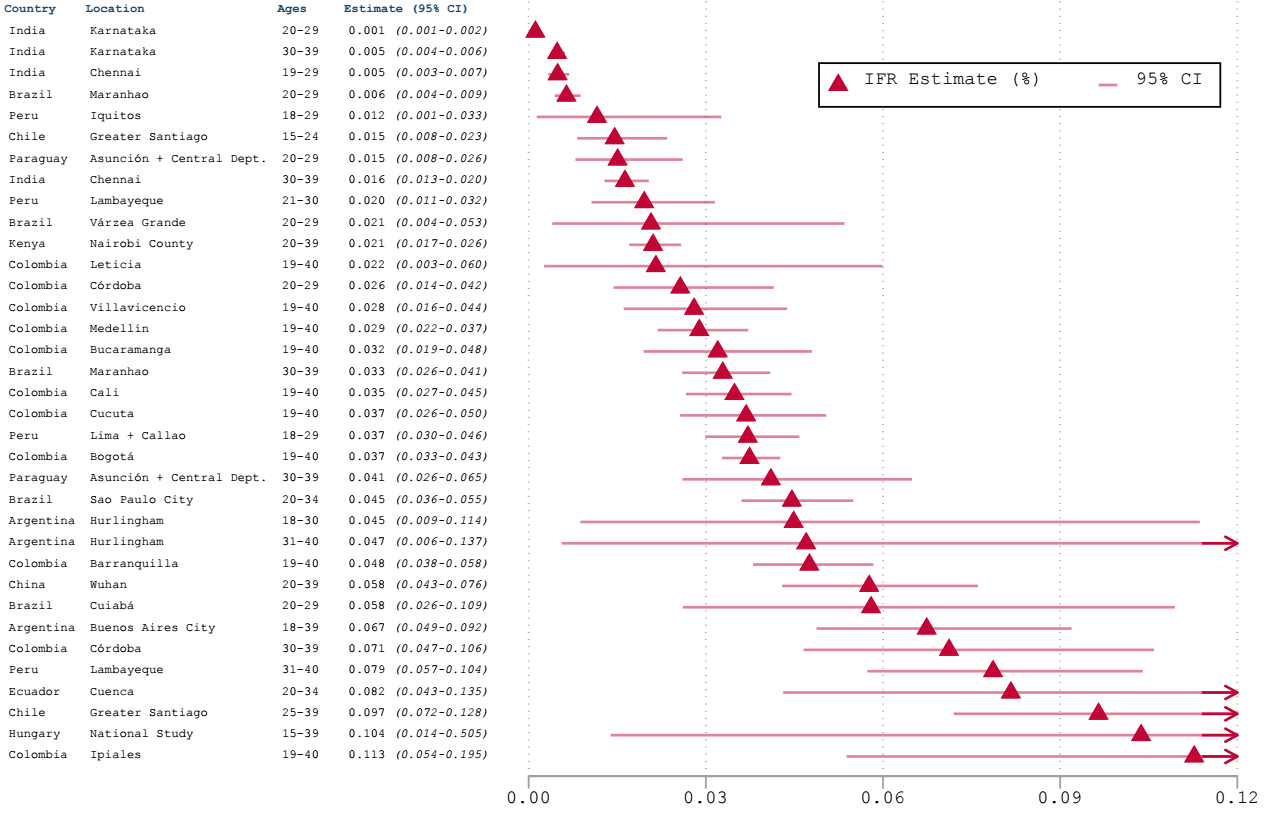

- for links to the studies at each location see our GitHub repository under appendix material

**Figure A6 – Middle-aged Adults IFR**

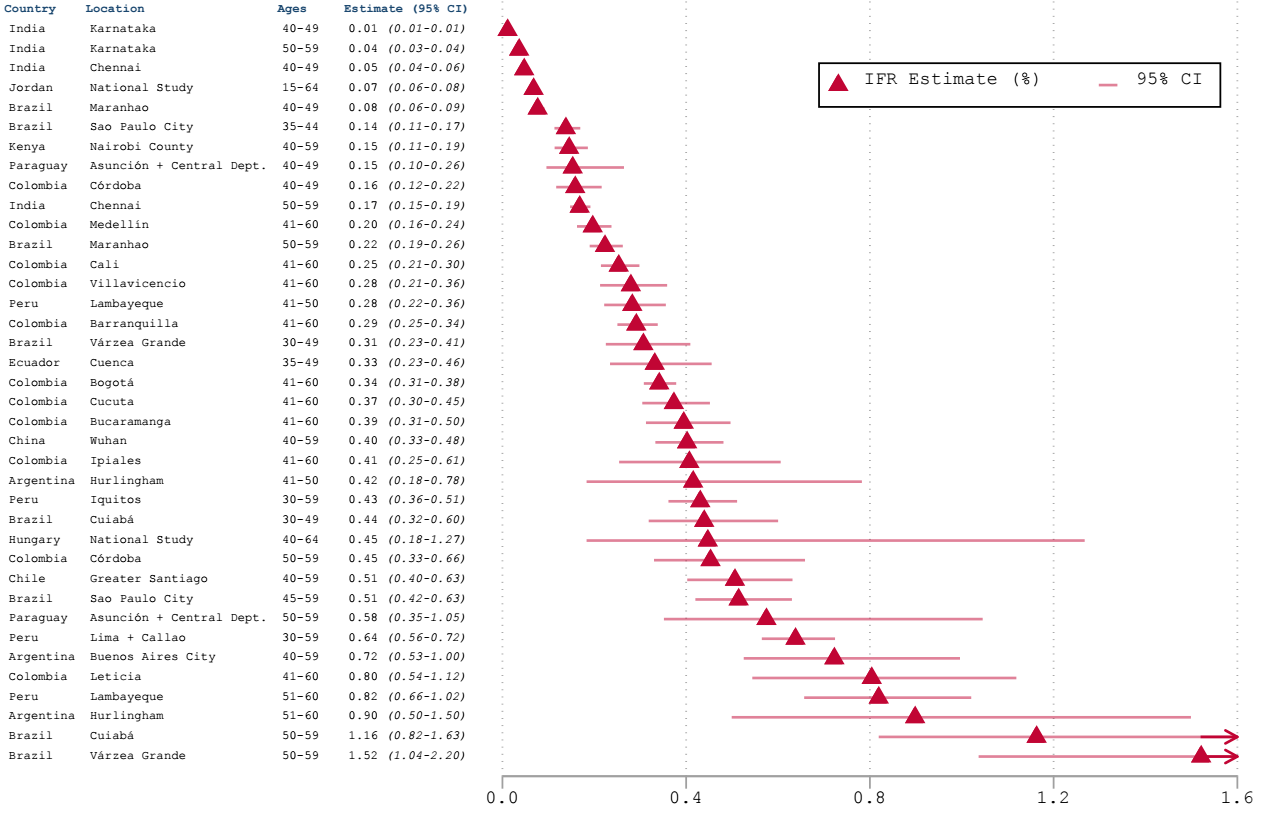

- for links to the studies at each location see our GitHub repository under appendix material

**Figure A7 – Older Adults IFR**

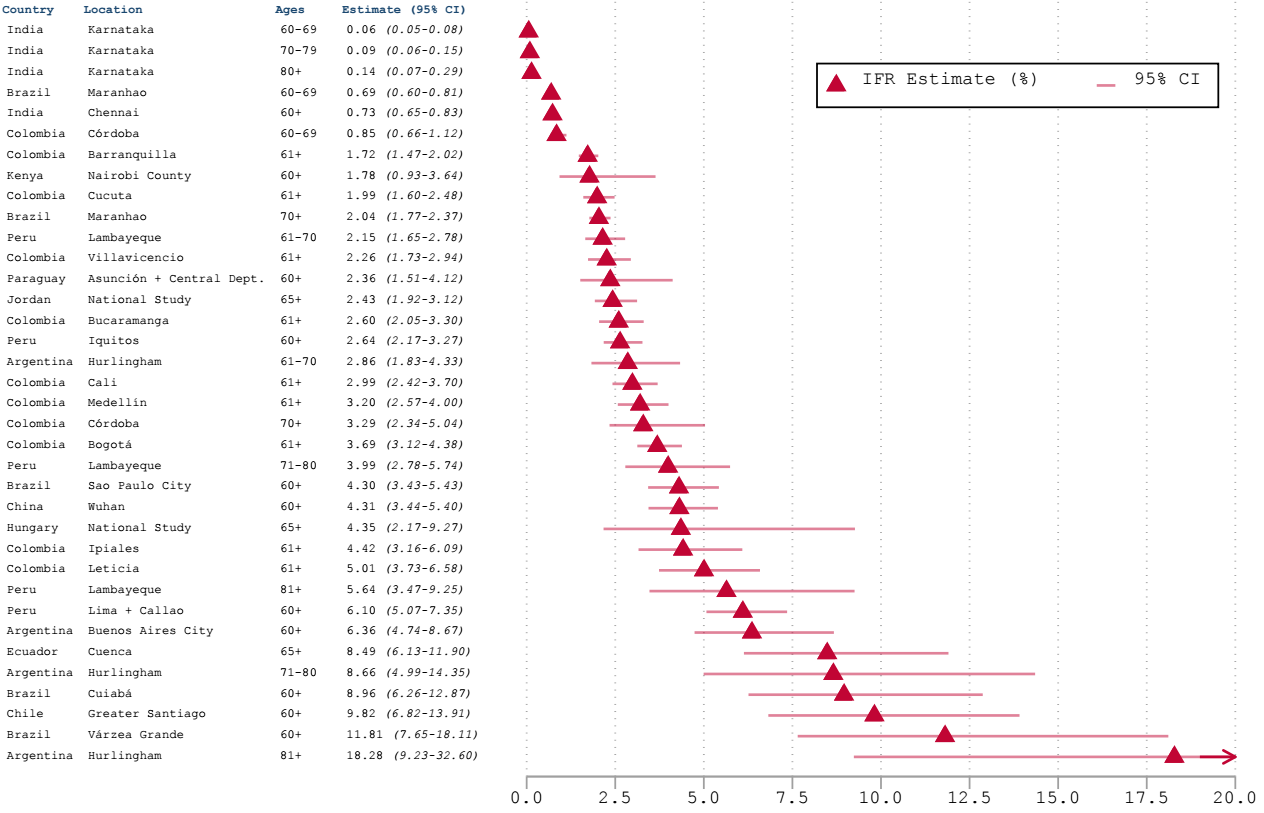
- for links to the studies at each location see our GitHub repository under appendix material

**b. Metaregression Results (Additional)**

For our metaregression, we took crude IFRs estimated for each age group, categorized them by median age, and used the meta regress command in Stata with random-effects and the Knapp-Hartung method for standard error adjustment. This used only studies from countries in which >50% of deaths were well-certified in the past decade (18), as discussed in the body of the paper:

**Figure A8 – Age-specific IFR Metaregression in Levels**

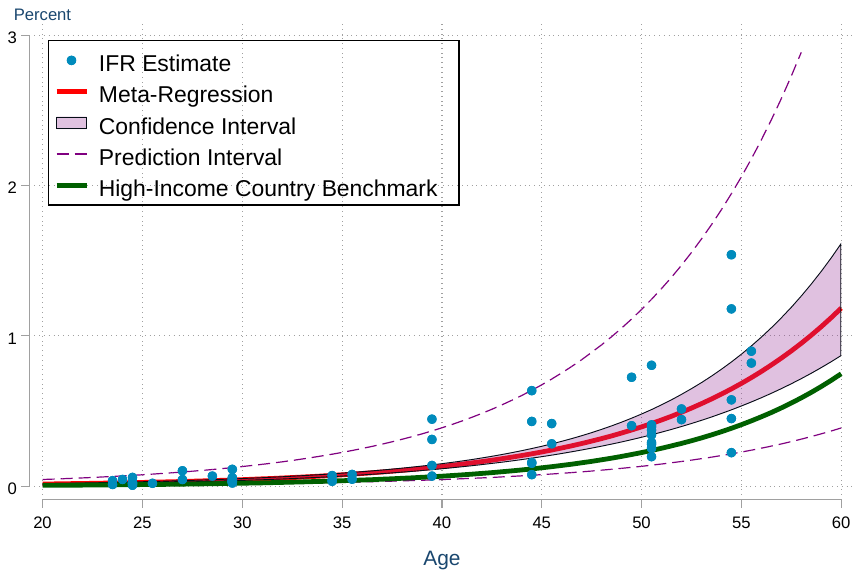

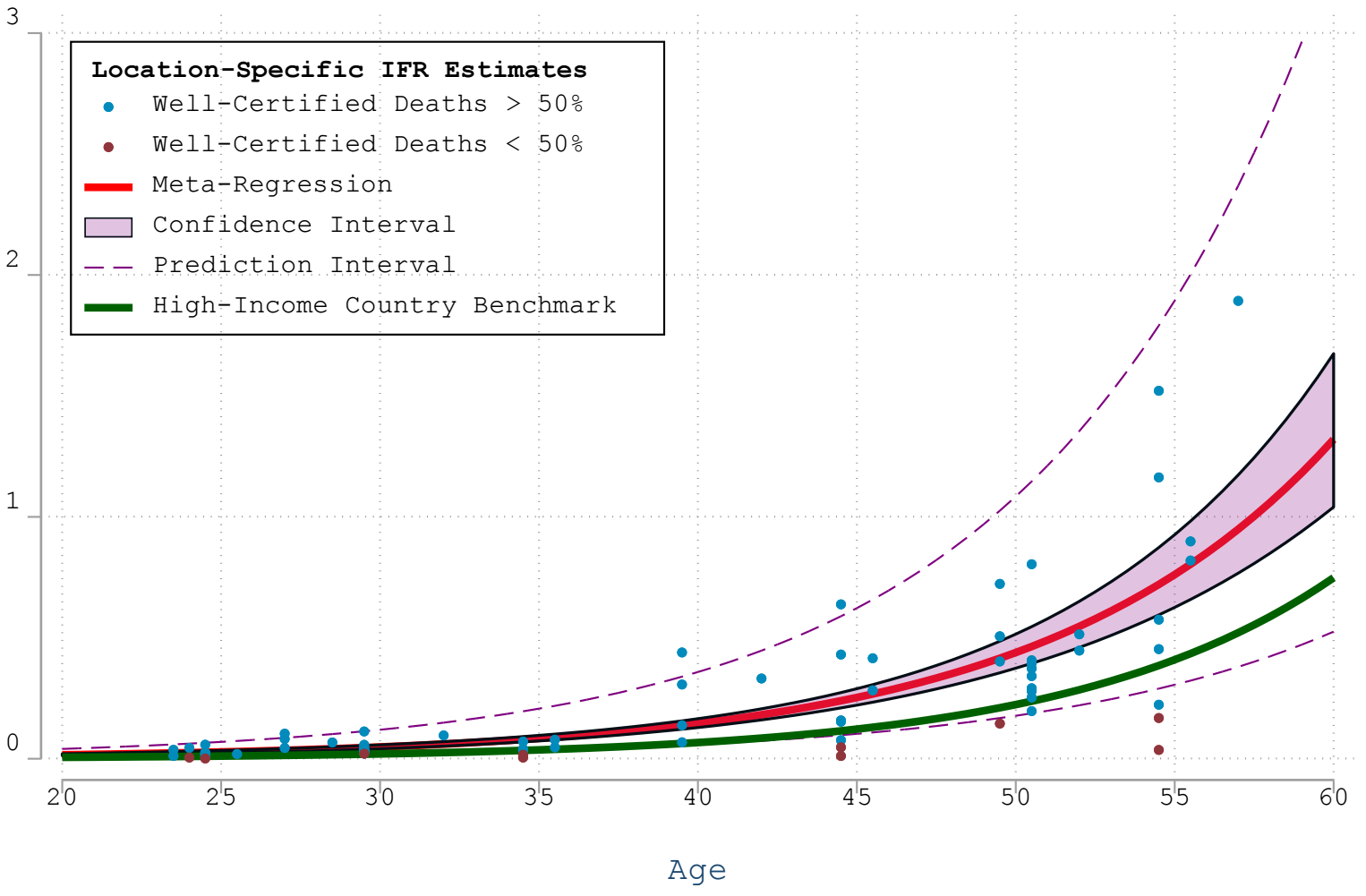

- for links to the studies at each location and categorization by percentage of deaths well-certified (18), see our GitHub repository under appendix material

**c. Population IFR**

**Figure A9 – Population IFR**

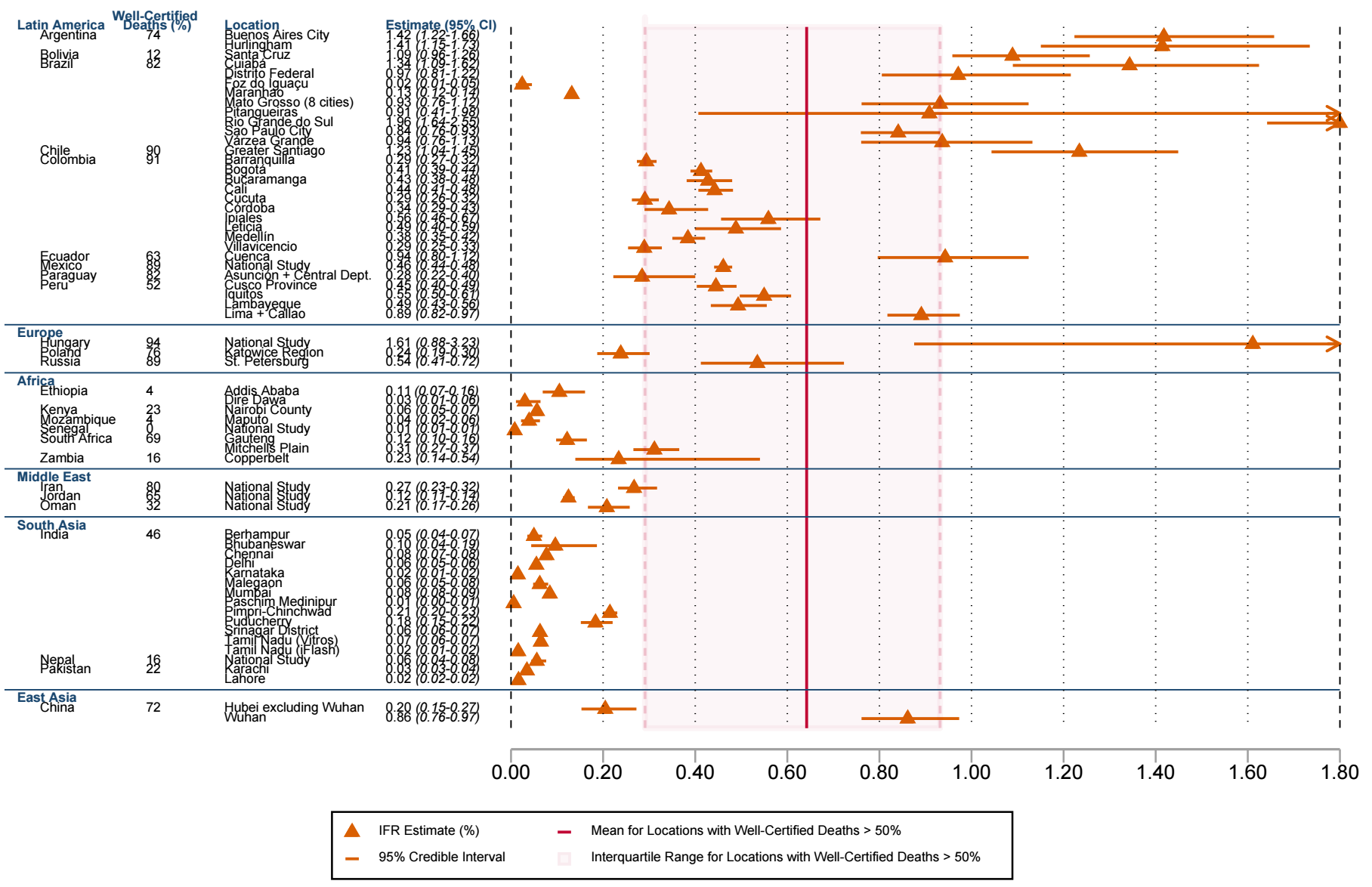

- for links to the studies at each location and categorization by percentage of deaths well-certified (18), see our GitHub repository under appendix material

d. **Risk of Bias Ratings**

Prior studies assessed serology studies for risk of bias with respect to factors such as representative or random sampling, adjustment for test characteristics, and non-response bias (1-3). Risk of bias assessments can differ for reasons including incomplete information on study design due to reliance on unofficial sources and assigning high risk of bias due to absence of test adjustments.

We mitigate these sources of bias by performing test adjustments and including only studies with official sources. Our review also includes only seroprevalence studies with representative sampling of the general population. In addition to non-representative sampling, other factors can bias seroprevalence estimates, such as non-response bias that may artificially increase or decrease estimates (55, 85, 86). Therefore, in our GitHub repository under appendix material we assess studies for risk of bias with respect to non-response bias, risk of seroreversion, and risk of COVID-19 death under-estimation based on percent well-certified deaths in the past decade (18) or in comparison to excess deaths.

e. **Covariate Examination**

**Table A6 – Correlations Between Covariates, IFR, and Well-Certified Deaths**

| **Covariate** | **Population IFR** | **Well-Certified Deaths** |
| --- | --- | --- |
| Human Development Index | 0.63 (0.27-0.83) | 0.90 (0.78-0.96) |
| Log of GDP per capita | 0.60 (0.23-0.82) | 0.88 (0.73-0.95) |
| Log(Healthcare Spending) | 0.60 (0.22-0.82) | 0.93 (0.82-0.97) |
| Log of GNI per capita | 0.60 (0.22-0.82) | 0.88 (0.72-0.95) |
| Hospital Beds Per Capita | 0.57 (0.18-0.80) | 0.48 (0.06-0.76) |
| Universal Health Coverage Index | 0.55 (0.16-0.79) | 0.95 (0.88-0.98) |
| Skilled Healthcare Workers Per Capita | 0.49 (0.08-0.76) | 0.69 (0.36-0.86) |
| Global Health Security Index | 0.47 (0.05-0.75) | 0.59 (0.21-0.81) |
| Life Expectancy at Birth | 0.43 (0.0-0.73) | 0.71 (0.40-0.88) |
| Healthy Life Expectancy at Age 60 | 0.40 (-0.04-0.71) | 0.83 (0.61-0.93) |

This table demonstrates the relationship between various covariates (18), IFR, and the measure of well-certified deaths. This shows that well-certified death is likely to be a primary explanatory variable, which is confounded by relationships with GDP and other national measures when these are used instead. An example of this is shown in the Directed Acyclic Graph below, made using the Daggity online tool: [http://www.dagitty.net/dags.html#](http://www.dagitty.net/dags.html)

**Figure A10 – Directed Acyclic Graph of Covariate Relationships**

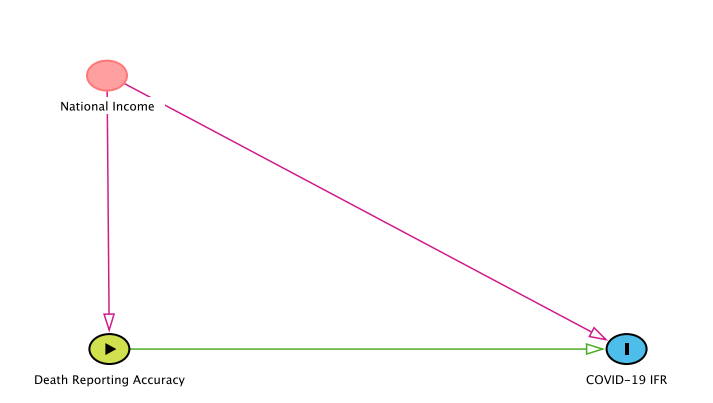

f. **Seroprevalence Comparison to High-income Countries**

Many high-income countries may have limited SARS-CoV-2 transmission more than developing countries (87-89). To illustrate this, the table and figure below compare seroprevalence estimates for national studies of high-income countries and developing countries with representative sampling before March 2021:

**Table A7 – Reported Seroprevalence for National Studies**

| **Country classification** | **Location** | **Reported seroprevalence** | **Median date of sampling** | **Adjustment for sensitivity / specificity** |
| --- | --- | --- | --- | --- |
| high-income country | Andorra | 11.0% | May 15 | unadjusted |
|  | France | 4.5% (CI: 3.9 - 5.0%) | May 17 | unadjusted |
|  | Italy | 2.5% (CI: 2.3 - 2.6%) | June 19 | unknown |
|  | Ireland | 1.7% (CI: 1.1 - 2.4%) | July 4 | unknown |
|  | Canada (Ab-C) | high specificity: 1.7%  (CI: 1.4 - 2.0%)  high sensitivity: 3.5%  (CI: 3.1 - 4.0%) | July (June – August) | unknown |
|  | Denmark  (We Test) | 0.8% | October 6 | unadjusted |
|  | USA | 11.9% (CI: 10.5 - 13.5%) | October 30 | Bayesian |
|  | South Korea | 0.3% (CI: 0.06 - 0.6%) | October 31 | Bayesian |
|  | Germany  (BUND) | 1.1% (CI: 0.9 - 1.3%) | November 6 | unadjusted |
|  | Germany  (SOEP) | 1.7% (CI: 1.2 - 2.3%) | November 11 | Rogan-Gladen |
|  | Slovenia | 4.1% (CI: 3.0 - 5.2%) | November 11 | Bayesian |
|  | Austria | 4.7% (CI: 3.8 - 5.6%) | November 13 | unadjusted |
|  | Great Britain | 8.8% (CI: < +/-1.0%) | November 17 | unknown |
|  | Spain | 9.9% (CI: 9.4 - 10.4%) | November 22 | unknown |
|  | Denmark (SSI) | 4.1% (CI: 3.1 - 4.9%) | December 16 | Rogan-Gladen |
|  | England (ONS) | 15.3% (CI: 14.7 - 15.9%) | January 4, 2021 | unknown |
|  | Norway | 0.9% (CI: 0.7 - 1.0%) | January 5 | unknown |
|  | England (REACT-2) | 13.9% (CI: 13.7 - 14.1%) | February 1 | Rogan-Gladen |
|  | Estonia | 11.5% (CI: 10.3 - 12.8%) | February 16 | unknown |
|  | Netherlands | ~14% | February | unknown |
| developing country | Hungary | 0.7% (CI: 0.5 - 0.9%) | May 8, 2020 | unadjusted |
|  | Afghanistan | 31.5% (CI: < +/-5%) | June | unknown |
|  | Iran | 14.2% (CI: 13.3 - 15.2%) | August 20 | Bayesian |
|  | Malaysia | 0.6% (CI: 0.4 – 0.9%) | September 6 | unadjusted |
|  | Mexico | 24.9% (CI: 22.2 - 26.7%) | September 30 | Rogan-Gladen |
|  | Nepal | 14.4% (CI: 11.8 - 17.0%) | October 15 | unknown |
|  | Mongolia | 1.5% (CI: 1.1 - 1.6%) | November 8 | Rogan-Gladen |
|  | Senegal | 28.4% (CI: 26.1 - 30.8%) | November 9 | adjusted (unknown method) |
|  | Oman | 22.0% (CI: 19.6 - 24.6%) | November 11 | unknown |
|  | Palestine | ~40% | December | unknown |
|  | Lebanon | 18.5% (CI: 16.8 - 20.2%) | December 26 | Rogan-Gladen |
|  | India | 24.1% (CI: 23.0 - 25.3%) | December 27 | Rogan-Gladen |
|  | Jordan | 34.2% (CI: 33.8 - 34.6%) | January 1, 2021 | unadjusted |

- CI: confidence interval

- for links to the studies at each location see our GitHub repository under appendix material

- for locations with multiple phases of sampling, only one late phase was included

- Brazil and Cape Verde were excluded due to use of the Wondfo assay

- Monaco, Tahiti/Moorea, and Pakistan were excluded due to insufficient information on study design and/or results

**Figure A11 – Population-wide Seroprevalence**

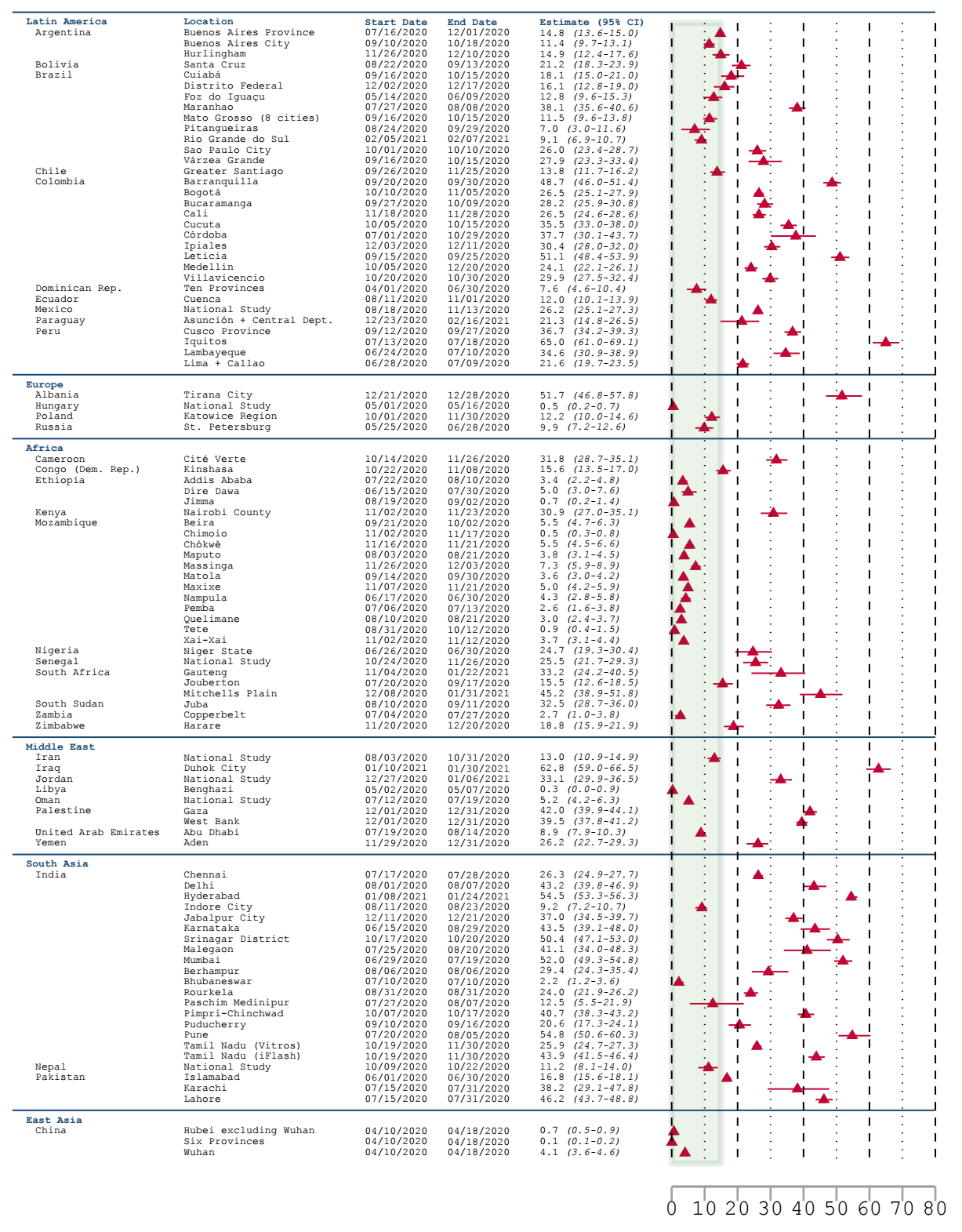

- green shading represents the range of national seroprevalence for high-income countries in table A7

- for links to the studies at each location see our GitHub repository under appendix material

East Asian countries such as Japan, South Korea, Mongolia, and Laos, were especially successful at limiting infection rates (90-93). Some subnational high-income country locations reported higher seroprevalence such as 42% in Ischgl, Austria (22), 20 - 25% in cantons of Switzerland (83), and 21% in New York City, USA (86). But to the best of our knowledge, no high-income countries reported non-vaccine-induced seroprevalence estimates of larger than 45% before March 2021, based on representative sampling of the general population. This contrasts with several subnational developing country locations reporting >50% seroprevalence, as shown in the figure above and in “*Seroconversion and Seroreversion*”, consistent with high-income countries’ greater success at limiting SARS-CoV-2 transmission. This success was also reflected in relatively lower infection rates in older populations that have the higher IFRs shown in “*IFRs by Age Group*” and “*Metaregression Results (Additional)*”:

**Figure A12 – Ratio of Seroprevalence for Older Adults (60+ years) Compared to Younger Adults (18-59 years)**

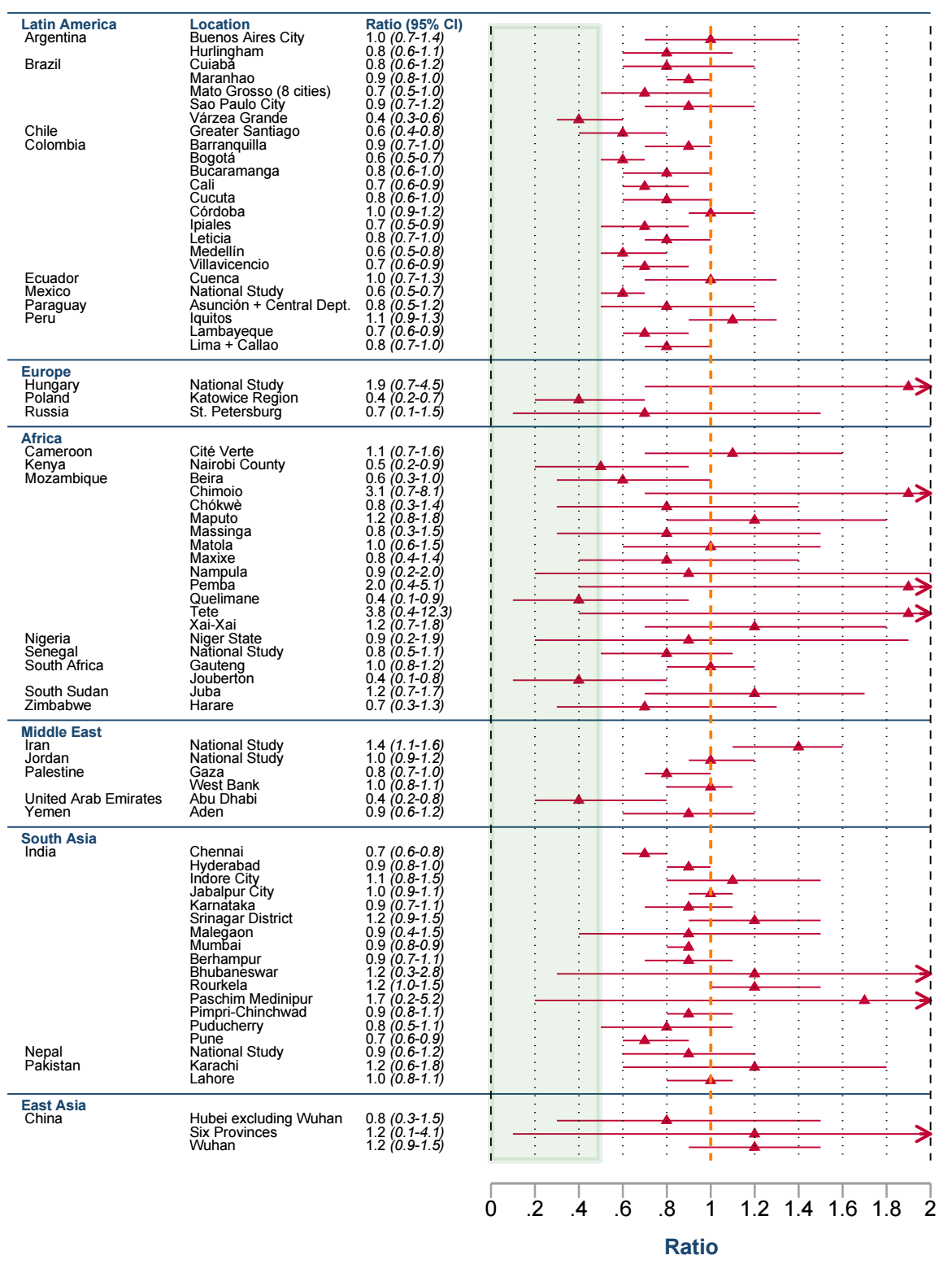

- green shading represents the range of national seroprevalence for high-income countries from our prior work (4)

- for links to the studies at each location see our GitHub repository under appendix material

g. **IFRs from Included Studies**

Early 2020 IFR estimates made mostly for China and high-income countries ranged from ~0.3% - ~1% (94, 95). For comparison, the table below lists reported IFRs from studies included in our literature search, and for which we did not generate an IFR estimate:

**Table A8 – Reported IFR in Studies for which a Meta-Analysis IFR was not Generated**

| **Location** | **IFR stated in the study** | **Type of death** | **Reason IFR estimate was not generated** |
| --- | --- | --- | --- |
| Brazil: Rio de Janeiro (multiple regions of the city) | Rio das Pedras: 0.2%  Maré: 0.3%  Rocinha: 0.3%  Cidade de Deus: 0.4%  Realengo: 1.2%  Campo Grande: 1.8% | reported | no death data 4 weeks post-midpoint,  insufficient information on assay |
| South Africa: Jouberton | wave 1: 0.12% (CI: 0.09 – 0.20%)  wave 1: 0.16% (CI: 0.13 – 0.23%)  wave 2: 0.50% (CI: 0.29 – 1.17%)  wave 2: 0.36% (CI: 0.24 – 0.72%) | excess  in-hospital  excess  in-hospital | no death data 4 weeks post-midpoint |
| South Africa:  Klerksdorp,  Pietermaritzburg | Klerksdorp:  0.3% (CI: 0.2 – 0.3%)  0.3% (CI: 0.3 – 0.3%)  Pietermaritzburg:  0.3% (CI: 0.3 – 0.3%)  0.6% (CI: 0.5 – 0.6%) | in-hospital  excess  in-hospital  excess | no death data 4 weeks post-midpoint |
| Sudan: Omdurman | 0.64% (CI: 0.62 – 0.75%) | excess | no death data 4 weeks post-midpoint,  sampling after February 2021,  study released after July 14 cutoff |
| Iran: Guilan province | 0.12% | reported | no death data 4 weeks post-midpoint |
| Iran: Mazandaran province | 0.33% | reported | no death data 4 weeks post-midpoint,  no stated start-week and end-week |
| Palestine | 0.11% | reported | no stated start-week and end-week |
| India: Indore (city, not district) | 0.17% | reported | no death data 4 weeks post-midpoint |
| India: Pune, 5 subwards* | 0.21% | reported | no death data 4 weeks post-midpoint |
| India: Tamil Nadu* | 0.05% | reported | Tamil Nadu split into 2 regions based on assay |

*age-specific IFR also reported

- CI: confidence interval

- for links to the studies at each location see our GitHub repository under appendix material

- another study reported age-specific, but not population-wide, IFR for Tamil Nadu (India) (96)

Results based on reported COVID-19 deaths in table A8 may under-estimate IFR due to poor death reporting, especially for developing countries that have a low percentage of deaths that are well-certified (18), as shown in the body of the paper. The table below compares reported IFRs from studies included in our literature search to our meta-analysis IFR estimates for the corresponding locations:

**Table A9 – Comparison of Meta-analysis IFRs to Reported IFRs**

| **Location** | **IFR stated in the study** | **Meta-analysis IFR** |
| --- | --- | --- |
| Brazil: Maranhão* | 0.14% (CI: 0.13 - 0.16%) [reported deaths]  0.28% (CI: 0.25 - 0.32%) [excess deaths] | 0.13% (CI: 0.12 - 0.14%) |
| Chile: Coquimbo-La Serena, Greater Santiago, and Talca | 1.67% (CI: 1.64 - 1.70%) | 1.23% (CI: 1.04 - 1.45%) |
| Colombia:  Córdoba (8 cities)* | 0.24% (CI: 0.23 - 0.25%) | 0.34% (CI: 0.29 - 0.43%) |
| Peru: Lambayeque | 0.5% | 0.49% (CI: 0.43 - 0.56%) |
| Poland: Katowice region | 0.62% (CI: 0.53 - 0.74%) | 0.62% (CI: 0.50 - 0.76%) |
| Russia: St. Petersburg (97)* | 0.83% (CI: 0.62 - 1.00%) [excess deaths] | 0.54% (CI: 0.41 - 0.73%) |
| Ethiopia:  Addis Ababa #2 | ≥0.09% | 0.20% (CI: 0.09 - 0.63%) |
| Kenya: Nairobi County* | 0.04% | 0.06% (CI: 0.05 - 0.07%) |
| South Africa:  Gauteng province | 0.28% (CI: 0.27 - 0.30%) [reported deaths]  0.67% (CI: 0.64 - 0.71%) [excess deaths] | 0.12% (CI: 0.10 - 0.17%) |
| South Africa:  Mitchells Plain | 0.3% (CI: 0.3 - 0.4%) [in-hospital deaths]  0.5% (CI: 0.4 - 0.6%) [excess deaths] | 0.31% (CI: 0.27 - 0.37%) |
| India: national | 0.08% (CI: 0.07 - 0.09%) to  0.11% (CI: 0.10 - 0.12%) | 0.06% (CI: 0.05 - 0.06%) |
| India: Chennai* | 0.17% (CI: 0.14 - 0.22%) | 0.08% (CI: 0.07 - 0.08%) |
| India: Delhi | 0.079% (CI: 0.076 - 0.081%) | 0.055% (CI: 0.05 - 0.06%) |
| India: Kashmir | 0.03% (CI: 0.03 - 0.04%) | 0.03% (CI: 0.026 - 0.030%) |
| India: Madurai district* | 0.04% (CI: 0.04 - 0.05%) | 0.03% (CI: 0.02 - 0.03%) |
| India: Mumbai (3 wards) | 0.12% | 0.08% (CI: 0.08 - 0.09%) |
| India: Pimpri-Chinchwad | 0.17% | 0.22% (CI: 0.20 - 0.23%) |
| India: Puducherry* | 0.08% | 0.18% (CI: 0.15 - 0.22%) |

*age-specific IFR also reported in the study

- CI: confidence interval

- for links to the studies at each location see our GitHub repository under appendix material

- IFRs are based on reported deaths, not excess deaths, unless otherwise noted

- another study reported age-specific, but not population-wide, IFRs for India from Mumbai (3 wards) and Karnataka (96)

Our meta-analysis IFRs in the table above may differ from reported IFRs for several reasons, including those discussed in “*Death Data*” and “*Seroconversion and Seroreversion*”, such as:

- corrections for test specificity, sensitivity, and seroreversion

- time lag from midpoint of serology sampling to date of death reporting

- deaths reported from case data vs. from “real time” official death totals vs. from excess deaths

h. **Out-of-Sample Analysis**

Our analysis excludes seroprevalence estimates that geographically overlap with an already included location, as discussed in “*Full Inclusion and Exclusion Criteria*”. The body of the paper also excludes IFR estimates that overlap with an included IFR, though we still calculated population-wide IFRs for these out-of-sample locations. The table below lists these IFR estimates:

**Table A10 – IFRs with Out-of-Sample Locations with Geographical Overlap**

| **Location** | **Excluded from body of paper** | **Meta-analysis IFR** |
| --- | --- | --- |
| Brazil: São Paulo* | No | 0.84% (CI: 0.76 - 0.93%) |
| Brazil: São Paulo #2* | Yes | 0.77% (CI: 0.66 - 0.88%) |
| Ethiopia: Addis Ababa* | No | 0.11% (CI: 0.07 - 0.16%) |
| Ethiopia: Addis Ababa #2 | Yes | 0.20% (CI: 0.09 - 0.63%) |
| Ethiopia: Addis Ababa #3* | Yes | 0.002% (CI: 0.001 - 0.005%) |
| India: national* | Yes | 0.06% (CI: 0.05 - 0.06%) |
| India: Kashmir (Srinagar district)* | No | 0.06% (CI: 0.06 - 0.07%) |
| India: Kashmir* | Yes | 0.03% (CI: 0.026 - 0.030%) |
| India: Tamil Nadu (Vitros districts) | No | 0.07% (CI: 0.06 - 0.07%) |
| India: Madurai district  (in Tamil Nadu)* | Yes | 0.03% (CI: 0.02 - 0.03%) |
| China: Wuhan* | No | 0.86% (CI: 0.76 - 0.97%) |
| China: Wuhan #2* | Yes | 0.71% (CI: 0.59 - 1.06%) |

*age-specific seroprevalence also reported in the paper

IFRs from studies of the same location may differ by sampling time due to factors such as improved treatment or new SARS-CoV-2 variants. Despite this, population-wide IFRs were relatively similar for studies that sampled the same location, as illustrated in the table above. These consilient results increase confidence that methodological differences between studies likely do not strongly bias our IFR estimates, in contrast to the order of magnitude difference in IFR between locations stratified by percentage of well-certified deaths, as shown in the body of the paper. Addis Ababa #3 remains the only outlier, possibly due to lower test specificity resulting from cross-reactivity (see “*Seroconversion and Seroreversion*”), low sample size in comparison to the other two Addis Ababa studies, or sampling in late April 2020 when under-estimation of COVID-19 deaths may have been greater than the July/August 2020 time period during which the other two studies sampled.

i. **Excess Mortality for Age-specific IFR**

**Table A11 – Ratio of Excess Mortality to Reported COVID-19 Deaths for Age-specific IFR Locations**

| **Argentina** | ***Buenos Aires City*** | 1.07 (CI: 1.0, 1.5) |
| --- | --- | --- |
| **Argentina** | ***Municipality of Hurlingham*** | 1.07 (CI: 1.0, 1.5) |
| **Brazil** | ***Maranhao*** | 1.41 (CI: 1.0, 2.4) |
| **Brazil** | ***Sao Paulo City*** | 1.02 (CI: 1.0, 1.3) |
| **Brazil** | ***Cuiaba, Mato Grosso*** | 1.00 (CI: 1.0, 1.0) |
| **Brazil** | ***Varzea Grande, Mato Grosso*** | 1.00 (CI: 1.0, 1.0) |
| **Chile** | ***Coquimbo-La Serena, Greater Santiago, Talca*** | 1.00 (CI: 1.0, 1.0) |
| **China** | ***Wuhan*** | 1.00 (CI: 1.0, 1.0) |
| **Colombia** | ***Leticia (Amazonas)*** | 1.09 (CI: 1.0, 1.6) |
| **Colombia** | ***Barranquilla (Atlantico)*** | 1.09 (CI: 1.0, 1.6) |
| **Colombia** | ***Medellin (Antioquia)*** | 1.09 (CI: 1.0, 1.6) |
| **Colombia** | ***Bucaramanga (Santander)*** | 1.09 (CI: 1.0, 1.6) |
| **Colombia** | ***Cucuta (Norte Santander)*** | 1.09 (CI: 1.0, 1.6) |
| **Colombia** | ***Villavicencio (Meta)*** | 1.09 (CI: 1.0, 1.6) |
| **Colombia** | ***Bogota*** | 1.09 (CI: 1.0, 1.6) |
| **Colombia** | ***Cali (Valle del Cauca)*** | 1.09 (CI: 1.0, 1.6) |
| **Colombia** | ***Ipiales (Narino)*** | 1.09 (CI: 1.0, 1.6) |
| **Colombia** | ***Cordoba: 8 cities*** | 1.09 (CI: 1.0, 1.6) |
| **Ecuador** | ***Cuenca (Azuay)*** | 1.01 (CI: 1.0, 1.1) |
| **Hungary** | ***National Study*** | 1.04 (CI: 1.0, 1.4) |
| **India** | ***Karnataka*** | 4.89 (CI: 2.6, 8.2) |
| **India** | ***Chennai*** | 4.80 (CI: 2.7, 7.9) |
| **Jordan** | ***National Study*** | 1.57 (CI: 1.0, 3.0) |
| **Kenya** | ***Nairobi County*** | 13.29 (CI: 7.1, 23.1) |
| **Paraguay** | ***Asuncion + Central Department*** | 1.10 (CI: 1.0, 1.6) |
| **Peru** | ***Lambayeque*** | 1.09 (CI: 1.0, 1.6) |
| **Peru** | ***Lima (Metropolitana) + Callao*** | 1.09 (CI: 1.0, 1.6) |
| **Peru** | ***Iquitos, Loreto*** | 1.09 (CI: 1.0, 1.6) |

*For Mexico, the WMD estimate of excess mortality ratio was based on only confirmed COVID-19 deaths. For comparability with other estimates, we adjusted this to the ratio of confirmed+suspected COVID-19 deaths.*

j. **PRISMA Flow Diagram**

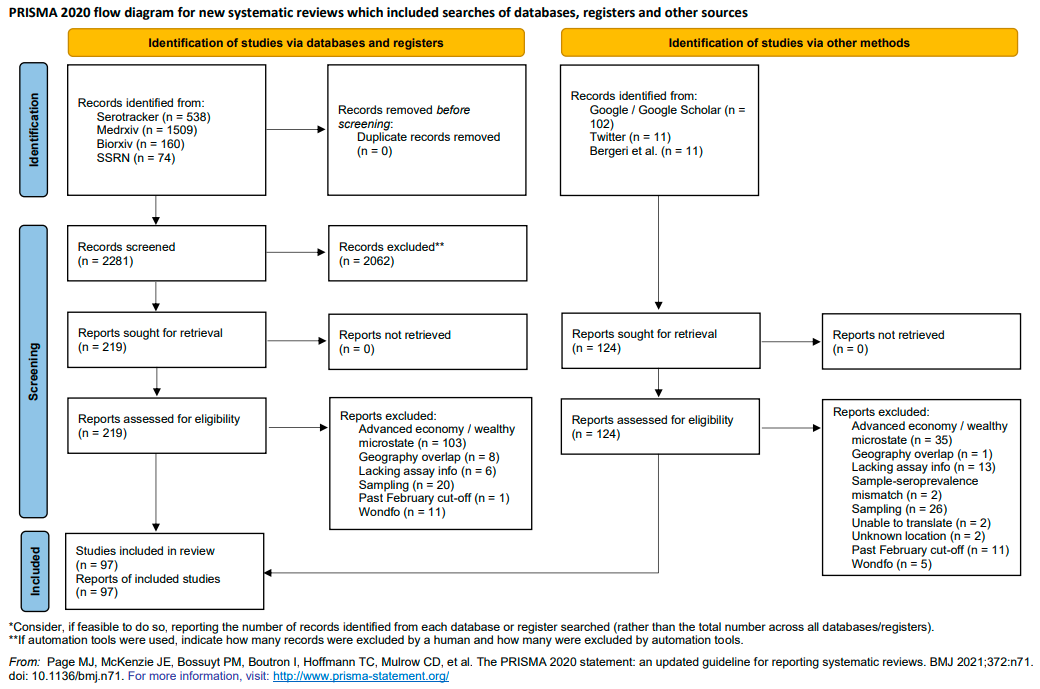

**Appendix References**

1.	Bobrovitz N, Arora RK, Yan T, Rahim H, Duarte N, Boucher E, et al. Lessons from a rapid systematic review of early SARS-CoV-2 serosurveys. medRxiv. 2020:2020.05.10.20097451.

2.	Chen X, Chen Z, Azman AS, Deng X, Sun R, Zhao Z, et al. Serological evidence of human infection with SARS-CoV-2: a systematic review and meta-analysis. The Lancet Global Health.

3.	Byambasuren O, Dobler CC, Bell K, Rojas DP, Clark J, McLaws M-L, et al. Comparison of seroprevalence of SARS-CoV-2 infections with cumulative and imputed COVID-19 cases: Systematic review. PLOS ONE. 2021;16(4):e0248946.

4.	Levin AT, Hanage WP, Owusu-Boaitey N, Cochran KB, Walsh SP, Meyerowitz-Katz G. Assessing the age specificity of infection fatality rates for COVID-19: systematic review, meta-analysis, and public policy implications. European Journal of Epidemiology. 2020;35(12):1123-38.

5.	COVID-19 Cases: Ministerio de Salud Argentina; [Available from: <http://datos.salud.gob.ar/dataset/covid-19-casos-registrados-en-la-republica-argentina/archivo/fd657d02-a33a-498b-a91b-2ef1a68b8d16>.

6.	COVID-19 in Colombia: Instituto Nacional De Salud; 2021 [Available from: <https://www.ins.gov.co/Noticias/paginas/coronavirus.aspx>.

7.	COVID-19 PANEL - ESPIRITO SANTO STATE 2021 [Available from: <https://coronavirus.es.gov.br/painel-covid-19-es>.

8.	Coronavirus - COVID-19: Parana Governo Do Estado; 2021 [Available from: <https://www.saude.pr.gov.br/Pagina/Coronavirus-COVID-19>.

9.	COVID19 Registro Fallecidos 2021 [Available from: <https://public.tableau.com/app/profile/mspbs/viz/COVID19PY-Registros/FALLECIDOS>.

10.	Karlinsky A, Kobak D. Tracking excess mortality across countries during the COVID-19 pandemic with the World Mortality Dataset. eLife. 2021;10:e69336.

11.	Ramachandran S, Malani A. All-cause mortality during SARS-CoV-2 Pandemic in India: Nationally-representative estimates independent of official death registry. medRxiv. 2021:2021.07.20.21260577.

12.	Watson OJ, Alhaffar M, Mehchy Z, Whittaker C, Akil Z, Brazeau NF, et al. Leveraging community mortality indicators to infer COVID-19 mortality and transmission dynamics in Damascus, Syria. Nature Communications. 2021;12(1):2394.

13.	Dyer O. Covid-19: Russia admits to understating deaths by more than two thirds. BMJ. 2020;371:m4975.

14.	Dyer O. Covid-19: Mexico acknowledges 50 000 more deaths than official figures show. BMJ. 2020;371:m4182.

15.	Technical criteria to update the death toll from COVID-19 in Peru. Peruvian State; 2021.

16.	Nepal Dashboard. World Health Organization; 2021.

17.	COVID-19 MÉXICO Comunicado Técnico Diario.

18.	Fullman N, Barber RM, Abajobir AA, Abate KH, Abbafati C, Abbas KM, et al. Measuring progress and projecting attainment on the basis of past trends of the health-related Sustainable Development Goals in 188 countries: an analysis from the Global Burden of Disease Study 2016. The Lancet. 2017;390(10100):1423-59.

19.	Community Assessment for Public Health Emergency Response Toolkit. CDC; 2019.

20.	World Economic and Financial Surveys World Economic Outlook Database—WEO Groups and Aggregates Information. 2021.

21.	Gajda M, Kowalska M, Zejda JE. Impact of Two Different Recruitment Procedures (Random vs. Volunteer Selection) on the Results of Seroepidemiological Study (SARS-CoV-2). International Journal of Environmental Research and Public Health. 2021;18(18):9928.

22.	Knabl L, Mitra T, Kimpel J, Rössler A, Volland A, Walser A, et al. High SARS-CoV-2 seroprevalence in children and adults in the Austrian ski resort of Ischgl. Communications Medicine. 2021;1(1):4.

23.	Laurette M, Marion V, Eduard G, Alex W. Research Square. 2021.

24.	Golding J, Northstone K, Miller LL, Davey Smith G, Pembrey M. Differences between blood donors and a population sample: implications for case-control studies. International journal of epidemiology. 2013;42(4):1145-56.

25.	Pham D, Nguyen D, Nguyen TA, Tran C, Tran L, Devare S, et al. Seroprevalence of HTLV-1/2 Among Voluntary Blood Donors in Vietnam. AIDS research and human retroviruses. 2019;35(4):376-81.

26.	He D, Artzy-Randrup Y, Musa SS, Gräf T, Naveca F, Stone L. The unexpected dynamics of COVID-19 in Manaus, Brazil: Was herd immunity achieved? medRxiv. 2021:2021.02.18.21251809.

27.	Boyce RM, Shook-Sa BE, Aiello AE. A Tale of 2 Studies: Study Design and Our Understanding of Severe Acute Respiratory Syndrome Coronavirus 2 Seroprevalence. Clinical Infectious Diseases. 2020.

28.	Shook-Sa BE, Boyce RM, Aiello AE. Estimation Without Representation: Early Severe Acute Respiratory Syndrome Coronavirus 2 Seroprevalence Studies and the Path Forward. The Journal of Infectious Diseases. 2020;222(7):1086-9.

29.	Rogan WJ, Gladen B. Estimating prevalence from the results of a screening test. Am J Epidemiol. 1978;107(1):71-6.

30.	Lipsitch M, Grad YH, Sette A, Crotty S. Cross-reactive memory T cells and herd immunity to SARS-CoV-2. Nature Reviews Immunology. 2020;20(11):709-13.

31.	Dorigatti I, Lavezzo E, Manuto L, Ciavarella C, Pacenti M, Boldrin C, et al. SARS-CoV-2 antibody dynamics and transmission from community-wide serological testing in the Italian municipality of Vo’. Nature Communications. 2021;12(1):4383.

32.	Kshatri JS, Bhattacharya D, Praharaj I, Mansingh A, Parai D, Kanungo S, et al. Seroprevalence of SARS-CoV-2 in Bhubaneswar, India: findings from three rounds of community surveys. Epidemiology and infection. 2021;149:e139.

33.	Laxmaiah A, Rao NM, Arlappa N, Babu J, Kumar PU, Singh P, et al. SARS-CoV-2 seroprevalence in the city of Hyderabad, India in early 2021. medRxiv. 2021:2021.07.18.21260555.

34.	Wagner R, Peterhoff D, Beileke S, Günther F, Berr M, Einhauser S, et al. Estimates and Determinants of SARS-Cov-2 Seroprevalence and Infection Fatality Ratio Using Latent Class Analysis: The Population-Based Tirschenreuth Study in the Hardest-Hit German County in Spring 2020. Viruses. 2021;13(6):1118.

35.	Sharma N, Sharma P, Basu S, Bakshi R, Gupta E, Agarwal R, et al. Second wave of the Covid-19 pandemic in Delhi, India: high seroprevalence not a deterrent? medRxiv. 2021:2021.09.09.21263331.

36.	Álvarez-Antonio C, Meza-Sánchez G, Calampa C, Casanova W, Carey C, Alava F, et al. Seroprevalence of anti-SARS-CoV-2 antibodies in Iquitos, Peru in July and August, 2020: a population-based study. The Lancet Global Health. 2021;9(7):e925-e31.

37.	Fox SJ, Potu P, Lachmann M, Srinivasan R, Meyers LA. The COVID-19 herd immunity threshold is not low: A re-analysis of European data from spring of 2020. medRxiv. 2020:2020.12.01.20242289.

38.	Sharma N, Sharma P, Basu S, Saxena S, Chawla R, Dushyant K, et al. The seroprevalence of severe acute respiratory syndrome coronavirus 2 in Delhi, India: a repeated population-based seroepidemiological study. Transactions of The Royal Society of Tropical Medicine and Hygiene. 2021.

39.	Bellizzi S, Alsawalha L, Sheikh Ali S, Sharkas G, Muthu N, Ghazo M, et al. A three-phase population based sero-epidemiological study: Assessing the trend in prevalence of SARS-CoV-2 during COVID-19 pandemic in Jordan. One Health. 2021;13:100292.

40.	Poustchi H, Darvishian M, Mohammadi Z, Shayanrad A, Delavari A, Bahadorimonfared A, et al. SARS-CoV-2 antibody seroprevalence in the general population and high-risk occupational groups across 18 cities in Iran: a population-based cross-sectional study. The Lancet Infectious Diseases. 2021;21(4):473-81.

41.	Emmerich P, Murawski C, Ehmen C, von Possel R, Pekarek N, Oestereich L, et al. Limited specificity of commercially available SARS-CoV-2 IgG ELISAs in serum samples of African origin. Tropical Medicine & International Health. 2021;26(6):621-31.

42.	Abdella S, Riou S, Tessema M, Assefa A, Seifu A, Blachman A, et al. Prevalence of SARS-CoV-2 in urban and rural Ethiopia:&#xa0;Randomized household serosurveys reveal level of spread during the first wave of the pandemic. EClinicalMedicine. 2021;35.

43.	Peluso MJ, Takahashi S, Hakim J, Kelly JD, Torres L, Iyer NS, et al. SARS-CoV-2 antibody magnitude and detectability are driven by disease severity, timing, and assay. medRxiv. 2021:2021.03.03.21251639.

44.	Steinhardt LC, Ige F, Iriemenam NC, Greby SM, Hamada Y, Uwandu M, et al. Cross-Reactivity of Two SARS-CoV-2 Serological Assays in a Setting Where Malaria Is Endemic. Journal of Clinical Microbiology. 2021;59(7):e00514-21.

45.	Alemu BN, Addissie A, Mamo G, Deyessa N, Abebe T, Abagero A, et al. Sero-prevalence of anti-SARS-CoV-2 Antibodies in Addis Ababa, Ethiopia. bioRxiv. 2020:2020.10.13.337287.

46.	Nkuba AN, Makiala SM, Guichet E, Tshiminyi PM, Bazitama YM, Yambayamba MK, et al. High Prevalence of Anti–Severe Acute Respiratory Syndrome Coronavirus 2 (Anti–SARS-CoV-2) Antibodies After the First Wave of Coronavirus Disease 2019 (COVID-19) in Kinshasa, Democratic Republic of the Congo: Results of a Cross-sectional Household-Based Survey. Clinical Infectious Diseases. 2021.

47.	Wiens KE, Mawien PN, Rumunu J, Slater D, Jones FK, Moheed S, et al. Seroprevalence of Severe Acute Respiratory Syndrome Coronavirus 2 IgG in Juba, South Sudan, 2020(1). Emerg Infect Dis. 2021;27(6):1598-606.

48.	Gelman A, Carpenter B. Bayesian analysis of tests with unknown specificity and sensitivity. medRxiv. 2020:2020.05.22.20108944.

49.	Silveira MF, Mesenburg MA, Dellagostin OA, de Oliveira NR, Maia MA, Santos FD, et al. Time-dependent decay of detectable antibodies against SARS-CoV-2: A comparison of ELISA with two batches of a lateral-flow test. Braz J Infect Dis. 2021;25(4):101601-.

50.	Conklin SE, Martin K, Manabe YC, Schmidt HA, Miller J, Keruly M, et al. Evaluation of Serological SARS-CoV-2 Lateral Flow Assays for Rapid Point-of-Care Testing. Journal of clinical microbiology. 2021;59(2):e02020-20.

51.	Hartwig FP, Vidaletti LP, Barros AJD, Victora GD, Menezes AMB, Mesenburg MA, et al. Combining serological assays and official statistics to describe the trajectory of the COVID-19 pandemic: results from the EPICOVID19-RS study in Rio Grande do Sul (Southern Brazil). medRxiv. 2021:2021.05.21.21257634.

52.	Stefanelli P, Bella A, Fedele G, Fiore S, Pancheri S, Benedetti E, et al. Longevity of seropositivity and neutralizing titers among SARS-CoV-2 infected individuals after 4 months from baseline: a population-based study in the province of Trento. medRxiv. 2020:2020.11.11.20229062.

53.	Pérez-Olmeda M, Saugar JM, Fernández-García A, Pérez-Gómez B, Pollán M, Avellón A, et al. Evolution of antibodies against SARS-CoV-2 over seven months: experience of the Nationwide Seroprevalence ENE-COVID Study in Spain. medRxiv. 2021:2021.03.11.21253142.

54.	NEW STATEWIDE DATA SHOW EVIDENCE OF FOUR-FOLD INCREASE IN RECENT COVID-19 INFECTIONS. University of Wisconsin-Madison; 2020.

55.	Barchuk A, Shirokov D, Sergeeva M, Tursun­zade R, Dudkina O, Tychkova V, et al. Evaluation of the performance of SARS-­CoV­-2 antibody assays for a longitudinal population­based study of COVID-­19 spread in St. Petersburg, Russia. Journal of Medical Virology. 2021;93(10):5846-52.

56.	Beverland A, Keogan M, Connell J, De Gascun C, Igoe D. Longitudinal Serological Analysis Following a National Seroprevalence Study to Investigate COVID-19 Infection in People Living in Ireland. The Journal of Infectious Diseases. 2021;224(6):1100-1.

57.	Carreño JM, Mendu DR, Simon V, Shariff MA, Singh G, Menon V, et al. Longitudinal analysis of SARS-CoV-2 seroprevalence using multiple serology platforms. medRxiv. 2021:2021.02.24.21252340.

58.	Kahre E, Galow L, Unrath M, Haag L, Blankenburg J, Dalpke AH, et al. Kinetics and seroprevalence of SARS-CoV-2 antibodies – a comparison of 3 different assays. medRxiv. 2021:2021.03.10.21253273.

59.	Thiruvengadam R, Chattopadhyay S, Mehdi F, Desiraju BK, Chaudhuri S, Singh S, et al. Longitudinal serology in SARS-CoV-2 infected individuals in India – a prospective cohort study. medRxiv. 2021:2021.02.04.21251140.

60.	Muecksch F, Wise H, Batchelor B, Squires M, Semple E, Richardson C, et al. Longitudinal Serological Analysis and Neutralizing Antibody Levels in Coronavirus Disease 2019 Convalescent Patients. The Journal of Infectious Diseases. 2020;223(3):389-98.

61.	Sim M, Cockcroft C, Darby D, Ellis CR, Heaps A, Scargill J, et al. Paired sensitivity analysis of four SARS-CoV-2 serological immunoassays in a longitudinal cohort of convalescent hospital staff. Annals of Clinical Biochemistry. 2021:00045632211030957.

62.	Gallais F, Gantner P, Bruel T, Velay A, Planas D, Wendling M-J, et al. Evolution of human antibody responses up to one year after SARS-CoV-2 infection. medRxiv. 2021:2021.05.07.21256823.

63.	Perez-Saez J, Zaballa M-E, Yerly S, Andrey DO, Meyer B, Eckerle I, et al. Persistence and detection of anti-SARS-CoV-2 antibodies: immunoassay heterogeneity and implications for serosurveillance. medRxiv. 2021:2021.03.16.21253710.

64.	Ladage D, Rösgen D, Schreiner C, Ladage D, Adler C, Harzer O, et al. Persisting Antibody Response to SARS-CoV-2 in a Local Austrian Population. Frontiers in Medicine. 2021;8(881).

65.	Choe PG, Kim K-H, Kang CK, Suh HJ, Kang E, Lee SY, et al. Antibody Responses 8 Months after Asymptomatic or Mild SARS-CoV-2 Infection. Emerging Infectious Disease journal. 2021;27(3):928.

66.	Pagotto V, Luna L, Salto J, Manslau MW, Figar S, Mistchenko AS, et al. Long-Term Duration of Antibody Response to SARS CoV-2 in One of the Largest Slums of Buenos Aires. medRxiv. 2021:2021.03.05.21253010.

67.	Rodeles LM PL, Benitez R, Benzaquen N, Serravalle P, Long AK, et al. Seroprevalence of anti-SARS-CoV-2 IgG in asymptomatic and pauci-symptomatic people over a 5 month survey in Argentina. Rev Panama Salud Publica. 2021;45(66).

68.	den Hartog G, Vos ERA, van den Hoogen LL, van Boven M, Schepp RM, Smits G, et al. Persistence of Antibodies to Severe Acute Respiratory Syndrome Coronavirus 2 in Relation to Symptoms in a Nationwide Prospective Study. Clinical Infectious Diseases. 2021.

69.	Radon K, Bakuli A, Pütz P, Gleut RL, Guggenbuehl Noller JM, Olbrich L, et al. From first to second wave: follow-up of the prospective Covid-19 cohort (KoCo19) in Munich (Germany). medRxiv. 2021:2021.04.27.21256133.

70.	He Z, Ren L, Yang J, Guo L, Feng L, Ma C, et al. Seroprevalence and humoral immune durability of anti-SARS-CoV-2 antibodies in Wuhan, China: a longitudinal, population-level, cross-sectional study. The Lancet. 2021;397(10279):1075-84.

71.	Goto A, Go H, Miyakawa K, Yamaoka Y, Ohtake N, Kubo S, et al. Sustained Neutralizing Antibodies 6 Months Following Infection in 376 Japanese COVID-19 Survivors. Frontiers in Microbiology. 2021;12(1039).

72.	Domènech-Montoliu S, Puig-Barberà J, Pac-Sa MR, Vidal-Utrillas P, Latorre-Poveda M, Del Rio-González A, et al. Persistence of Anti-SARS-CoV-2 Antibodies Six Months after Infection in an Outbreak with Five Hundred COVID-19 Cases in Borriana (Spain): A Prospective Cohort Study. COVID. 2021;1(1):71-82.

73.	Isho B, Abe KT, Zuo M, Jamal AJ, Rathod B, Wang JH, et al. Persistence of serum and saliva antibody responses to SARS-CoV-2 spike antigens in COVID-19 patients. Science Immunology. 2020;5(52):eabe5511.

74.	Alvim RGF, Lima TM, Rodrigues DAS, Marsili FF, Bozza VBT, Higa LM, et al. Development and large-scale validation of a highly accurate SARS-COV-2 serological test using regular test strips for autonomous and affordable finger-prick sample collection, transportation, and storage. medRxiv. 2021:2020.07.13.20152884.

75.	Oxford University researchers release cheap, quick COVID-19 antibody test: University of Oxford; 2021 [Available from: <https://www.ox.ac.uk/news/2021-03-29-oxford-university-researchers-release-cheap-quick-covid-19-antibody-test>.

76.	Townsend A, Rijal P, Xiao J, Tan TK, Huang K-YA, Schimanski L, et al. A haemagglutination test for rapid detection of antibodies to SARS-CoV-2. Nature Communications. 2021;12(1):1951.

77.	UK Biobank COVID-19 antibody study: final results. In: Government U, editor. 2021.

78.	Over 1 million St. Petersburg residents have had COVID-19 since the onset of the pandemic: European University at St. Petersburg; 2020 [Available from: <https://eusp.org/en/news/over-1-million-st-petersburg-residents-have-had-covid-19-since-the-onset-of-the-pandemic>.

79.	Petersen MS, Hansen CB, Fríðheim Kristiansen M, Fjallsbak JP, Larsen S, Hansen JL, et al. SARS-CoV-2 natural antibody response persists up to 12 months in a nationwide study from the Faroe Islands. medRxiv. 2021:2021.04.19.21255720.

80.	Gudbjartsson DF, Helgason A, Jonsson H, Magnusson OT, Melsted P, Norddahl GL, et al. Spread of SARS-CoV-2 in the Icelandic Population. N Engl J Med. 2020.

81.	The Germans and Corona: ifo INSTITUT; 2020 [Available from: <https://www.ifo.de/publikationen/2020/monographie-autorenschaft/die-deutschen-und-corona>.

82.	Mutevedzi PC, Kawonga M, Kwatra G, Moultrie A, Baillie V, Mabena N, et al. Estimated SARS-CoV-2 infection rate and fatality risk in Gauteng Province, South Africa: a population-based seroepidemiological survey. International Journal of Epidemiology. 2021.

83.	Perez-Saez J, Lauer SA, Kaiser L, Regard S, Delaporte E, Guessous I, et al. Serology-informed estimates of SARS-COV-2 infection fatality risk in Geneva, Switzerland. medRxiv. 2020:2020.06.10.20127423.

84.	Ng DL, Goldgof GM, Shy BR, Levine AG, Balcerek J, Bapat SP, et al. SARS-CoV-2 seroprevalence and neutralizing activity in donor and patient blood. Nature Communications. 2020;11(1):4698.

85.	Aziz NA, Corman VM, Echterhoff AKC, Müller MA, Richter A, Schmandke A, et al. Seroprevalence and correlates of SARS-CoV-2 neutralizing antibodies from a population-based study in Bonn, Germany. Nature Communications. 2021;12(1):2117.

86.	Parrott JC, Maleki AN, Vassor VE, Osahan S, Hsin Y, Sanderson M, et al. Prevalence of SARS-CoV-2 Antibodies in New York City Adults, June–October 2020: A Population-Based Survey. The Journal of Infectious Diseases. 2021;224(2):188-95.

87.	Lu N, Cheng K-W, Qamar N, Huang K-C, Johnson JA. Weathering COVID-19 storm: Successful control measures of five Asian countries. American Journal of Infection Control. 2020;48(7):851-2.

88.	Okell LC, Verity R, Watson OJ, Mishra S, Walker P, Whittaker C, et al. Have deaths from COVID-19 in Europe plateaued due to herd immunity? The Lancet. 2020;395(10241):e110-e1.

89.	Fuller JA HA, Victory KR, et al. Mitigation Policies and COVID-19–Associated Mortality — 37 European Countries, January 23–June 30, 2020. MMWR Morb Mortal Wkly Rep. 2020;70:58-62.

90.	Miyawaki A, Tsugawa Y. Health and Public Health Implications of COVID-19 in Asian Countries. Asian Economic Policy Review.n/a(n/a).

91.	Flower B, Marks M. Did Laos really control the transmission of SARS-CoV-2 in 2020? The Lancet Regional Health – Western Pacific. 2021;13.

92.	Lee K, Jo S, Lee J. Seroprevalence of SARS-CoV-2 antibodies in South Korea. Journal of the Korean Statistical Society. 2021;50(3):891-904.

93.	Jayasundara P, Peariasamy KM, Law KB, Abd Rahim KNK, Lee SW, Ghazali IMM, et al. Sustaining effective COVID-19 control in Malaysia through large-scale vaccination. medRxiv. 2021:2021.07.05.21259999.

94.	Meyerowitz-Katz G, Merone L. A systematic review and meta-analysis of published research data on COVID-19 infection fatality rates. International Journal of Infectious Diseases. 2020;101:138-48.

95.	Brazeau N, Verity R, Jenks S, Fu H, Whittaker C, Winskill P, et al. Report 34: COVID-19 infection fatality ratio: estimates from seroprevalence. Imperial College London; 2020.

96.	Cai R, Novosad P, Tandel V, Asher S, Malani A. Representative estimates of COVID-19 infection fatality rates from four locations in India: cross-sectional study. BMJ Open. 2021;11(10):e050920.

97.	Barchuk A, Skougarevskiy D, Kouprianov A, Shirokov D, Dudkina O, Tursun-zade R, et al. COVID-19 pandemic in Saint Petersburg, Russia: combining surveillance and population-based serological study data in May, 2020–April, 2021. medRxiv. 2021:2021.07.31.21261428.
