## Supplementary material for "Assessing the Burden of COVID-19 in Developing Countries: Systematic Review, Meta-Analysis, and Public Policy Implications": IFR supplement

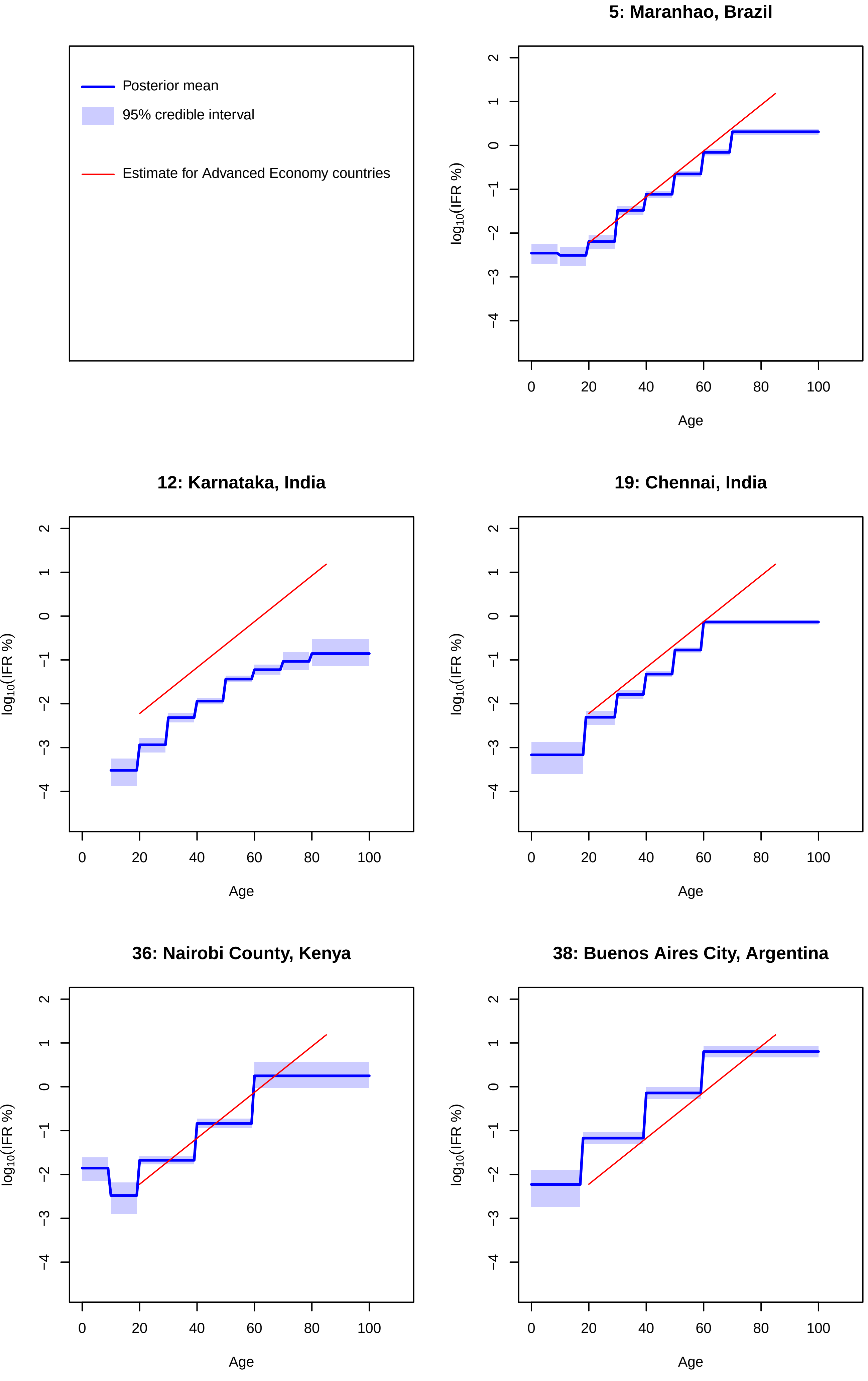
Posterior mean
95% credible interval
Estimate for Advanced Economy countries
5: Maranhao, Brazil
12: Karnataka, India
19: Chennai, India
36: Nairobi County, Kenya
38: Buenos Aires City, Argentina
$\log_{10}$(IFR %)
Age
0
20
40
60
80
100
2
1
0
−1
−2
−3
−4

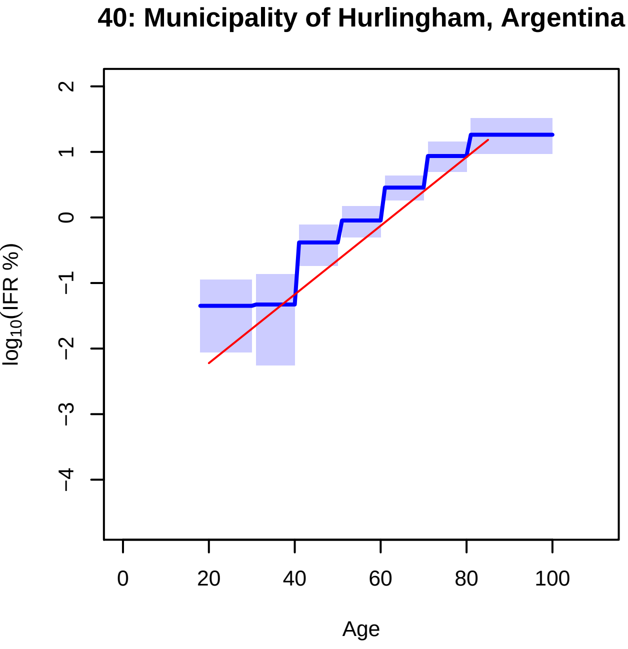
40: Municipality of Hurlingham, Argentina
log10(IFR %)
2
1
0
−1
−2
−3
−4
0
20
40
60
80
100
Age

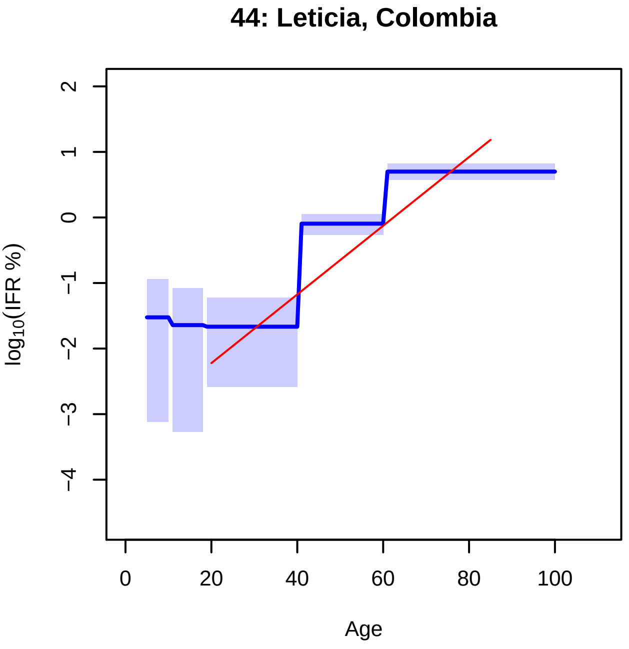
44: Leticia, Colombia
log10(IFR %)
2
1
0
−1
−2
−3
−4
0
20
40
60
80
100
Age

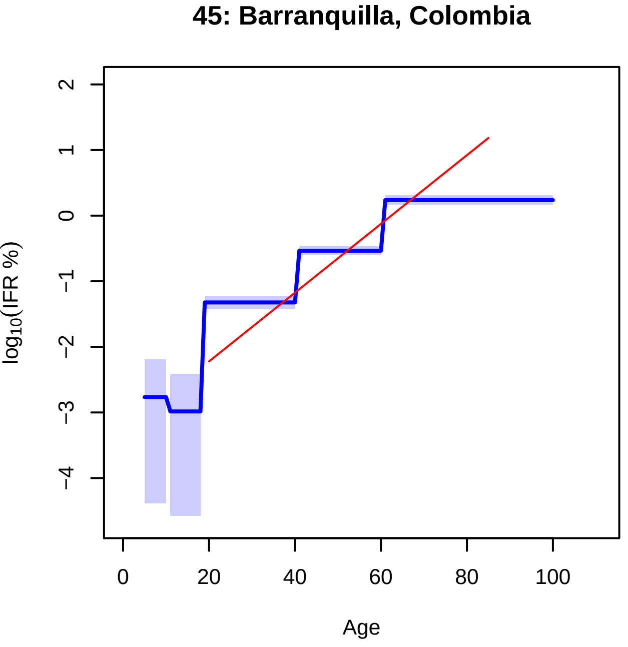
45: Barranquilla, Colombia
log10(IFR %)
2
1
0
−1
−2
−3
−4
0
20
40
60
80
100
Age

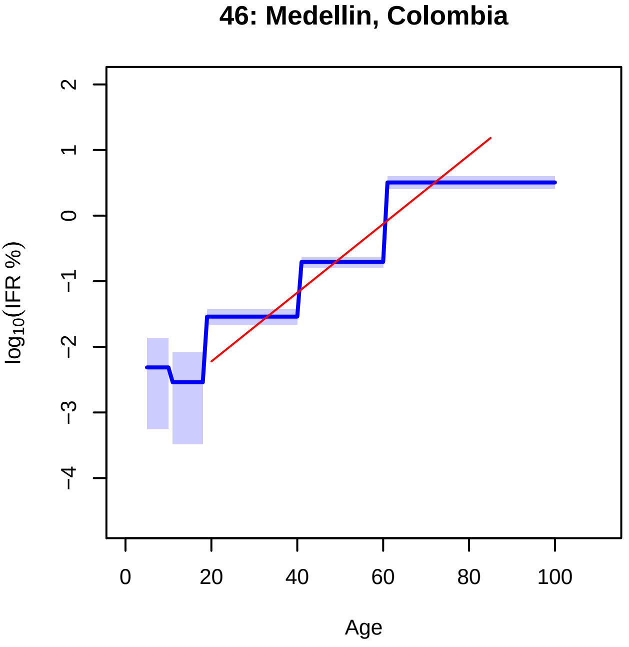
46: Medellin, Colombia
log10(IFR %)
2
1
0
−1
−2
−3
−4
0
20
40
60
80
100
Age

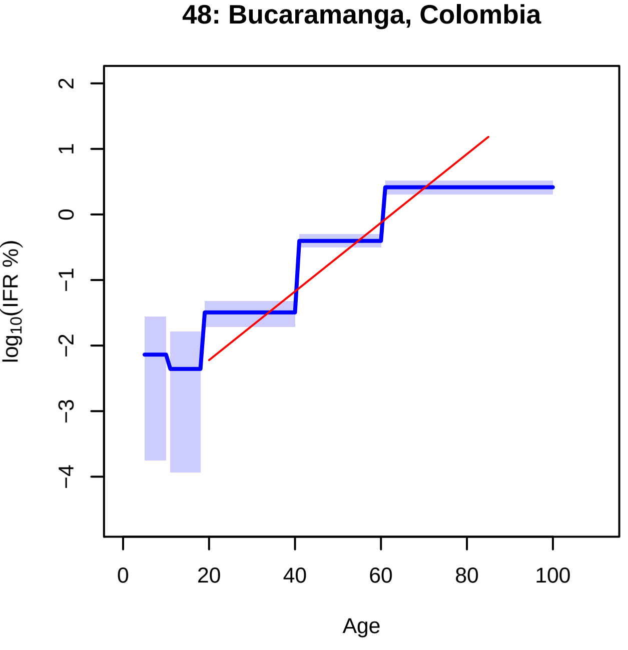
48: Bucaramanga, Colombia
log10(IFR %)
2
1
0
−1
−2
−3
−4
0
20
40
60
80
100
Age

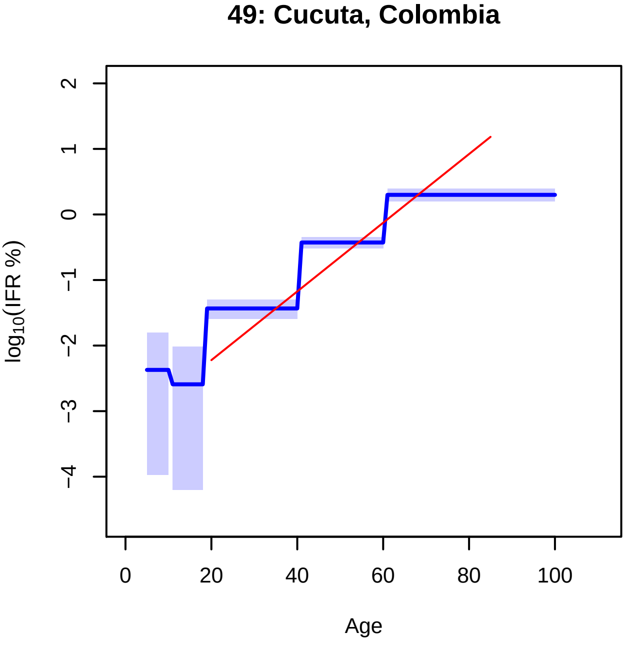
49: Cucuta, Colombia
log10(IFR %)
2
1
0
−1
−2
−3
−4
0
20
40
60
80
100
Age

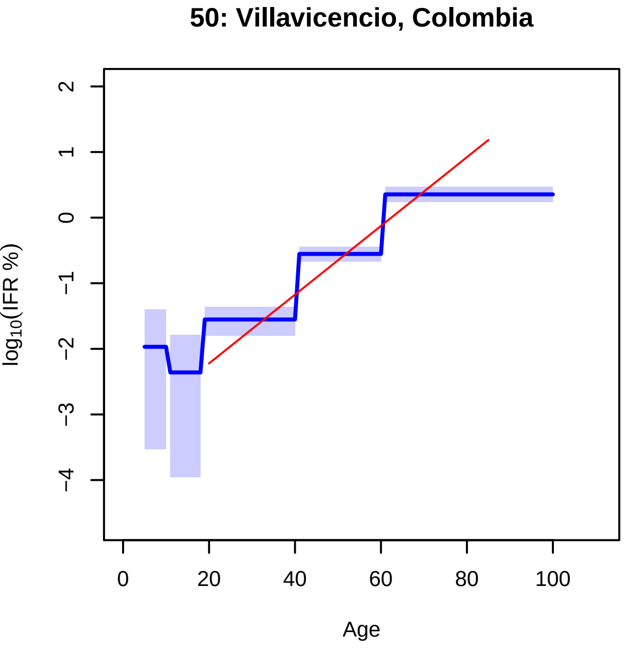
50: Villavicencio, Colombia
log10(IFR %)
Age
2
1
0
−1
−2
−3
−4
0
20
40
60
80
100
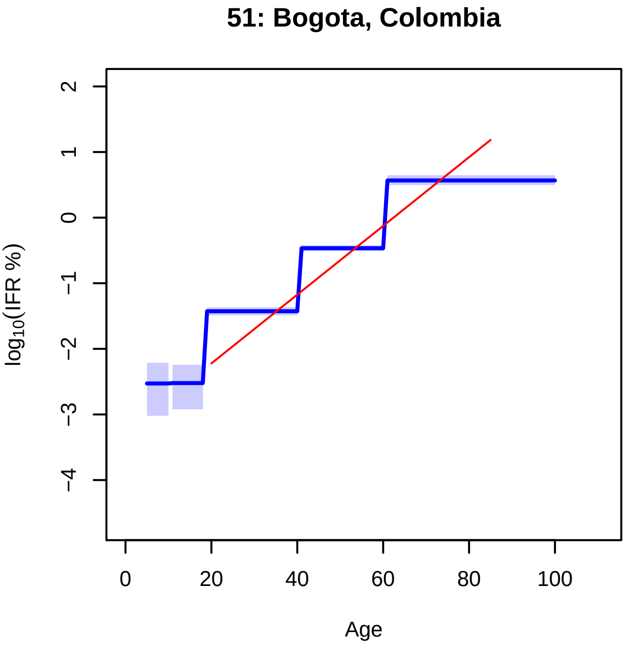
51: Bogota, Colombia
log10(IFR %)
Age
2
1
0
−1
−2
−3
−4
0
20
40
60
80
100
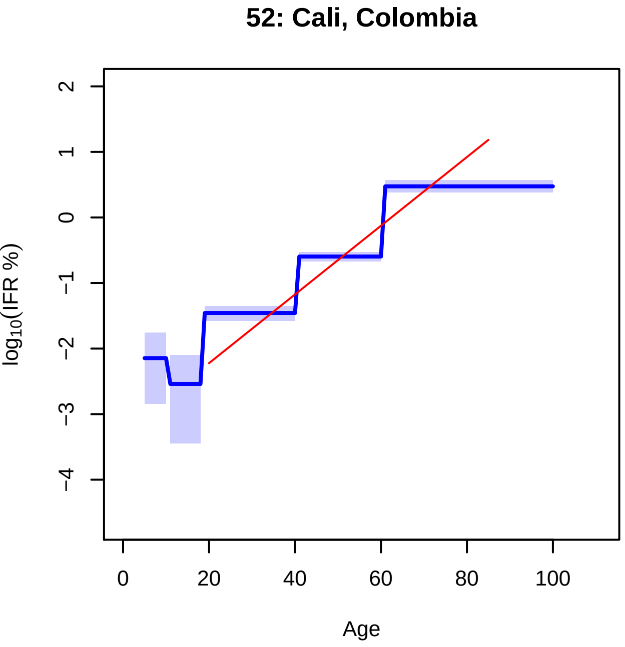
52: Cali, Colombia
log10(IFR %)
Age
2
1
0
−1
−2
−3
−4
0
20
40
60
80
100
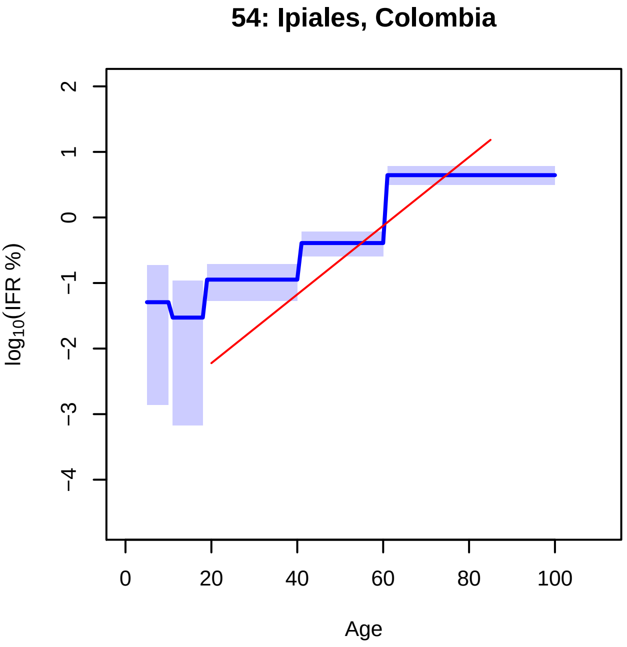
54: Ipiales, Colombia
log10(IFR %)
Age
2
1
0
−1
−2
−3
−4
0
20
40
60
80
100
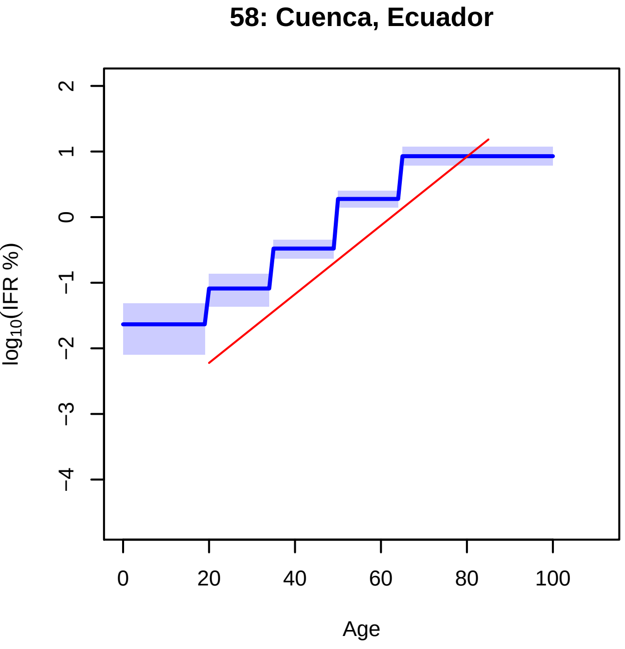
58: Cuenca, Ecuador
log10(IFR %)
Age
2
1
0
−1
−2
−3
−4
0
20
40
60
80
100
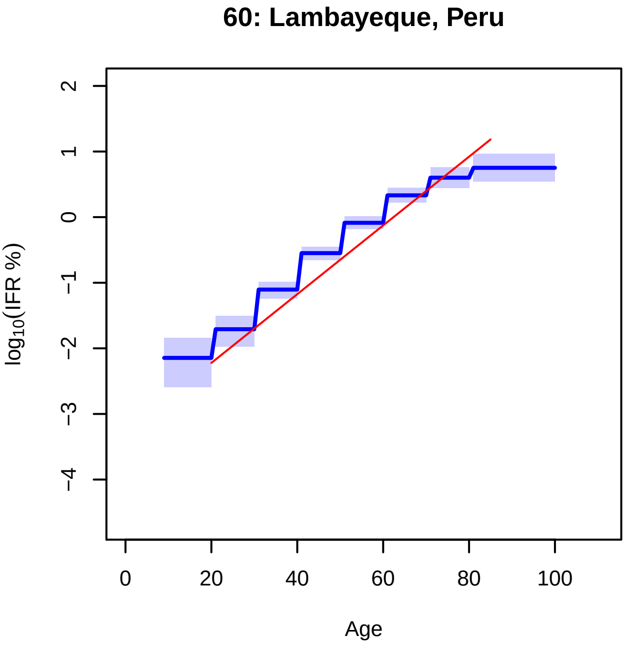
60: Lambayeque, Peru
log10(IFR %)
Age
2
1
0
−1
−2
−3
−4
0
20
40
60
80
100

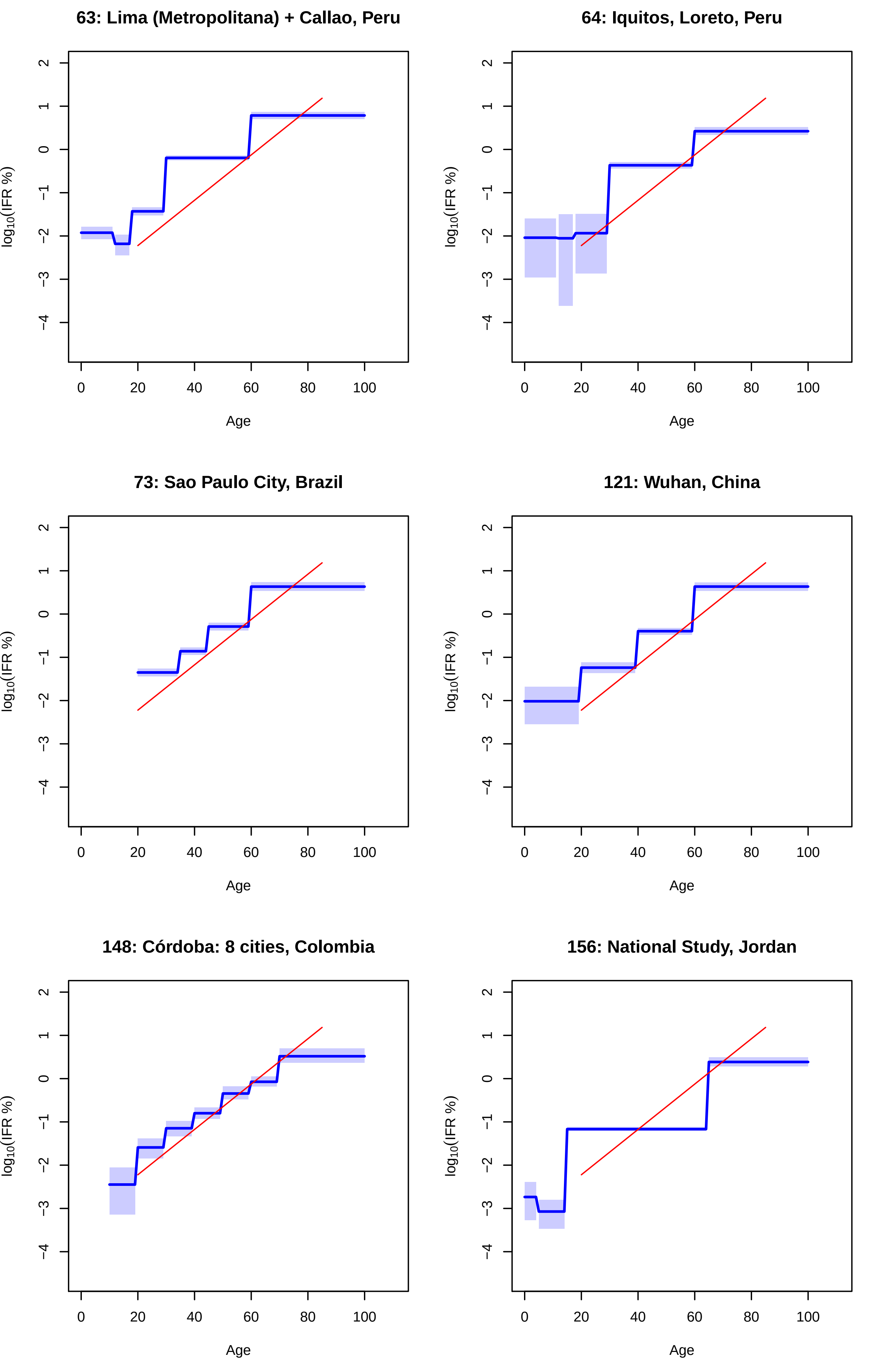

63: Lima (Metropolitana) + Callao, Peru
64: Iquitos, Loreto, Peru
73: Sao Paulo City, Brazil
121: Wuhan, China
148: Córdoba: 8 cities, Colombia
156: National Study, Jordan
$\log_{10}$(IFR %)
Age

**192: National Study, Hungary**

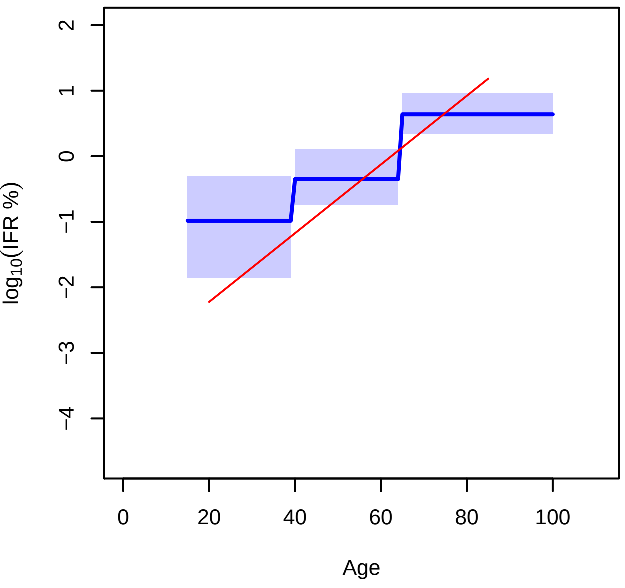


**209: Cuiabá, Mato Grosso, Brazil**

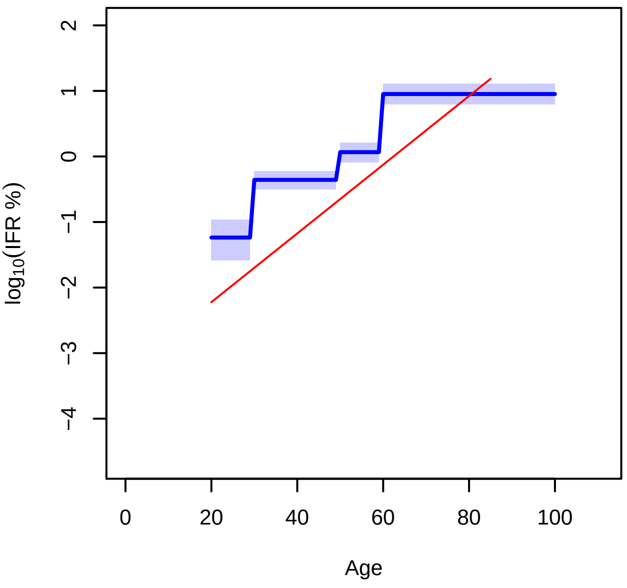


**216: Várzea Grande, Mato Grosso, Brazil**

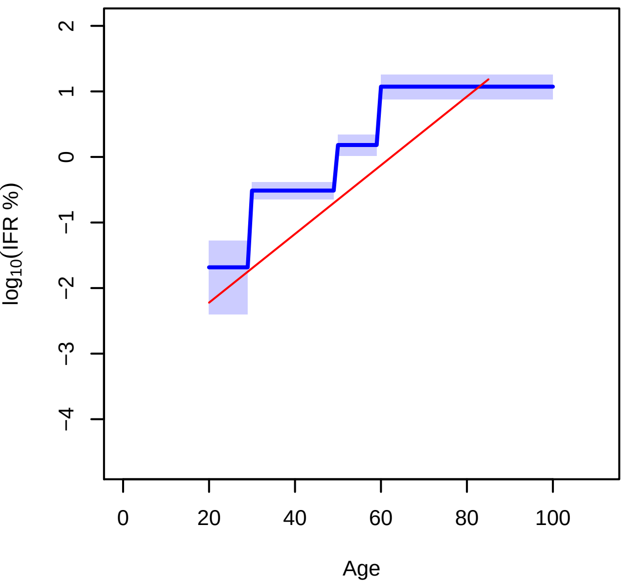


**219: 3 cities, Chile**

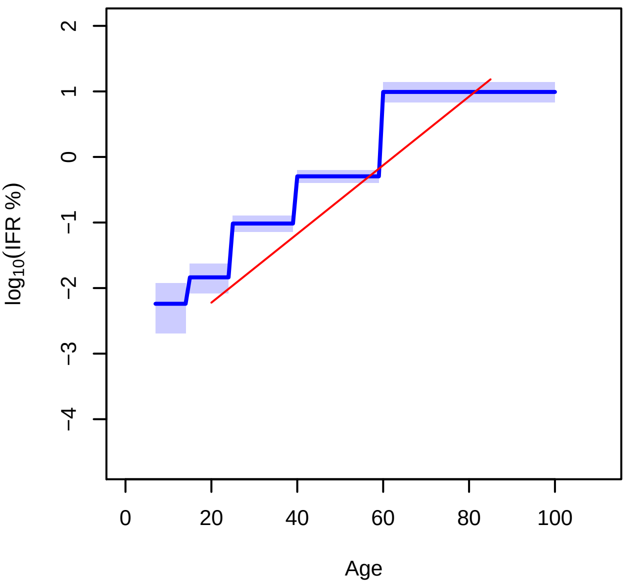


**221: Asunción + Central Department, Paraguay**

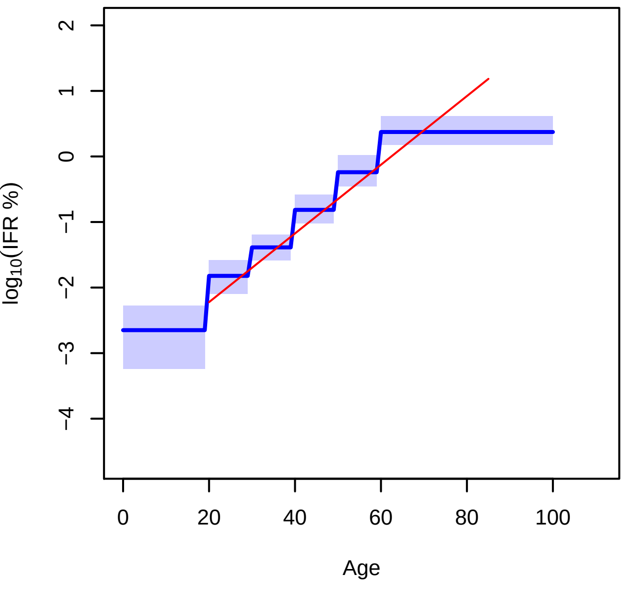
